# Multimodal structural neuroimaging data unveil data-driven subtypes of treatment-resistant depression

**DOI:** 10.1101/2023.09.12.23295423

**Authors:** Federica Colombo, Federico Calesella, Beatrice Bravi, Lidia Fortaner-Uyà, Camilla Monopoli, Emma Tassi, Matteo Carminati, Raffaella Zanardi, Irene Bollettini, Sara Poletti, Paolo Brambilla, Alessandro Serretti, Eleonora Maggioni, Chiara Fabbri, Francesco Benedetti, Benedetta Vai

## Abstract

**Background:** An estimated 30% of Major Depressive Disorder (MDD) patients exhibit resistance to conventional antidepressant treatments. Identifying reliable biomarkers of treatment-resistant depression (TRD) represents a major goal of precision psychiatry, which is hampered by the clinical and biological heterogeneity underlying MDD.

**Methods:** To parse heterogeneity and uncover biologically-driven subtypes of MDD, we applied an unsupervised data-driven framework to stratify 102 MDD patients on their neuroimaging signature, including extracted measures of cortical thickness, grey matter volumes, and white matter fractional anisotropy. Our novel analytical pipeline integrated different machine learning algorithms to harmonize neuroimaging data, perform data dimensionality reduction, and provide a stability-based relative clustering validation. The obtained clusters were then characterized for TRD, history of childhood trauma and different profiles of depressive symptoms.

**Results:** Our results indicated two different clusters of patients, differentiable with 67% of accuracy: 1) one cluster (n=59) was associated with a higher proportion of TRD compared to the other, and higher scores of energy-related depressive symptoms, history of childhood abuse and emotional neglect; this cluster showed a widespread reduction in cortical thickness and volumes, along with fractional anisotropy in the right superior fronto-occipital fasciculus, stria terminalis, and corpus callosum; 2) the second cluster (n=43) was associated with cognitive and affective depressive symptoms and thicker cortices and wider volumes compared to the other.

**Discussion:** Our stratification of MDD patients based on structural neuroimaging identified clinically-relevant subgroups of TRD with specific symptomatic and childhood trauma profiles, which are informative for tailoring personalized and more effective interventions of treatment resistance.

## Introduction

Major depressive disorder (MDD) represents one of the most common psychiatric disorders worldwide, with almost 17% of men and 25% of women experiencing at least one depressive episode during their lifetime (1). One of the most alarming issues contributing to the social and economic burden of the disorder is the uncertainty linked to clinical outcomes: despite the wide variability in treatment modalities, about 30% of patients do not achieve remission after antidepressant treatments (2) or relapse soon after, paving the way to treatment-resistant depression (TRD) (3). The lack of response to at least one antidepressant treatment is associated with more severe clinical outcomes, such as higher risk of suicide, more frequent relapses, psychiatric and medical comorbidities, health deterioration and functional impairment (4, 5). The prediction of clinical outcomes is further complicated by the absence of objective criteria for treatment choice, and several trials are often needed to find the optimal treatment for a patient (6). Indeed, there is a dire need for a “precision medicine” approach to MDD for the development of more effective treatments tailored on the individual profile.

The failure of achieving remission to at least two antidepressant treatments of adequate dose and duration is the most commonly used definition of TRD (7), and a recent general consensus on the clinical definition of TRD recommended future subgroup or stratified analyses on biological samples, because no biomarker has yet been validated to identify people with TRD (3). One of the main obstacles hampering the development of personalized treatment strategies is indeed the clinical and biological heterogeneity of MDD. Different clinical subtypes of clinical manifestations have been recognized according to established diagnostic criteria (8), and most of them have been associated with TRD (9). Although the heritability of MDD is estimated at 35-40% (10), approximately 67% of variance in MDD is explained by environmental factors (11). An example of such factors is the experience of childhood trauma, which is considered as one of the strongest predictors of poor antidepressant response (12–14). Compared to those not exposed to adverse childhood experiences, individuals with an history of childhood trauma typically have an earlier onset of MDD, more deleterious course, lower rates of remissions and response to treatments (15, 16). However, the extent to which the aetiology of TRD is shared with treatment-responsive depression is still unclear. Therefore, the delineation of objective biomarkers of TRD is of the utmost importance to define novel treatments targeting the underlying neurobiology. In terms of neural correlates, structural neuroimaging studies indicate that volumetric changes in fronto-striatal and hippocampal volumes, together with reduced structural integrity in the corpus callosum and superior longitudinal fasciculus, differentiate TRD from treatment-responsive MDD and healthy controls (17, 18). Nevertheless, most of these findings failed to be consistently replicated, possibly due to the heterogeneity in the assessment of treatment resistance and in statistical methods (19).

As an attempt to uncover subgroups of psychiatric conditions, several studies employed unsupervised machine learning approaches in order to stratify patients based on shared characteristics (20–22). The most common approach is to apply unsupervised clustering algorithms on clinical data. While these studies suggest that specific symptoms-based clusters could be possibly linked to different effectiveness of antidepressant treatments (23–25), the analysis of symptoms only is likely to be poorly informative given the very large heterogeneity of major depression clinical profile (26). An alternative approach is to identify biologically-driven subtypes by grouping patients based on shared neuro-biological features (27). Recent studies applied clustering techniques on resting-state (28–31) and structural neuroimaging data (32, 33), providing new insights in how distinct neurobiological signatures could be linked to various clinical manifestations of depression. Despite the variability in the number of subtypes, most of the studies converge on the identification of anxiety-, anhedonia-, and insomnia-related data-driven clusters, as well as on clusters’ differences in depression severity and recurrence (34). Some of these “biotypes” were found to be also predictive of response to antidepressant treatments (28), but they failed to be replicated in independent samples (35). Indeed, a common difficulty in the implementation of unsupervised algorithms is the lack of ground truth information and the absence of a defined approach to assess clustering validity, which further complicates the replicability of subtypes nested within clinical populations (36, 37). Moreover, none of these studies combined different neuroimaging modalities to investigate whether multimodal information could enhance the discovery of relevant subgroups.

In the current study, we aim to discover biologically-driven subtypes of patients with different levels of antidepressant treatment response based on multimodal structural neuroimaging, including grey matter volumes, cortical thickness, and extracted fractional anisotropy (FA) values of white matter tracts, in MDD patients. We implemented a novel analytical pipeline including data harmonization, dimensionality reduction, and stability-based relative clustering validation to identify clusters of patients that are stable and generalizable to unseen observations (38). The identified data-driven clusters were then profiled for clinical variables of interest, including TRD, depressive symptomatology, and history of childhood trauma.

## Materials and Methods

### Participants

A total of 102 patients with a current diagnosis of MDD and an ongoing depressive episode were recruited at IRCCS San Raffaele Scientific Institute (Milan, Italy). Detailed inclusion and exclusion criteria are reported in Methods S1. After a complete description of the study, written informed consent was obtained. All procedures contributing to this work comply with the ethical standards of the relevant national and institutional committees on human experimentation and with the Helsinki Declaration of 1975, as revised in 2008. The study is approved by the local ethical committee.

### Clinical and sociodemographic measures

Age, sex, number of previous mood episodes, age of onset, duration of illness, body mass index (BMI), and years of education were collected. To obtain a measure for pharmacological treatment, medication load was calculated for each subject categorizing each medication into low-dose to high-dose groupings, scored as 0 (no medication), 1–4 (low to high dosages) as described by Sackeim (39) (see Methods S2 for detailed criteria). Treatment resistance was defined as a failure to respond to at least two antidepressant treatments of adequate dosages and duration (3, 7). Clinical data were assessed by the psychiatrist in charge using best estimation procedure, taking into account available charts, case notes, and information provided by at least one relative (40). In a subsample of 64 patients, severity of depression was rated on the Beck Depression Inventory–Short Form (BDI-SF) (41), and composite scores for different domains (Negative Self-Esteem, Anergy, and Dysphoria) were derived (42). Childhood trauma was evaluated using the 28-items Childhood Trauma Questionnaire (CTQ) (43). CTQ is a self-administrated inventory that was developed for a reliable retrospective assessment of neglect and abuse during childhood. The questionnaire is composed of 5 subscales that evaluate different aspects of neglect and abuse (physical, sexual, and emotional abuse, and physical and emotional neglect). Moreover, a Minimization/Denial validity scale was used to identify underreported maltreatment (44).

### MRI data acquisition and pre-processing

All subjects underwent a magnetic resonance scan at C.E.R.M.A.C. (Centro di Eccellenza Risonanza Magnetica ad Alto Campo, University Vita-Salute San Raffaele, Milan, Italy). T1-weighted and diffusion tensor images (DTI) were acquired on two 3.0 Tesla scanners. Technical details for the two scanners are reported in Methods S3.

T1-weighted neuroanatomical images were processed using the Computational Anatomy Toolbox (CAT12) for SPM (45). T1 images were normalized to an anatomical model and segmented into gray matter, white matter, and cerebrospinal fluid (CSF). Check of spatial alignment and sample homogeneity was performed to exclude outliers. Then, low-pass spatial filtering (smoothing) techniques were used to remove any potential artefacts in the tissue maps. Cortical thickness was derived for 68 regions defined by the Desikan-Killiany atlas (46) and 148 from the Destrieux atlas (47), whereas volumes of cortical and subcortical grey matter volumes were extracted for 122 regions labelled by the Neuromorphometrics atlas (http://Neuromorphometrics.com/). Finally, total intracranial volume (TIV) was computed. Whole-brain tract-wise average fractional anisotropy (FA) values were extracted according to ENIGMA-DTI protocols (http://enigma.ini.usc.edu/protocols/dtiprotocols/). DTI images were pre-processed using FMRIB Software Library (FSL) tools. Specifically, all volumes were corrected for eddy current induced distortions and subjects’ movements (48). Then, a brain mask was created using Brain Extraction Tool (BET) (49), which deletes non-brain tissues from the image. Next, by FSL’s DTIFIT command, included in FDT (50), a voxel-wise diffusion tensor model was fit to the data in order to obtain parametric maps of FA. All subject’s FA images were then processed using FSL’s TBSS analytic method and were aligned to the MNI space, by using local deformation procedures performed by FMRIB’s FNIRT. The mean of all aligned FA was then created, and a “thinning” process was applied to create a skeletonized mean FA image representing the centers of all common tracts. A threshold of 0.2 was set to this image in order to control for intersubject variability and reduce the likelihood of partial volume effect. Quality control, including inspections of data, vector gradients, registration, and average skeleton projection distance, were performed according to the ENIGMA-DTI protocol. Finally, all individual FA images were projected onto the skeleton by searching perpendicular from the skeleton for maximum FA values (51). Average FA values were calculated from voxels in each subject’s white matter skeleton within 63 tract-wise ROIs, derived from the Johns Hopkins University (JHU) white matter parcellation atlas (52). FA values were separately calculated for right and left hemispheres in each ROI, except for body of corpus callosum (BCC), corpus callosum (CC), fornix (FX), genu of corpus callosum (GCC) and splenium of corpus callosum (SCC).

Since MRI data were acquired with two different scanners, we implemented the ComBat algorithm to correct for potential technical artefacts (i.e., “batch effects”) that could affect meaningful relationships between biological signal and clinical outcomes, leading to unreliable conclusions (53). The ComBat algorithm was adapted to be performed on regional gray matter estimates and extracted tract-based FA values. Of note, age, sex, and TIV (only for grey matter measures) were considered as biological covariates in order to be protected from the removal of scanner effects.

### Statistical analysis

#### Stability-based relative clustering validation

Clustering analyses were performed using the *reval* Python library (https://github.com/IIT-LAND/reval_clustering), which implements a stability-based relative clustering approach within a cross-validation framework to identify the clustering solution that best replicates on unseen data. (38) (For further details, see Methods S4). For the current study, *reval* was adapted to be used on neuroimaging features, choosing a Gaussian mixture model (GMM) with full covariance matrix as clustering algorithm, and support vector machine (SVM) as a supervised classifier. Given the high-dimensionality of the data, we performed the analyses both with and without dimensionality reduction to investigate the impact of multidimensionality on clustering performance (github repository: https://github.com/fede-colombo/NeuReval).

In the first analysis, data were mean centered and normalized to the same scale. Input variables were adjusted for confounding effects of age, sex and TIV (only in grey matter) using linear regression. All these steps were fitted on the training set and then applied to the test set. A 2-folds cross-validation scheme (50% training, 50% test) was implemented to control for the size imbalance that derives from training-test splitting. The cross-validation scheme was repeated 10 times with 10 random labelling iterations to ensure robustness. To optimize the SVM hyperparameters (i.e., soft-margin C ranged 0.01, 0.1, 1, 10, 100, 1000) and the type of kernel (i.e., linear or radial), a grid search was performed in the training set and selection was based on normalized stability. Since previous studies identified from 2 to 4 clusters of MDD based on neuroimaging data (34), the entire cross-validation procedure was iterated over a number of GMM components from 2 to 5. The best clustering solution was the one that minimized the normalized stability throughout cross-validation. To assess the internal validity of the identified clusters, we also computed the silhouette and Davies-Bouldin scores by applying GMM on the entire dataset and with the same range of number of components used in the stability-based regime, and the adjusted mutual information (AMI) index was calculated to evaluate the similarity of clusters’ labels derived from the stability-based approach and the ones obtained from silhouette and Davies-Bouldin scores model’s selection.

The same analysis was repeated by applying Uniform Manifold Approximation and Projection (UMAP) for dimensionality reduction. UMAP was initialised with a number of neighbours equal to 30, minimum distance equal to 0.0, and Euclidean distance metric, as suggested in the documentation (https://umap-learn.readthedocs.io/en/latest/clustering.html). Together with SVM hyperparameters, also the number of UMAP components was optimized within cross-validation through grid search in the training set, iterating over 2 to 5 components.

Despite the stability-based relative clustering approach allows us to assess the stability of a given clustering solution by means of cross-validation, it does not assess whether the identified clusters reflect the existence of true subgroups within the data. Therefore, we further investigated whether the selected clustering solutions significantly deviated from the null hypothesis that data derive from a single multivariate Gaussian distribution computed from 10,000 Monte Carlo simulations using the *sigclust* library in R (54).

#### Clinical, demographic, and neuroimaging characterization of the clusters

For both the analyses, the identified clusters were compared for neuroimaging features, TRD, age, sex, education, type of scanner, number of episodes, age of onset, duration of illness, medication load, BMI, CTQ total score and subscales, BDI total scores and domains, and HDRS-21 total score. Two-sample t-tests were performed for continuous variables, whereas χ2 was calculated for categorical variables. For neuroimaging data, Cohen’s d effects sizes were also computed and comparisons were deemed as significant if they passed a false-discovery rate (FDR) q<0.05 threshold to correct for multiple comparisons.

To investigate whether the data-driven clusters reflect clinically-meaningful subgroups of depressed patients, we performed multivariate analysis of variance (MANOVA) considering BDI-SF domains, and CTQ subscales as dependent variables and clusters’ labels as fixed factors. Age, sex, number of previous episodes, and medication load were entered as covariates to control for potential confounding effects. In case of significant results, subsequent linear discriminant analysis (LDA) was performed to assess how the dependent variables discriminate the clusters. All the analyses were performed in R using the *mancova* (*jmv* library) and *lda* (*MASS* library) functions.

## Results

### Unsupervised data-driven identification of depression clusters based on neuroimaging data

The clinical and demographic characteristics of the sample are shown in Table 1. The stability-based relative validation clustering approach on multimodal structural neuroimaging data without data reduction identified that a 2-clusters solution minimized the normalized stability (0.316), showing a good discriminative performance with an accuracy of 68.4% (Figure 2b) (Table 2). The identified clustering solution deviated from the null hypothesis that clusters were generated from one single Gaussian distribution (p=0.035). Density plot of the log probabilities estimated by the GMM model showed that, although the subtypes distributions are partially overlapping, they reflected two distinct Gaussian distributions (Figure 2A). Similar results were obtained with UMAP dimensionality reduction (Table 2, Figure S1A and S1B).

**Figure 1.**
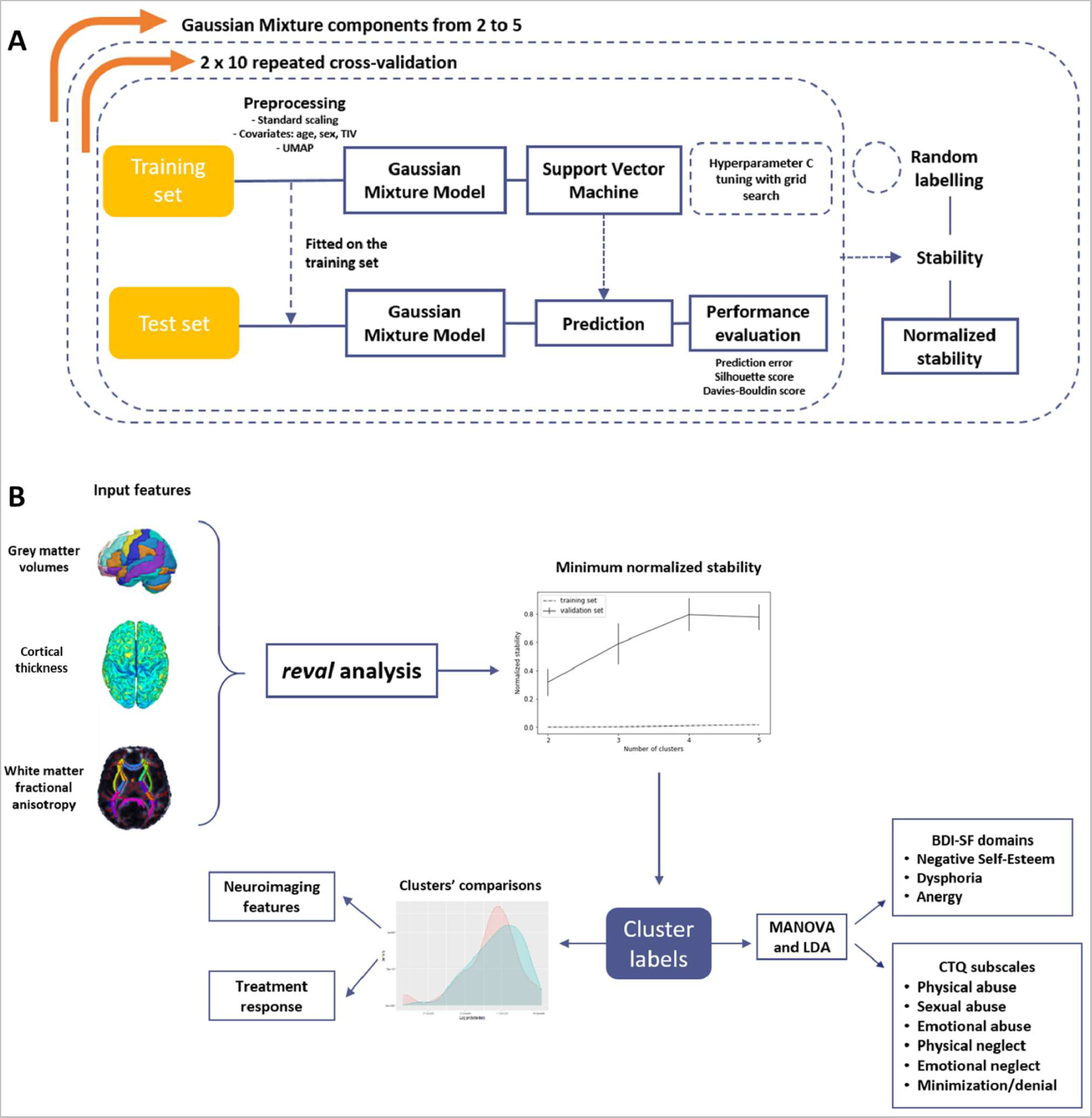
Schematic representation of the statistical analysis plan. A) *reval* algorithm pipeline. B) analysis pipeline for applying *reval* on neuroimaging data. Clusters’ labels derived from the best clustering solution (i.e., minimum normalized stability) were used for MANOVA and clusters’ comparisons on neuroimaging features and treatment response. Adapted from (38).

**Figure 2.**
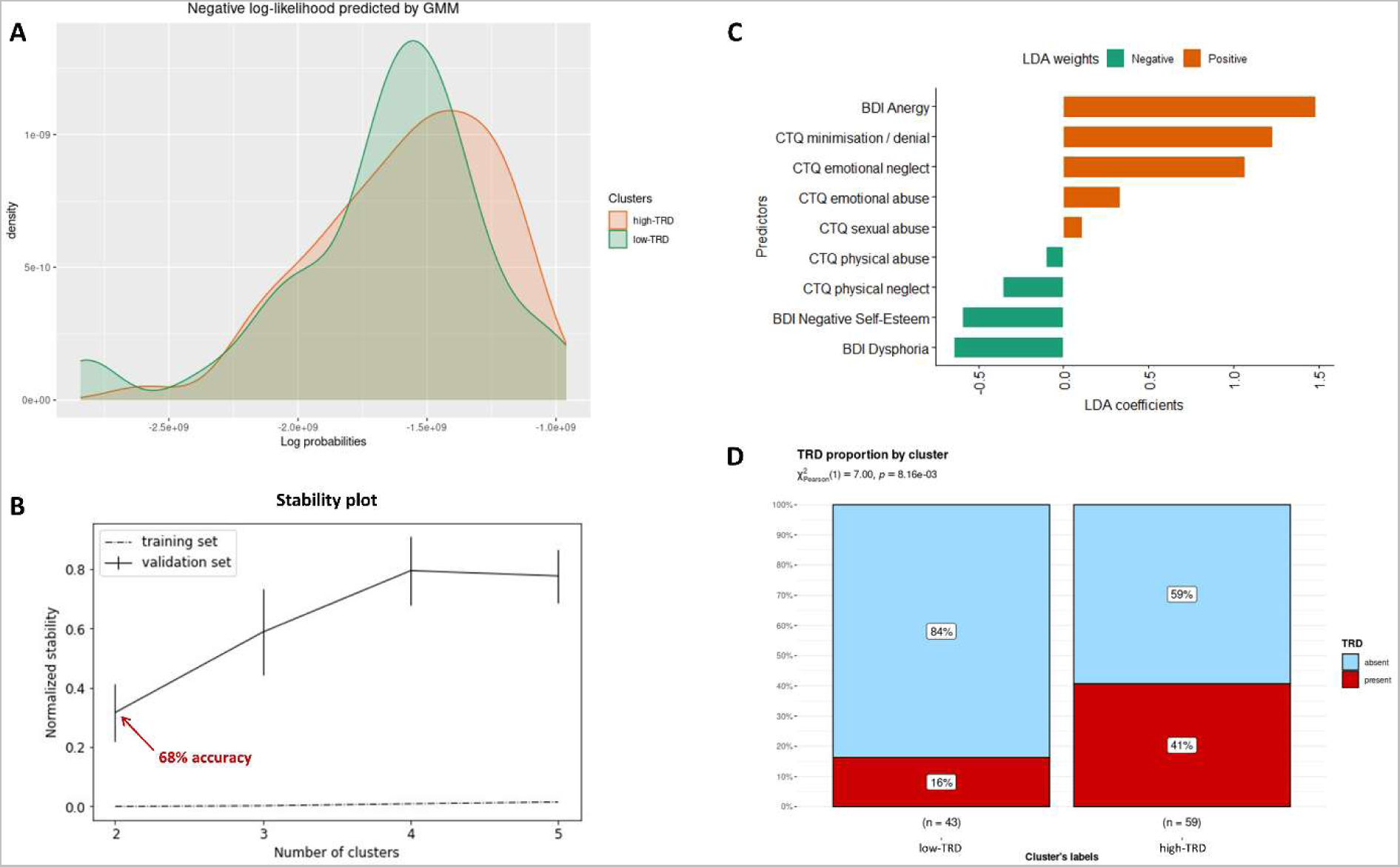
Unsupervised data-driven identification of depression subtypes based on neuroimaging data. This figure represents results from the stability-based relative clustering approach. A) Probability density distributions identified by Gaussian Mixture Model and Support Vector Machine without UMAP data reduction. B) Normalized stability plot. Error bars represent the 95% confidence interval for normalized stability from the 10×2 repeated cross-validation. The optimal number of clusters that minimizes the normalized stability was 2, achieving an accuracy of 68%. C) Standardized coefficients of the linear discriminant analysis considering clusters’ labels as fixed factors and BDI and CTQ domains as dependent variables. Positive weights (orange) reflect a higher probability to be assigned to the high-TRD cluster, whereas negative weights (green) indicate a higher probability to belong to the low-TRD cluster. D) Graphical representation of the proportion of TRD patients between the identified clusters.

**Table 1.** Descriptive statistics of the whole sample.

| Variable | mean (SD) |
| --- | --- |
| Age | 49.43 (10.00) |
| Sex | 33 M, 69 F |
| scanner | 46 old, 56 new* |
| number of episodes | 5.67 (6.21) |
| age of onset | 32.30 (11.78) |
| duration of illness | 17.13 (10.51) |
| Education years | 12.75 (3.82) |
| medication load | 4.57 (2.03) |
| BMI | 24.82 (5.27) |
| TRD | 71 non-TRD, 31 TRD |
| HDRS-21 total score | 21.03 (5.91) |

Abbreviations: SD, standard deviation; BMI, body mass index; TRD, treatment-resistant depression; HDRS, Hamilton Depression Rating Scale. \* T1-weighted and diffusion tensor images were acquired on two 3.0 Tesla scanners. Until June 2016, 46 MDD patients were acquired with a Gyroscan Intera, Philips (old scanner). From October 2016, 56 MDD patients were examined with the Ingenia CX, Philips (new scanner).

**Table 2.**
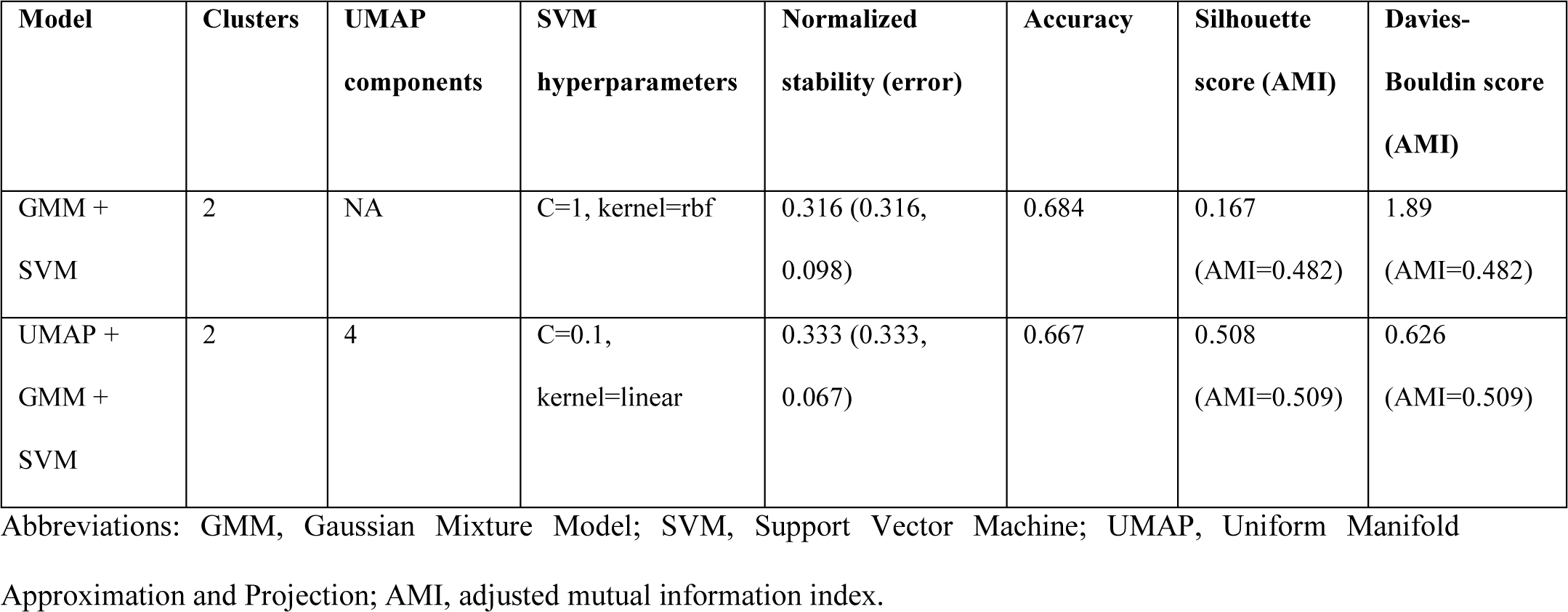
Performances and hyper-parameters for the best clustering solutions.

| Model | Clusters | UMAP components | SVM hyperparameters | Normalized stability (error) | Accuracy | Silhouette score (AMI) | Davies-Bouldin score (AMI) |
| --- | --- | --- | --- | --- | --- | --- | --- |
| GMM + SVM | 2 | NA | C=1, kernel=rbf | 0.316 (0.316, 0.098) | 0.684 | 0.167 (AMI=0.482) | 1.89 (AMI=0.482) |
| UMAP + GMM + SVM | 2 | 4 | C=0.1, kernel=linear | 0.333 (0.333, 0.067) | 0.667 | 0.508 (AMI=0.509) | 0.626 (AMI=0.509) |

Abbreviations: GMM, Gaussian Mixture Model; SVM, Support Vector Machine; UMAP, Uniform Manifold Approximation and Projection; AMI, adjusted mutual information index.

### Neurobiological-driven clusters are related to treatment response and specific neurobiological profiles

The identified clusters without UMAP data reduction were significantly different for TRD (p=0.008), sex (p=0.03), and education (p=0.016). Cluster 1 (N=43) was mainly composed by treatment-responsive patients whereas Cluster 2 (N=59) was characterised by higher TRD rates (Figure 2d). Notably, a higher F:M ratio was observed in Cluster 2 compared to Cluster 1. No significant differences in age, number of episodes, age of onset, duration of illness, and BMI was found between the two clusters (Table 3). Cortical thickness in temporo-parietal structures was significantly higher in Cluster 1 compared to Cluster 2 with large effect (p_FDR_< 0.001) (Figure 3A). Considering gray matter volumes, the clusters were different with large effects in the bilateral thalami and parahippocampal gyrus, right middle occipital gyrus, right superior occipital gyrus, left temporal pole, right precuneus, right postcentral gyrus, left medial frontal cortex, left gyrus rectus, left precentral gyrus, and left superior parietal lobule, with higher volumes in Cluster 1 compared to Cluster 2 (p_FDR_ < 0.001)) (Figure 3b). As for white matter, higher FA values were found in Cluster 1 compared to Cluster 2 (Figure 3C). Specifically, the right superior fronto-occipital fasciculus (p_FDR_ = 0.032), the body of the corpus callosum (p_FDR_ = 0.024), and the right stria terminalis (p_FDR_ = 0.037) differentiated the two clusters with a moderate effect. (Table S1). A similar clustering solution was identified in the UMAP-reduced model (Results S1).

**Figure 3.**
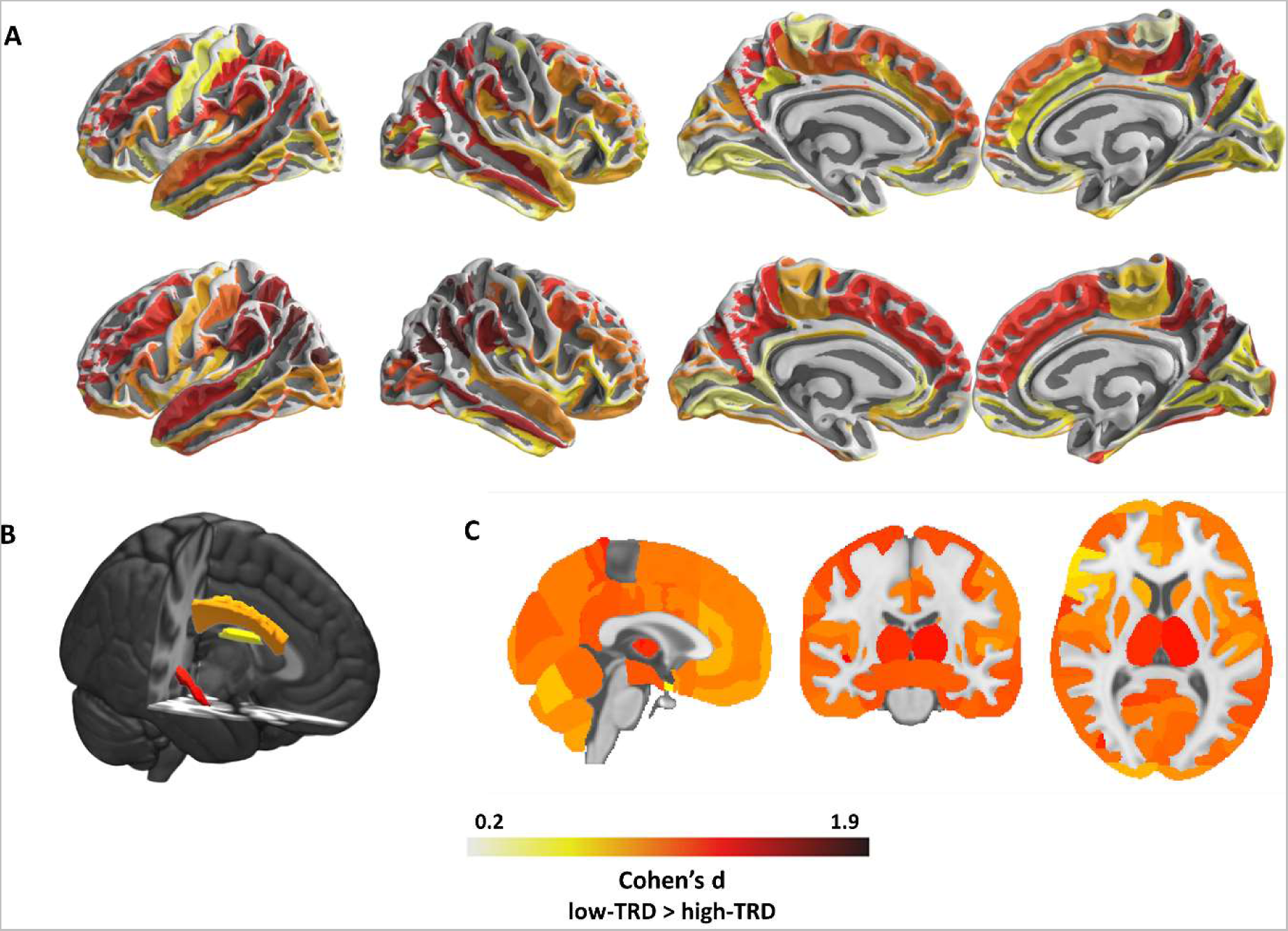
Standardized effect sizes differences (Cohen’s d) in cortical, subcortical, and white matter structures associated with the identified clusters. Increasing red color indicates higher effect sizes (low-TRD>high-TRD). A) Effect sizes for cortical thickness based on Destrieux (top) and Desikan-Killiany (down) atlases. B) Effect sizes for extracted tract-based FA values. C) Effect sizes for extracted grey matter volumes.

**Table 3.** Demographic and clinical characteristics of the identified data-driven clusters without UMAP dimensionality reduction.

| Variable | Cluster 1 (N=43) | Cluster 2 (N=59) | t / $\chi^2$ | p |
| --- | --- | --- | --- | --- |
| age | 50.98 (8.73) | 48.31 (10.77) | 1.34 | 0.184 |
| sex | 19 M, 24 F | 14 M, 45 F | 4.76 | 0.03* |
| scanner | 19 old, 24 new | 27 old, 32 new | 0.03 | 0.874 |
| number of episodes | 5.47 (5.15) | 5.81 (6.91) | -0.28 | 0.781 |
| age of onset | 33.93 (11.18) | 31.12 (12.15) | 1.19 | 0.236 |
| duration of illness | 17.05 (11.05) | 17.19 (10.19) | -0.07 | 0.947 |
| Education years | 11.70 (3.80) | 13.53 (3.68) | -2.44 | 0.016* |
| medication load | 4.35 (2.13) | 4.71 (2.14) | -0.85 | 0.398 |
| BMI | 25.29 (4.63) | 24.99 (4.76) | 0.32 | 0.375 |
| TRD | 36 non-TRD, 7 TRD | 35 non-TRD, 24 TRD | 7.00 | 0.008** |
| HDRS-21 total score | 23.19 (7.26) | 22.27 (7.00) | 0.64 | 0.522 |
| CTQ total score<sup>a</sup> | 44.92 (15.04) | 41.40 (13.41) | 0.94 | 0.351 |
| CTQ physical abuse<sup>a</sup> | 6.25 (2.47) | 6.13 (2.46) | 0.20 | 0.845 |
| CTQ emotional neglect<sup>a</sup> | 13.83 (5.54) | 12.90 (5.27) | 0.67 | 0.504 |
| CTQ emotional abuse<sup>a</sup> | 9.38 (4.54) | 8.93 (4.31) | 0.40 | 0.693 |
| CTQ physical neglect<sup>a</sup> | 8.54 (4.03) | 7.03 (3.04) | 1.71 | 0.093 |
| CTQ sexual abuse<sup>a</sup> | 6.08 (3.51) | 6.18 (3.38) | -0.10 | 0.918 |
| CTQ minimization/denial<sup>a</sup> | 7.92 (3.13) | 9.40 (3.05) | -1.86 | 0.067 |
| BDI total score<sup>a</sup> | 16.13 (8.96) | 16.73 (7.74) | -0.28 | 0.778 |
| BDI Self-Esteem<sup>a</sup> | 5.92 (3.48) | 5.80 (3.08) | 0.14 | 0.889 |
| BDI Anergy<sup>a</sup> | 4.79 (3.02) | 6.15 (2.53) | -1.93 | 0.058 |
| BDI Dysphoria<sup>a</sup> | 5.42 (3.62) | 5.20 (2.70) | 0.27 | 0.784 |

Results for continuous variables are reported as mean (standard deviation), whereas sample size is reported for categorical variables. Abbreviations: BMI, body mass index; TRD, treatment-resistant depression; HDRS-21, 21-Hamilton Depression Rating Scale; CTQ, Childhood Trauma Questionnaire; BDI, Beck Depression Inventory.
<sup>a</sup> subsample of 64 MDD patients with clinical scales (cluster 1: N=24; cluster 2: N=40). \*p<0.05, \*\*p<0.001

### Neuroimaging-based subtypes reflect specific childhood trauma and depression-related signatures

The identified data-driven clusters reflected subtypes of depressed patients based on childhood trauma and depressive symptomatology using a multivariate approach. MANOVA results showed a significant between-clusters difference (Wilks’ lambda=0.68, F(9,50)=2.64, p=0.014), considering age, sex, medication load, and number of episodes as nuisance covariates. Results from univariate statistics were not significant (Table 3). The following LDA identified one discriminant function (variate) that significantly differentiated the two clusters (Wilks’ lambda=0.70, p=0.013). Standardised discriminant coefficients revealed that BDI Anergy (b=1.48), CTQ minimisation/denial (b=1.23), and CTQ emotional neglect (b=1.07) were largely positively associated with the variate, whereas Dysphoria (b = −0.65), Negative Self-Esteem (b= −0.59), and CTQ physical neglect (b = −0.36) showed a negative relationship with the variate. These results suggest that higher scores on the BDI Anergy, CTQ minimisation/denial and emotional neglect domains indicate a higher probability to be assigned to Cluster 2 Conversely, patients with higher scores on the BDI Dysphoria, Negative Self-Esteem, and CTQ physical neglect are more likely to belong to Cluster 1 (Figure 2C). Similar findings were also obtained using clusters’ labels derived from the UMAP-reduced model (Results S1).

## Discussion

By implementing a novel cross-validated clustering procedure, we identified two subgroups of patients driven from grey matter and DTI that were differentially associated with treatment resistance, depressive symptomatology and childhood trauma. In particular, one cluster showed a mixed profile with a higher proportion of treatment-resistant patients (high-TRD), higher F:M sex ratio, and more likely to be associated with energy-related depressive symptomatology and history of childhood emotional neglect and abuse. A second cluster was largely characterised by treatment-responsive patients (low-TRD), with higher cognitive and affective symptoms of depression. In terms of neuroimaging profiles, the high-TRD cluster displayed reduced widespread cortical thickness and volumes compared to the low-TRD one, along with lower white matter integrity. The identified clusters were differentiable with an accuracy of 67%, suggesting that structural neuroimaging features can enhance the discovery of clinically-meaningful subtypes of depression and treatment resistant patients.

Considering the clinical signatures of the identified subgroups, the BDI Anergy domain contributed the most in differentiating the two groups, suggesting a positive relationship with the high-TRD cluster and symptoms related to fatigue, decreased appetite, and work difficulties, especially in females. Previous studies showed that symptoms reflecting an altered energy intake/output balance (e.g., appetite changes, sleep disturbances, and fatigue), characterize a specific MDD subtype characterised by altered immune-metabolic functions (55, 56). In line with our stratification model, convergent evidence indicates that this symptomatology is more frequent in females with an early onset of diseases, recurrent episodes, and poor treatment outcomes (57). Also CTQ minimization/denial, emotional abuse and neglect scores contributed to discrimination between the two subtypes, reflecting a higher probability to belong to the high-TRD subtype. Previous evidence showed a possible link between childhood trauma and neurovegetative depressive symptomatology, reporting higher exposure to traumatic events in MDD patients with atypical features compared to those without (58). Indeed, it is possible that the high-TRD cluster identifies a specific MDD subgroup with a higher sensitivity to early stress and more current anergic symptoms, in analogy to the extensive research documenting higher prevalence of anergic/atypical symptoms in reactive depression, both categories being less responsive to monoamine reuptake inhibitors (59, 60). This symptom profile and childhood trauma may also suggest a subthreshold bipolarity, usually associated with TRD (61).

The data-driven subgroups also displayed distinct grey matter and structural connectivity profiles. Volumetric reductions in the precuneus, parahipppocampal gyrus, thalamus, temporal pole, and pre- and post-central gyri, were found in the high-TRD subtype. Most of these structures represent core areas in cortico-striatal-thalamic circuits, which are involved in attribution of salience to external stimuli, emotional regulation of inner states, and cognition (62, 63). Aberrant function and connectivity within these networks has been previously associated with treatment resistance (64, 65). Notably, a similar stratification was found based on functional neuroimaging features, with hyperconnectivity in thalamic and fronto-striatal networks associated with increased anhedonia and psychomotor retardation and less responsiveness to TMS intervention (28). Our study provides additional evidence supporting these findings, indicating that these functional abnormalities might reflect morphological alterations. When considering cortical thickness, the largest effects sizes were observed for temporo-parietal structures, with higher thickness values in the low-TRD subtype. Similar results were observed in a larger cohort study, where two MDD subgroups based on cortical thickness and surface reflected differences on general cognitive ability (33). Our results take a step forward providing evidence of an association with treatment response and other clinically meaningful measures. Together with grey matter, lower FA values in the superior fronto-occipital fasciculus, body of corpus callosum, stria terminalis, and hippocampal cingulum, were found in the high-TRD subtype. Previous studies brought evidence of decreased FA affecting these structures in TRD patients compared to healthy controls and first-episode MDD patients (66, 67), whereas microstructural changes in the cingulum and stria terminalis are predictive of remission after 8-weeks of antidepressant treatment (68). Accordingly, the preserved integrity of these structures might reflect a better cortico-limbic modulation and inter-hemispheric integration, enhancing probability of remission after treatment.

Previous studies aimed at identifying subgroups of MDD patients using neuroimaging data, showing how the clinical heterogeneity of MDD can be disentangled based on shared neurobiological correlates (27–29, 69). By exploring the relationships with childhood trauma and treatment response, we demonstrated that the identified biologically-driven subtypes are informative about relevant clinical features outside depressive symptomatology, providing a more comprehensive picture of the multifaceted manifestations of MDD. A large drawback of previous studies is the application of conventional clustering approaches tied to the specific data at hand, limiting the generalizability of the identified subtypes. Contrary, the clustering pipeline implemented in our study has the advantage to translate the unsupervised setting into a supervised classification problem, providing a measure of clusters’ stability and replicability to unseen data through cross-validation. Furthermore, a high-value strength of this work is the clinical validation of the discovered clusters, which can potentially provide insights for new effective treatments tailored on the patient’s biological and clinical profile. For instance, patients assigned to the high-TRD cluster might benefit from antidepressant therapies inducing neuroplastic changes, such as electroconvulsive therapy (70–73), repetitive transcranial magnetic stimulation (74, 75), deep brain stimulation (76, 77), and ketamine (78–80), whereas treatments targeting the immune-metabolic pathways, such as physical exercise, diet or anti-inflammatory treatments, may be also useful in alleviating the energy-related symptoms (81–83). Furthermore, considering that the high-TRD cluster was also associated with high scores for CTQ, this group may benefit from adjunctive psychotherapeutic interventions focused on trauma (84, 85). On the other hand, psychotherapies targeting cognitive distortions, such as cognitive behavioural therapy, might represent optimal treatment options for patients assigned to the low-TRD group (86–88).

The limitations of the current study should be acknowledged. First, all patients were recruited in the same clinical center, limiting the possibility to extend our findings to other cohorts. However, the cross-validation framework provides a good approximation of clusters’ generalizability (38). Second, since all patients were under pharmacological treatment at the time of scanning, we cannot rule out long-lasting effects of drugs on neuroimaging features. Another limitation comes from the sample size, which further may limit results generalizability. Our stratification model should be evaluated in larger cohorts to provide realistic applications in clinical practice. Finally, we recognize that noise in biological data might have affected clustering solutions. It should be considered, though, that the pre-processing steps employed (e.g., ComBat harmonization) helped in mitigating this issue.

In conclusion, our results indicate that structural neuroimaging data can be used to define novel subtypes of depression that are informative about treatment resistance, depressive symptomatology, and childhood trauma. A multimodal stratification including other kind of data (e.g., functional neuroimaging, genetic data, and immune-inflammatory markers) may improve the prediction of disease’s outcomes, uncovering how different pathophysiological mechanisms interact with different clinical features of depression.

## Data Availability

All data produced in the present study are available upon reasonable request to the authors.

## Acknowledgments

This study was supported by the Italian Ministry of Health (grant numbers GR 2019-12370616 and PNRR-MAD-2022-12375859).

## References

1. Kessler RC, Birnbaum H, Bromet E, Hwang I, Sampson N, Shahly V (2010): Age differences in major depression: results from the National Comorbidity Survey Replication (NCS-R). Psychological medicine. 40:225–237.

2. Conway CR, George MS, Sackeim HA (2017): Toward an evidence-based, operational definition of treatment-resistant depression: when enough is enough. JAMA psychiatry. 74:9–10.

3. Sforzini L, Worrell C, Kose M, Anderson IM, Aouizerate B, Arolt V, et al. (2022): A Delphi-method-based consensus guideline for definition of treatment-resistant depression for clinical trials. Molecular psychiatry. 27:1286–1299.

4. Johnston KM, Powell LC, Anderson IM, Szabo S, Cline S (2019): The burden of treatment-resistant depression: a systematic review of the economic and quality of life literature. Journal of affective disorders. 242:195–210.

5. Fekadu A, Wooderson SC, Markopoulo K, Donaldson C, Papadopoulos A, Cleare AJ (2009): What happens to patients with treatment-resistant depression? A systematic review of medium to long term outcome studies. Journal of affective disorders. 116:4–11.

6. Rush AJ, Trivedi MH, Wisniewski SR, Nierenberg AA, Stewart JW, Warden D, et al. (2006): Acute and longer-term outcomes in depressed outpatients requiring one or several treatment steps: a STAR*D report. The American journal of psychiatry. 163:1905–1917.

7. Thase ME, Rush AJ (1997): When at first you don’t succeed: sequential strategies for antidepressant nonresponders. Journal of Clinical Psychiatry. 58:23–29.

8. American Psychiatric Association D-TF (2013): Diagnostic and statistical manual of mental disorders: DSM-5™, 5th ed. Arlington, VA, US: American Psychiatric Publishing, Inc.

9. Fagiolini A, Kupfer DJ (2003): Is treatment-resistant depression a unique subtype of depression? Biological Psychiatry. 53:640–648.

10. Geschwind DH, Flint J (2015): Genetics and genomics of psychiatric disease. Science. 349:1489–1494.

11. Sullivan PF, Neale MC, Kendler KS (2000): Genetic epidemiology of major depression: review and meta-analysis. American journal of psychiatry. 157:1552–1562.

12. Williams LM, Debattista C, Duchemin AM, Schatzberg AF, Nemeroff CB (2016): Childhood trauma predicts antidepressant response in adults with major depression: data from the randomized international study to predict optimized treatment for depression. Transl Psychiatry. 6:e799.

13. Douglas KM, Porter RJ (2012): The effect of childhood trauma on pharmacological treatment response in depressed inpatients. Psychiatry Res. 200:1058–1061.

14. Chekroud AM, Zotti RJ, Shehzad Z, Gueorguieva R, Johnson MK, Trivedi MH, et al. (2016): Cross-trial prediction of treatment outcome in depression: a machine learning approach. The lancet Psychiatry. 3:243–250.

15. Teicher MH, Gordon JB, Nemeroff CB (2022): Recognizing the importance of childhood maltreatment as a critical factor in psychiatric diagnoses, treatment, research, prevention, and education. Molecular psychiatry. 27:1331–1338.

16. Nanni V, Uher R, Danese A (2012): Childhood maltreatment predicts unfavorable course of illness and treatment outcome in depression: a meta-analysis. American Journal of Psychiatry. 169:141–151.

17. Klok MPC, van Eijndhoven PF, Argyelan M, Schene AH, Tendolkar I (2019): Structural brain characteristics in treatment-resistant depression: review of magnetic resonance imaging studies. BJPsych Open. 5:e76.

18. Runia N, Yücel DE, Lok A, de Jong K, Denys D, van Wingen GA, et al. (2022): The neurobiology of treatment-resistant depression: A systematic review of neuroimaging studies. Neuroscience and biobehavioral reviews. 132:433–448.

19. Miola A, Meda N (2023): Structural and functional features of treatment-resistant depression: A systematic review and exploratory coordinate-based meta-analysis of neuroimaging studies.

20. Pelin H, Ising M, Stein F, Meinert S, Meller T, Brosch K, et al. (2021): Identification of transdiagnostic psychiatric disorder subtypes using unsupervised learning. Neuropsychopharmacology. 46:1895–1905.

21. Dinga R, Marquand AF, Veltman DJ, Beekman AT, Schoevers RA, van Hemert AM, et al. (2018): Predicting the naturalistic course of depression from a wide range of clinical, psychological, and biological data: a machine learning approach. Translational psychiatry. 8:241.

22. Wen J, Fu CH, Tosun D, Veturi Y, Yang Z, Abdulkadir A, et al. (2022): Characterizing heterogeneity in neuroimaging, cognition, clinical symptoms, and genetics among patients with late-life depression. JAMA psychiatry. 79:464–474.

23. Chekroud AM, Gueorguieva R, Krumholz HM, Trivedi MH, Krystal JH, McCarthy G (2017): Reevaluating the efficacy and predictability of antidepressant treatments: a symptom clustering approach. JAMA psychiatry. 74:370–378.

24. Kaster TS, Downar J, Vila-Rodriguez F, Baribeau DA, Thorpe KE, Daskalakis ZJ, et al. (2023): Differential symptom cluster responses to repetitive transcranial magnetic stimulation treatment in depression. Eclinicalmedicine. 55:101765.

25. Rost N, Dwyer DB, Gaffron S, Rechberger S, Maier D, Binder EB, et al. (2023): Multimodal predictions of treatment outcome in major depression: A comparison of data-driven predictors with importance ratings by clinicians. Journal of Affective Disorders.

26. Lorenzo-Luaces L, Buss JF, Fried EI (2021): Heterogeneity in major depression and its melancholic and atypical specifiers: a secondary analysis of STAR* D. BMC psychiatry. 21:1–11.

27. Beijers L, Wardenaar KJ, van Loo HM, Schoevers RA (2019): Data-driven biological subtypes of depression: systematic review of biological approaches to depression subtyping. Molecular psychiatry. 24:888–900.

28. Drysdale AT, Grosenick L, Downar J, Dunlop K, Mansouri F, Meng Y, et al. (2017): Resting-state connectivity biomarkers define neurophysiological subtypes of depression. Nature medicine. 23:28–38.

29. Tokuda T, Yoshimoto J, Shimizu Y, Okada G, Takamura M, Okamoto Y, et al. (2018): Identification of depression subtypes and relevant brain regions using a data-driven approach. Scientific reports. 8:14082.

30. Wang Y, Tang S, Zhang L, Bu X, Lu L, Li H, et al. (2021): Data-driven clustering differentiates subtypes of major depressive disorder with distinct brain connectivity and symptom features. The British Journal of Psychiatry. 219:606–613.

31. Liang S, Deng W, Li X, Greenshaw AJ, Wang Q, Li M, et al. (2020): Biotypes of major depressive disorder: neuroimaging evidence from resting-state default mode network patterns. NeuroImage: Clinical. 28:102514.

32. Zhou Y, Zhao L, Zhou N, Zhao Y, Marino S, Wang T, et al. (2019): Predictive big data analytics using the UK biobank data. Scientific reports. 9:1–10.

33. Yeung HW, Shen X, Stolicyn A, De Nooij L, Harris MA, Romaniuk L, et al. (2021): Spectral clustering based on structural magnetic resonance imaging and its relationship with major depressive disorder and cognitive ability. European Journal of Neuroscience. 54:6281–6303.

34. Buch AM, Liston C (2021): Dissecting diagnostic heterogeneity in depression by integrating neuroimaging and genetics. Neuropsychopharmacology. 46:156–175.

35. Dinga R, Schmaal L, Penninx BW, van Tol MJ, Veltman DJ, van Velzen L, et al. (2019): Evaluating the evidence for biotypes of depression: Methodological replication and extension of. NeuroImage: Clinical. 22:101796.

36. Feczko E, Miranda-Dominguez O, Marr M, Graham AM, Nigg JT, Fair DA (2019): The heterogeneity problem: Approaches to identify psychiatric subtypes. Trends in Cognitive Sciences. 23:584–601.

37. Nunes A, Trappenberg T, Alda M (2020): The definition and measurement of heterogeneity. Translational psychiatry. 10:299.

38. Landi I, Mandelli V, Lombardo MV (2021): reval: A Python package to determine best clustering solutions with stability-based relative clustering validation. Patterns. 2:100228.

39. Sackeim HA (2001): The definition and meaning of treatment-resistant depression. Journal of Clinical Psychiatry. 62:10–17.

40. Leckman JF, Sholomskas D, Thompson D, Belanger A, Weissman MM (1982): Best estimate of lifetime psychiatric diagnosis: a methodological study. Archives of general psychiatry. 39:879–883.

41. Reynolds WM, Gould JW (1981): A psychometric investigation of the standard and short form Beck Depression Inventory. Journal of consulting and clinical psychology. 49:306.

42. Foelker Jr GA, Shewchuk RM, Niederehe G (1987): Confirmatory factor analysis of the short form Beck Depression Inventory in elderly community samples. Journal of Clinical Psychology. 43:111–118.

43. Bernstein DP, Stein JA, Newcomb MD, Walker E, Pogge D, Ahluvalia T, et al. (2003): Development and validation of a brief screening version of the Childhood Trauma Questionnaire. Child abuse & neglect. 27:169–190.

44. Berstein DP, Fink L (1998): Childhood trauma questionnaire: A retrospective self-report. The Psychological Corporation: San Antonio.

45. Gaser C, Dahnke R (2016): CAT-a computational anatomy toolbox for the analysis of structural MRI data. Hbm. 2016:336–348.

46. Desikan RS, Ségonne F, Fischl B, Quinn BT, Dickerson BC, Blacker D, et al. (2006): An automated labeling system for subdividing the human cerebral cortex on MRI scans into gyral based regions of interest. Neuroimage. 31:968–980.

47. Destrieux C, Fischl B, Dale A, Halgren E (2010): Automatic parcellation of human cortical gyri and sulci using standard anatomical nomenclature. Neuroimage. 53:1–15.

48. Horsfield MA (1999): Mapping eddy current induced fields for the correction of diffusion-weighted echo planar images. Magnetic resonance imaging. 17:1335–1345.

49. Smith SM (2002): Fast robust automated brain extraction. Human brain mapping. 17:143–155.

50. Behrens TE, Woolrich MW, Jenkinson M, Johansen-Berg H, Nunes RG, Clare S, et al. (2003): Characterization and propagation of uncertainty in diffusion-weighted MR imaging. Magnetic Resonance in Medicine: An Official Journal of the International Society for Magnetic Resonance in Medicine. 50:1077–1088.

51. Smith SM, Jenkinson M, Johansen-Berg H, Rueckert D, Nichols TE, Mackay CE, et al. (2006): Tract-based spatial statistics: voxelwise analysis of multi-subject diffusion data. Neuroimage. 31:1487–1505.

52. Mori S, Oishi K, Jiang H, Jiang L, Li X, Akhter K, et al. (2008): Stereotaxic white matter atlas based on diffusion tensor imaging in an ICBM template. Neuroimage. 40:570–582.

53. Fortin J-P, Parker D, Tunç B, Watanabe T, Elliott MA, Ruparel K, et al. (2017): Harmonization of multi-site diffusion tensor imaging data. NeuroImage. 161:149–170.

54. Huang H, Liu Y, Yuan M, Marron J (2015): Statistical significance of clustering using soft thresholding. Journal of Computational and Graphical Statistics. 24:975–993.

55. Lamers F, Milaneschi Y, De Jonge P, Giltay E, Penninx B (2018): Metabolic and inflammatory markers: associations with individual depressive symptoms. Psychological medicine. 48:1102–1110.

56. Milaneschi Y, Lamers F, Berk M, Penninx BW (2020): Depression heterogeneity and its biological underpinnings: toward immunometabolic depression. Biological Psychiatry. 88:369–380.

57. Vreijling SR, van Haeringen M, Milaneschi Y, Huider F, Bot M, Amin N, et al. (2023): Sociodemographic, lifestyle and clinical characteristics of energy-related depression symptoms: A pooled analysis of 13,965 depressed cases in 8 Dutch cohorts. Journal of Affective Disorders. 323:1–9.

58. Withers AC, Tarasoff JM, Stewart JW (2013): Is depression with atypical features associated with trauma history? J Clin Psychiatry. 74:500–506.

59. Quitkin FM, Stewart JW, Mcgrath PJ, Tricamo E, Rabkin JG, Ocepek-Welikson K, et al. (1993): Columbia atypical depression: a subgroup of depressives with better response to MAOI than to tricyclic antidepressants or placebo. The British Journal of Psychiatry. 163:30–34.

60. Stewart JW (2007): Treating depression with atypical features. Journal of clinical psychiatry. 68:25–29.

61. Olgiati P, Serretti A (2022): Post-traumatic stress disorder and childhood emotional abuse are markers of subthreshold bipolarity and worse treatment outcome in major depressive disorder. International clinical psychopharmacology. 37:1.

62. Bora E, Harrison BJ, Davey CG, Yücel M, Pantelis C (2012): Meta-analysis of volumetric abnormalities in cortico-striatal-pallidal-thalamic circuits in major depressive disorder. Psychological medicine. 42:671–681.

63. Price JL, Drevets WC (2012): Neural circuits underlying the pathophysiology of mood disorders. Trends in cognitive sciences. 16:61–71.

64. Lui S, Wu Q, Qiu L, Yang X, Kuang W, Chan RC, et al. (2011): Resting-state functional connectivity in treatment-resistant depression. American Journal of Psychiatry. 168:642–648.

65. Yamamura T, Okamoto Y, Okada G, Takaishi Y, Takamura M, Mantani A, et al. (2016): Association of thalamic hyperactivity with treatment-resistant depression and poor response in early treatment for major depression: a resting-state fMRI study using fractional amplitude of low-frequency fluctuations. Translational psychiatry. 6:e754–e754.

66. de Diego-Adelino J, Pires P, Gómez-Ansón B, Serra-Blasco M, Vives-Gilabert Y, Puigdemont D, et al. (2014): Microstructural white-matter abnormalities associated with treatment resistance, severity and duration of illness in major depression. Psychological medicine. 44:1171–1182.

67. Guo W-b, Liu F, Chen J-d, Xu X-j, Wu R-r, Ma C-q, et al. (2012): Altered white matter integrity of forebrain in treatment-resistant depression: a diffusion tensor imaging study with tract-based spatial statistics. Progress in Neuro-Psychopharmacology and Biological Psychiatry. 38:201–206.

68. Korgaonkar MS, Williams LM, Song YJ, Usherwood T, Grieve SM (2014): Diffusion tensor imaging predictors of treatment outcomes in major depressive disorder. The British Journal of Psychiatry. 205:321–328.

69. Price RB, Gates K, Kraynak TE, Thase ME, Siegle GJ (2017): Data-driven subgroups in depression derived from directed functional connectivity paths at rest. Neuropsychopharmacology. 42:2623–2632.

70. Bracht T, Walther S, Breit S, Mertse N, Federspiel A, Meyer A, et al. (2023): Distinct and shared patterns of brain plasticity during electroconvulsive therapy and treatment as usual in depression: an observational multimodal MRI-study. Translational Psychiatry. 13:6.

71. Sartorius A, Demirakca T, Böhringer A, von Hohenberg CC, Aksay SS, Bumb JM, et al. (2016): Electroconvulsive therapy increases temporal gray matter volume and cortical thickness. European Neuropsychopharmacology. 26:506–517.

72. Schmitgen MM, Kubera KM, Depping MS, Nolte HM, Hirjak D, Hofer S, et al. (2020): Exploring cortical predictors of clinical response to electroconvulsive therapy in major depression. European archives of psychiatry and clinical neuroscience. 270:253–261.

73. Yrondi A, Nemmi F, Billoux S, Giron A, Sporer M, Taib S, et al. (2019): Grey Matter changes in treatment-resistant depression during electroconvulsive therapy. Journal of Affective Disorders. 258:42–49.

74. Jannati A, Oberman LM, Rotenberg A, Pascual-Leone A (2023): Assessing the mechanisms of brain plasticity by transcranial magnetic stimulation. Neuropsychopharmacology. 48:191–208.

75. Ge R, Humaira A, Gregory E, Alamian G, MacMillan EL, Barlow L, et al. (2022): Predictive value of acute neuroplastic response to rTMS in treatment outcome in depression: a concurrent TMS-fMRI trial. American Journal of Psychiatry. 179:500–508.

76. Cattarinussi G, Moghaddam HS, Aarabi MH, Squarcina L, Sambataro F, Brambilla P, et al. (2022): White Matter Microstructure Associated with the Antidepressant Effects of Deep Brain Stimulation in Treatment-Resistant Depression: A Review of Diffusion Tensor Imaging Studies. International Journal of Molecular Sciences. 23:15379.

77. Conner CR, Quevedo J, Soares JC, Fenoy AJ (2022): Brain metabolic changes and clinical response to superolateral medial forebrain bundle deep brain stimulation for treatment-resistant depression. Molecular Psychiatry.1–7.

78. Dai D, Lacadie CM, Holmes SE, Cool R, Anticevic A, Averill C, et al. (2020): Ketamine normalizes the structural alterations of inferior frontal gyrus in depression. Chronic Stress. 4:2470547020980681.

79. Taraku B, Woods RP, Boucher M, Espinoza R, Jog M, Al-Sharif N, et al. (2023): Changes in white matter microstructure following serial ketamine infusions in treatment resistant depression. Human Brain Mapping. 44:2395–2406.

80. Kopelman J, Keller TA, Panny B, Griffo A, Degutis M, Spotts C, et al. (2023): Rapid neuroplasticity changes and response to intravenous ketamine: a randomized controlled trial in treatment-resistant depression. Translational Psychiatry. 13:1–9.

81. Rethorst CD, Tu J, Carmody TJ, Greer TL, Trivedi MH (2016): Atypical depressive symptoms as a predictor of treatment response to exercise in Major Depressive Disorder. Journal of affective disorders. 200:156–158.

82. Vreijling SR, Penninx BW, Bot M, Watkins E, Owens M, Kohls E, et al. (2022): Effects of dietary interventions on depressive symptom profiles: results from the MooDFOOD depression prevention study. Psychological Medicine. 52:3580–3589.

83. Hang X, Zhang Y, Li J, Li Z, Zhang Y, Ye X, et al. (2021): Comparative efficacy and acceptability of anti-inflammatory agents on major depressive disorder: a network meta-analysis. Frontiers in Pharmacology. 12:691200.

84. Mannarino AP, Cohen JA, Deblinger E (2014): Trauma-focused cognitive-behavioral therapy. Evidence-based approaches for the treatment of maltreated children: Considering core components and treatment effectiveness.165–185.

85. Shapiro F (2001): Eye movement desensitization and reprocessing (EMDR): Basic principles, protocols, and procedures. Guilford Press.

86. Wiles N, Thomas L, Abel A, Ridgway N, Turner N, Campbell J, et al. (2013): Cognitive behavioural therapy as an adjunct to pharmacotherapy for primary care based patients with treatment resistant depression: results of the CoBalT randomised controlled trial. The Lancet. 381:375–384.

87. Li JM, Zhang Y, Su WJ, Liu LL, Gong H, Peng W, et al. (2018): Cognitive behavioral therapy for treatment-resistant depression: A systematic review and meta-analysis. Psychiatry research. 268:243–250.

88. Quigley L, Dozois DJ, Bagby RM, Lobo DS, Ravindran L, Quilty LC (2019): Cognitive change in cognitive-behavioural therapy v. pharmacotherapy for adult depression: a longitudinal mediation analysis. Psychological medicine. 49:2626–2634.

