## Supplementary material for "Multimodal structural neuroimaging data unveil data-driven subtypes of treatment-resistant depression": Methods S1

**Supplementary Materials**

**Methods S1. Inclusion and exclusion criteria**

All patients met DSM-5 criteria for MDD, according to the Structured Clinical Interview for the Diagnosis of Axis I Disorders (SCID-I). To be included in the study, patients had to be 18-65 years old and to have a major depressive episode without psychotic features at the time of scanning. Exclusion criteria for all subjects were major medical and neurological disorders, pregnancy, intellectual disability, history of drug or alcohol abuse or dependency, uncontrolled systemic disease or other significant uncontrolled somatic disorder known to affect mood. In order to reduce the impact of potential misdiagnosis with bipolar disorders and to exclude possible depressive symptoms not ascribable to MDD, only patients with at least two previous depressive episodes and a rated severity of depression higher than 8 on the 21-Hamilton Depression Rating Scale (HDRS-21) (1) were included.

**Methods S2. Rating criteria for medication load calculation**

All individual medication scores for each medication category (i.e., atypical antipsychotics, typical antipsychotics, benzodiazepines, mood stabilizers, serotonin-norepinephrine reuptake inhibitors - SNRI, selective serotonin reuptake inhibitors - SSRI, tricyclic antidepressant, anticonvulsants) were then combined in each individual participant to obtain a single composite score. Each medication category was rated from low to high dosage according to specific criteria specified by Sackeim (2). Classification was based according to the following general guidelines:

1. Any trial with duration < 4 weeks, as well as monotherapies with medications without clear efficacy for unipolar depression, should be rated as “1”, regardless of dosage.
2. For alprazolam, specific anticonvulsants, and lithium the maximum score should be “2”.
3. Lack of compliance and/or trial discontinuation reduce the overall strength of the trial.
4. In case of combined therapies, each medication should be rated independently. An exception should be made for lithium, which increases by one point the ratings of the trials if it was taken for at least 2 weeks and antidepressant score is higher than 3.

**Methods S3. MRI data acquisition details**

Until June 2016, 46 MDD patients were acquired with Gyroscan Intera, Philips, Netherlands employing a 8 channels SENSE head coil (T1-weighted MPRAGE sequences: TR 25.00 ms, TE 4.6 ms, field of view FOV=230 mm,91 matrix=256×256, in-plane resolution 0.9×0.9 mm, yielding 220 transversal slices with a thickness of 0.8 mm). For DTI, SE EPI sequences (TR/TE=9000/58 ms, FoV (mm) 232(ap), 126 (fh), 240.00 (rl); acquisition matrix = 112×85; voxel acquisition 2.14×2.73×2.3; 55 contiguous, with in-plane voxel size 1.88×1.88 mm; SENSE acceleration factor=2; 1 b0 and 35 non-collinear directions of the diffusion gradients; b value=900 s/mm2) were used. From October 2016, 56 MDD patients were examined with the new scanner 3T Ingenia CX, Philips, The Netherlands using a 32-channel sensitivity encoding SENSE head coil (T1-weighted MPRAGE sequence: TR 8.00 ms, TE 3.7 ms, field of view FOV = 256 mm, matrix = 256 x 256, in-plane resolution 1 x 1 mm, yielding 182 transversal slices with a thickness of 1 mm). For DTI, SE EPI sequences (EPI factor= 43; TR/TE=5900/78 ms, FoV (mm) 232 (ap), 129 (fh), 240.00 (rl); acquisition matrix 112×85; 56 contiguous, 2.3-mm thick axial slices reconstructed with in-plane pixel size 1.88×1.88 mm; SENSE acceleration factor= 2; Multiband acceleration factor= 2; ten b0 and 96 non-collinear directions of the diffusion gradients: 60 b values=2855 s/mm2, 6 b values=700 s/mm2, 30 b values=1000 s/mm2) were acquired. Fat saturation was performed to avoid chemical shift artefacts.

**Methods S4. Stability-based relative clustering validation**

Relative clustering validation analyses performed in our study are based on the Python package *reval* (<https://github.com/IIT-LAND/reval_clustering>). Compared to commonly used approaches that rely on the inherent characteristics of a given dataset (e.g., silhouette score), *reval* has the advantage to identify clusters that are robust and reproducible in other samples of the same population Further details about *reval* methodology are reported in (3). First, *reval* splits the original dataset into a training and test sets, and runs a clustering algorithm through a range of clusters on each set. A classification algorithm is trained on the training set to learn clusters’ labels. Then, the trained algorithm is applied on the test set to predict clusters’ labels of new observations. By comparing predictions, a performance measure called stability (i.e., prediction error) is obtained, where lower values correspond to a higher generalizability of a given clustering solution. To overcome the non-uniqueness of clustering labelling, clusters’ labels are permuted and stability is normalized for the stability of random labelling. The entire procedure is repeated through cross-validation and the number of clusters that minimized the average normalized stability is chosen. In order to be implemented on extracted neuroimaging data, we adapted reval pipeline including the following changes: 1) standardization and covariates adjustment within cross-validation; 2) combine different neuroimaging modalities and apply different set of covariates for each of them; 3) implementation of data reduction techniques (e.g., UMAP) and optimization of their parameters within cross-validation. The code to reproduce the analysis is deposited here: <https://github.com/fede-colombo/NeuReval>

**Results S1. Clustering solution derived with UMAP dimensionality reduction**

Clusters derived from the UMAP-reduced model were still significantly different for TRD (p=0.042). No significant differences for sex, education, number of episodes, age of onset, duration of illness, and BMI were found (Table S2, Figure S1D). Results of T-tests comparisons for neuroimaging features were similar to those found without data reduction. Widespread significant differences in cortical thickness were found, with higher values in parietal, supramarginal, frontal, and temporal structures in Cluster 1 compared to Cluster 2 (p_FDR_ < 0.001). Grey matter volumes in the thalami, right precentral gyrus, right supplementary motor cortex, and left medial frontal cortex were significantly higher in Cluster 1 compared to Cluster 2 with relatively large effects, whereas moderate effects in the same direction were found for frontal, parahippocampal, cingulate, and precentral cortices, as well as in the amygdalae and hippocampi (p_FDR_ < 0.001). Higher FA values in the right cingulum (hippocampal part) (p_FDR_ = 0.017) and right fornix/stria terminalis (p_FDR_ = 0.046) were found in Cluster 1 compared to Cluster 2 with moderate effects (Table S3). Considering the clinical relevance of the identified clusters, the MANOVA still detected a significant between-clusters difference (Wilks’ lambda= 0.73, F(9, 50)= 2.17, p=0.039). Univariate analyses were not significant (Table S2). The following LDA identified one discriminant function that significantly differentiated the two clusters (Wilks’ lambda=0.73, R2 canonical= 0.27, χ2(9) = 17.77, p=0.038). Standardized coefficients for the following LDA for the clusters derived from the UMAP-reduced model are reported in Figure S1C and in Table S4.

**Table S1. T-test statistics and effect sizes of the neuroimaging features driving the separation between the two clusters derived without UMAP data reduction. Results are sorted by Cohen’s d**

| **Features** | **Cluster 1** | **Cluster 2** | **T** | **FDR** | **CI 95%** | **Cohen's d** |
| --- | --- | --- | --- | --- | --- | --- |
| **Grey matter volumes** | | | | | | |
| Left Thalamus Proper | 4.734 (0.606) | 4.071 (0.646) | 5.304 | <0.001** | [0.41 0.91] | 1.053 |
| Right Middle Occipital Gyrus | 3.614 (0.548) | 3.128 (0.457) | 4.738 | <0.001** | [0.28 0.69] | 0.977 |
| Right Thalamus Proper | 4.299 (0.652) | 3.681 (0.656) | 4.714 | <0.001** | [0.36 0.88] | 0.944 |
| Left Temporal Pole | 7.401 (0.785) | 6.683 (0.757) | 4.631 | <0.001** | [0.41 1.03] | 0.934 |
| Right Superior Occipital Gyrus | 2.502 (0.434) | 2.188 (0.284) | 4.148 | <0.001** | [0.16 0.47] | 0.886 |
| Left Medial Frontal Cerebrum | 1.412 (0.213) | 1.235 (0.189) | 4.331 | <0.001** | [0.1 0.26] | 0.885 |
| Left Gyrus Rectus | 1.606 (0.213) | 1.436 (0.177) | 4.268 | <0.001** | [0.09 0.25] | 0.880 |
| Right Precentral Gyrus | 7.302 (1.054) | 6.433 (0.953) | 4.278 | <0.001** | [0.46 1.27] | 0.871 |
| Right Precuneus | 7.78 (1.022) | 6.979 (0.895) | 4.118 | <0.001** | [0.41 1.19] | 0.843 |
| Left Parahippocampus Gyrus | 2.583 (0.249) | 2.368 (0.261) | 4.219 | <0.001** | [0.11 0.32] | 0.840 |
| Left Precentral Gyrus | 6.972 (1.199) | 6.097 (0.984) | 3.915 | <0.001** | [0.43 1.32] | 0.810 |
| Left Superior Parietal Lobule | 7.442 (1.108) | 6.605 (0.988) | 3.940 | <0.001** | [0.41 1.26] | 0.805 |
| Right Parahippocampus Gyrus | 2.462 (0.255) | 2.246 (0.277) | 4.062 | <0.001** | [0.11 0.32] | 0.804 |
| Right Postcentral Gyrus | 5.908 (0.854) | 5.193 (0.913) | 4.053 | <0.001** | [0.36 1.06] | 0.804 |
| Right Angular Gyrus | 7.466 (0.952) | 6.774 (0.806) | 3.866 | <0.001** | [0.34 1.05] | 0.796 |
| Left Entorhinal Area | 1.656 (0.181) | 1.515 (0.18) | 3.916 | <0.001** | [0.07 0.21] | 0.786 |
| Left Middle Temporal Gyrus | 10.79 (1.146) | 9.943 (1.032) | 3.841 | <0.001** | [0.41 1.29] | 0.783 |
| Right Planum Polare | 1.39 (0.193) | 1.253 (0.161) | 3.784 | <0.001** | [0.06 0.21] | 0.780 |
| Left Posterior Cingulate Gyrus | 3.343 (0.424) | 3.045 (0.354) | 3.759 | <0.001** | [0.14 0.46] | 0.775 |
| Right Hippocampus | 3.129 (0.282) | 2.916 (0.273) | 3.821 | <0.001** | [0.1 0.32] | 0.770 |
| Left Inferior Temporal Gyrus | 9.123 (1.133) | 8.316 (0.991) | 3.746 | <0.001** | [0.38 1.24] | 0.767 |
| Right Ventral Ventricle | 0.984 (0.087) | 0.912 (0.1) | 3.900 | <0.001** | [0.04 0.11] | 0.766 |
| Right Posterior Cingulate Gyrus | 2.923 (0.359) | 2.674 (0.304) | 3.688 | <0.001** | [0.11 0.38] | 0.759 |
| Left Planum Polare | 1.468 (0.192) | 1.33 (0.174) | 3.729 | <0.001** | [0.06 0.21] | 0.759 |
| Left Ventral Ventricle | 0.985 (0.104) | 0.909 (0.097) | 3.730 | <0.001** | [0.04 0.12] | 0.756 |
| Right Amygdala | 0.795 (0.092) | 0.731 (0.079) | 3.665 | <0.001** | [0.03 0.1 ] | 0.753 |
| Left Angular Gyrus | 6.767 (0.933) | 6.146 (0.748) | 3.603 | <0.001** | [0.28 0.96] | 0.748 |
| Left Medial Orbital Gyrus | 3.303 (0.315) | 3.067 (0.315) | 3.721 | <0.001** | [0.11 0.36] | 0.746 |
| Left Supramarginal Gyrus | 6.471 (0.875) | 5.853 (0.794) | 3.660 | <0.001** | [0.28 0.95] | 0.745 |
| Left Hippocampus | 2.955 (0.254) | 2.773 (0.238) | 3.671 | <0.001** | [0.08 0.28] | 0.744 |
| Left Superior Temporal Gyrus | 5.197 (0.613) | 4.778 (0.524) | 3.621 | <0.001** | [0.19 0.65] | 0.744 |
| Left Anterior Orbital Gyrus | 1.316 (0.149) | 1.206 (0.148) | 3.676 | <0.001** | [0.05 0.17] | 0.737 |
| Left Middle Cingulate Gyrus | 3.43 (0.412) | 3.172 (0.309) | 3.461 | 0.002* | [0.11 0.41] | 0.725 |
| Right Entorhinal Area | 1.734 (0.175) | 1.602 (0.187) | 3.648 | <0.001** | [0.06 0.2 ] | 0.723 |
| Left Amygdala | 0.798 (0.098) | 0.733 (0.083) | 3.506 | <0.001** | [0.03 0.1 ] | 0.721 |
| Right Middle Frontal Gyrus | 13.767 (1.796) | 12.456 (1.84) | 3.601 | <0.001** | [0.59 2.03] | 0.719 |
| Left Cuneus | 2.845 (0.507) | 2.52 (0.409) | 3.465 | 0.002* | [0.14 0.51] | 0.718 |
| Right Superior Temporal Gyrus | 5.356 (0.742) | 4.868 (0.63) | 3.491 | <0.001** | [0.21 0.77] | 0.718 |
| Right Cuneus | 2.606 (0.486) | 2.309 (0.362) | 3.378 | 0.002* | [0.12 0.47] | 0.709 |
| Left Fusiform Gyrus | 5.465 (0.581) | 5.044 (0.603) | 3.559 | <0.001** | [0.19 0.66] | 0.709 |
| Left Calcarine and Cerebrum | 1.998 (0.366) | 1.781 (0.269) | 3.309 | 0.003* | [0.09 0.35] | 0.695 |
| Right Temporal Pole | 7.371 (0.869) | 6.754 (0.911) | 3.471 | <0.001** | [0.26 0.97] | 0.691 |
| Left Middle Occipital Gyrus | 4.301 (0.458) | 3.948 (0.574) | 3.451 | 0.002* | [0.15 0.56] | 0.668 |
| Right Middle Temporal Gyrus | 11.078 (1.165) | 10.29 (1.204) | 3.326 | 0.002* | [0.32 1.26] | 0.663 |
| Left Posterior Insula | 1.808 (0.211) | 1.677 (0.187) | 3.245 | 0.003* | [0.05 0.21] | 0.663 |
| Right Temporal Transverse Gyrus | 0.957 (0.164) | 0.859 (0.137) | 3.202 | 0.003* | [0.04 0.16] | 0.661 |
| Cerebellar Lobule Cerebellar Vermal Lobules I-V | 3.124 (0.416) | 2.87 (0.361) | 3.208 | 0.003* | [0.1 0.41] | 0.658 |
| Right Inferior Occipital Gyrus | 4.367 (0.625) | 4.016 (0.457) | 3.126 | 0.004* | [0.13 0.58] | 0.658 |
| Right Temporal | 1.476 (0.215) | 1.342 (0.195) | 3.217 | 0.003* | [0.05 0.22] | 0.655 |
| Left Superior Occipital Gyrus | 2.074 (0.333) | 1.862 (0.321) | 3.220 | 0.003* | [0.08 0.34] | 0.649 |
| Right Superior Frontal Gyrus | 10.61 (1.467) | 9.69 (1.386) | 3.199 | 0.003* | [0.35 1.49] | 0.647 |
| Left Superior Frontal Gyrus | 10.604 (1.65) | 9.576 (1.587) | 3.155 | 0.004* | [0.38 1.67] | 0.636 |
| Right Supramarginal Gyrus | 5.757 (0.82) | 5.229 (0.852) | 3.156 | 0.004* | [0.2 0.86] | 0.629 |
| Right Calcarine and Cerebrum | 1.662 (0.337) | 1.491 (0.216) | 2.916 | 0.007* | [0.05 0.29] | 0.625 |
| Right Supplementary Motor Cortex | 3.738 (0.513) | 3.424 (0.495) | 3.101 | 0.004* | [0.11 0.52] | 0.625 |
| Left Middle Frontal Gyrus | 13.56 (1.815) | 12.467 (1.709) | 3.078 | 0.005* | [0.39 1.8 ] | 0.623 |
| Right Anterior Orbital Gyrus | 1.429 (0.159) | 1.322 (0.182) | 3.163 | 0.003* | [0.04 0.17] | 0.621 |
| Left Supplementary Motor Cortex | 3.846 (0.554) | 3.516 (0.526) | 3.033 | 0.005* | [0.11 0.55] | 0.613 |
| Right Subcallosal Area | 0.995 (0.14) | 0.917 (0.117) | 2.970 | 0.006* | [0.03 0.13] | 0.612 |
| Left Lingual Gyrus | 5.369 (0.788) | 4.922 (0.694) | 2.973 | 0.006* | [0.15 0.75] | 0.608 |
| Right Superior Parietal Lobule | 7.139 (0.932) | 6.564 (0.958) | 3.046 | 0.005* | [0.2 0.95] | 0.608 |
| Right Inferior Temporal Gyrus | 9.197 (1.143) | 8.549 (1.02) | 2.958 | 0.006* | [0.21 1.08] | 0.604 |
| Right Fusiform Gyrus | 5.723 (0.721) | 5.315 (0.65) | 2.936 | 0.007* | [0.13 0.68] | 0.598 |
| Left Precuneus | 8.15 (1.048) | 7.59 (0.854) | 2.879 | 0.008* | [0.17 0.95] | 0.596 |
| Right Lingual Gyrus | 5.316 (0.838) | 4.86 (0.711) | 2.886 | 0.008* | [0.14 0.77] | 0.594 |
| Right Middle Cingulate Gyrus | 3.401 (0.422) | 3.18 (0.342) | 2.813 | 0.009* | [0.06 0.38] | 0.583 |
| Left Inferior Occipital Gyrus | 4.317 (0.659) | 3.969 (0.551) | 2.812 | 0.009* | [0.1 0.59] | 0.580 |
| Left Frontal Pole | 2.608 (0.375) | 2.39 (0.378) | 2.891 | 0.007* | [0.07 0.37] | 0.579 |
| Left Central Operculum | 3.076 (0.412) | 2.852 (0.367) | 2.831 | 0.009* | [0.07 0.38] | 0.578 |
| Right Medial Frontal Cerebrum | 1.435 (0.224) | 1.312 (0.205) | 2.842 | 0.008* | [0.04 0.21] | 0.578 |
| Left Superior Medial Frontal Gyrus | 4.856 (0.596) | 4.495 (0.651) | 2.901 | 0.007* | [0.11 0.61] | 0.574 |
| Right Medial Orbital Gyrus | 3.223 (0.354) | 3.016 (0.366) | 2.873 | 0.008* | [0.06 0.35] | 0.573 |
| Right Putamen | 2.632 (0.496) | 2.384 (0.384) | 2.735 | 0.011* | [0.07 0.43] | 0.571 |
| Left Temporal | 1.53 (0.257) | 1.396 (0.224) | 2.746 | 0.011* | [0.04 0.23] | 0.563 |
| Left Parietal Operculum | 1.816 (0.289) | 1.657 (0.278) | 2.786 | 0.01* | [0.05 0.27] | 0.562 |
| Right Posterior Insula | 1.957 (0.242) | 1.832 (0.21) | 2.736 | 0.011* | [0.03 0.22] | 0.561 |
| Left Triangular Inferior Frontal Gyrus | 2.56 (0.413) | 2.369 (0.276) | 2.632 | 0.015* | [0.05 0.34] | 0.561 |
| Right Central Operculum | 3.125 (0.409) | 2.9 (0.401) | 2.771 | 0.01* | [0.06 0.39] | 0.557 |
| Right Superior Medial Frontal Gyrus | 6.23 (0.851) | 5.745 (0.891) | 2.789 | 0.01* | [0.14 0.83] | 0.555 |
| Left Postcentral Gyrus | 6.402 (0.906) | 5.888 (0.938) | 2.784 | 0.01* | [0.15 0.88] | 0.555 |
| Left Caudate | 2.351 (0.286) | 2.172 (0.353) | 2.833 | 0.008* | [0.05 0.31] | 0.550 |
| Left Lateral Orbital Gyrus | 1.716 (0.239) | 1.596 (0.203) | 2.662 | 0.013* | [0.03 0.21] | 0.548 |
| Left Medial Precentral Gyrus | 1.634 (0.202) | 1.499 (0.277) | 2.859 | 0.008* | [0.04 0.23] | 0.546 |
| Right Inferior Frontal Orbital Gyrus | 0.999 (0.193) | 0.908 (0.146) | 2.599 | 0.016* | [0.02 0.16] | 0.544 |
| Left Putamen | 2.72 (0.5) | 2.485 (0.376) | 2.593 | 0.016* | [0.05 0.42] | 0.543 |
| Left Frontal Operculum | 1.427 (0.171) | 1.334 (0.173) | 2.677 | 0.013* | [0.02 0.16] | 0.536 |
| Left Basal Cerebrum and Forebrain Brain | 0.32 (0.05) | 0.297 (0.04) | 2.557 | 0.017* | [0.01 0.04] | 0.531 |
| Right Medial Precentral Gyrus | 1.486 (0.27) | 1.345 (0.271) | 2.615 | 0.015* | [0.03 0.25] | 0.524 |
| Left Anterior Cingulate Gyrus | 3.616 (0.482) | 3.364 (0.487) | 2.596 | 0.016* | [0.06 0.44] | 0.520 |
| Right Anterior Insula | 3.472 (0.401) | 3.276 (0.361) | 2.542 | 0.018* | [0.04 0.35] | 0.518 |
| Cerebellar Lobule Cerebellar Vermal Lobules VI-VII | 1.609 (0.299) | 1.472 (0.247) | 2.455 | 0.023* | [0.03 0.25] | 0.507 |
| Right Basal Cerebrum and Forebrain Brain | 0.32 (0.044) | 0.299 (0.039) | 2.430 | 0.024* | [0. 0.04] | 0.498 |
| Left Inferior Frontal Gyrus | 2.257 (0.319) | 2.108 (0.288) | 2.435 | 0.023* | [0.03 0.27] | 0.496 |
| Left Exterior Cerebellum | 32.976 (4.536) | 30.967 (3.731) | 2.376 | 0.027* | [0.33 3.69] | 0.491 |
| Left Occipital Pole | 1.831 (0.447) | 1.642 (0.333) | 2.335 | 0.03* | [0.03 0.35] | 0.490 |
| Right Posterior Orbital Gyrus | 2.026 (0.323) | 1.874 (0.302) | 2.409 | 0.025* | [0.03 0.28] | 0.488 |
| Left Subcallosal Area | 1.04 (0.156) | 0.973 (0.123) | 2.332 | 0.03* | [0.01 0.12] | 0.485 |
| Right Medial Postcentral Gyrus | 0.584 (0.132) | 0.522 (0.128) | 2.344 | 0.029* | [0.01 0.11] | 0.472 |
| Right Occipital Fusiform Gyrus | 2.478 (0.409) | 2.308 (0.334) | 2.245 | 0.037* | [0.02 0.32] | 0.464 |
| Left Anterior Insula | 3.542 (0.365) | 3.367 (0.408) | 2.276 | 0.033* | [0.02 0.33] | 0.448 |
| Left Occipital Fusiform Gyrus | 2.615 (0.434) | 2.445 (0.361) | 2.094 | 0.052 | [0.01 0.33] | 0.432 |
| Left Temporal Transverse Gyrus | 0.935 (0.177) | 0.865 (0.156) | 2.099 | 0.051 | [0. 0.14] | 0.429 |
| Right Gyrus Rectus | 1.699 (0.258) | 1.595 (0.234) | 2.082 | 0.052 | [0. 0.2] | 0.424 |
| Left Pallidum | 0.261 (0.107) | 0.224 (0.075) | 1.954 | 0.070 | [-0. 0.07] | 0.413 |
| Right Lateral Orbital Gyrus | 1.568 (0.193) | 1.475 (0.243) | 2.135 | 0.046* | [0.01 0.18] | 0.413 |
| Right Exterior Cerebellum | 32.656 (4.626) | 31.004 (3.521) | 1.963 | 0.069 | [-0.02 3.33] | 0.411 |
| Right Frontal Pole | 2.738 (0.348) | 2.593 (0.401) | 1.943 | 0.070 | [-0. 0.29] | 0.381 |
| Left Posterior Orbital Gyrus | 2.326 (0.342) | 2.2 (0.333) | 1.868 | 0.082 | [-0.01 0.26] | 0.376 |
| Right Anterior Cingulate Gyrus | 2.982 (0.461) | 2.826 (0.424) | 1.743 | 0.106 | [-0.02 0.33] | 0.354 |
| Right Caudate | 2.654 (0.338) | 2.534 (0.359) | 1.730 | 0.109 | [-0.02 0.26] | 0.343 |
| Left Inferior Frontal Orbital Gyrus | 1.212 (0.198) | 1.147 (0.184) | 1.672 | 0.121 | [-0.01 0.14] | 0.339 |
| Right Parietal Operculum | 1.499 (0.275) | 1.413 (0.243) | 1.645 | 0.126 | [-0.02 0.19] | 0.337 |
| Right Accumbens | 0.328 (0.043) | 0.314 (0.043) | 1.676 | 0.121 | [-0. 0.03] | 0.336 |
| Right Occipital Pole | 1.684 (0.397) | 1.568 (0.299) | 1.602 | 0.137 | [-0.03 0.26] | 0.336 |
| Left Accumbens | 0.346 (0.05) | 0.332 (0.043) | 1.495 | 0.167 | [-0. 0.03] | 0.307 |
| Right Frontal Operculum | 1.458 (0.218) | 1.404 (0.172) | 1.358 | 0.212 | [-0.03 0.13] | 0.282 |
| Cerebellar Lobule Cerebellar Vermal Lobules VIII-X | 1.376 (0.186) | 1.323 (0.194) | 1.396 | 0.198 | [-0.02 0.13] | 0.278 |
| Left Medial Postcentral Gyrus | 0.62 (0.101) | 0.592 (0.151) | 1.130 | 0.298 | [-0.02 0.08] | 0.213 |
| Right Inferior Frontal Gyrus | 2.353 (0.349) | 2.283 (0.317) | 1.034 | 0.344 | [-0.06 0.2 ] | 0.211 |
| Right Triangular Inferior Frontal Gyrus | 2.45 (0.395) | 2.402 (0.336) | 0.642 | 0.576 | [-0.1 0.2] | 0.132 |
| Right Pallidum | 0.236 (0.099) | 0.225 (0.098) | 0.540 | 0.641 | [-0.03 0.05] | 0.109 |
| Optic Chiasm | 0.117 (0.024) | 0.117 (0.019) | -0.034 | 0.978 | [-0.01 0.01] | 0.007 |
| **Cortical thickness** | | | | | | |
| rinferiorparietal | 2.256 (0.105) | 2.044 (0.126) | 9.245 | <0.001** | [0.17 0.26] | 1.801 |
| linferiorparietal | 2.295 (0.116) | 2.09 (0.117) | 8.784 | <0.001** | [0.16 0.25] | 1.758 |
| rsuperiorparietal | 1.972 (0.121) | 1.731 (0.149) | 9.004 | <0.001** | [0.19 0.29] | 1.747 |
| rsupramarginal | 2.293 (0.113) | 2.082 (0.14) | 8.442 | <0.001** | [0.16 0.26] | 1.637 |
| lsuperiorparietal | 1.947 (0.116) | 1.734 (0.141) | 8.331 | <0.001** | [0.16 0.26] | 1.620 |
| rS_intrapariet_and_P_trans | 2.042 (0.133) | 1.8 (0.16) | 8.313 | <0.001** | [0.18 0.3 ] | 1.619 |
| lsupramarginal | 2.294 (0.1) | 2.104 (0.132) | 8.308 | <0.001** | [0.14 0.24] | 1.596 |
| lS_intrapariet_and_P_trans | 2.021 (0.133) | 1.802 (0.143) | 7.973 | <0.001** | [0.16 0.27] | 1.580 |
| lS_front_inf | 2.132 (0.133) | 1.919 (0.136) | 7.896 | <0.001** | [0.16 0.27] | 1.579 |
| lG_pariet_inf_Angular | 2.386 (0.144) | 2.155 (0.151) | 7.847 | <0.001** | [0.17 0.29] | 1.561 |
| rS_temporal_sup | 2.378 (0.094) | 2.204 (0.124) | 8.060 | <0.001** | [0.13 0.22] | 1.548 |
| rprecuneus | 2.076 (0.12) | 1.874 (0.138) | 7.871 | <0.001** | [0.15 0.25] | 1.544 |
| rmiddletemporal | 2.573 (0.116) | 2.371 (0.145) | 7.814 | <0.001** | [0.15 0.25] | 1.513 |
| rG_precuneus | 2.134 (0.14) | 1.89 (0.179) | 7.729 | <0.001** | [0.18 0.31] | 1.491 |
| rS_postcentral | 1.905 (0.138) | 1.674 (0.166) | 7.651 | <0.001** | [0.17 0.29] | 1.490 |
| lrostralmiddlefrontal | 2.219 (0.106) | 2.034 (0.137) | 7.713 | <0.001** | [0.14 0.23] | 1.487 |
| lsuperiortemporal | 2.488 (0.132) | 2.273 (0.153) | 7.591 | <0.001** | [0.16 0.27] | 1.486 |
| rG_pariet_inf_Supramar | 2.355 (0.136) | 2.137 (0.155) | 7.546 | <0.001** | [0.16 0.28] | 1.483 |
| lG_pariet_inf_Supramar | 2.381 (0.12) | 2.182 (0.146) | 7.545 | <0.001** | [0.15 0.25] | 1.467 |
| rG_parietal_sup | 2.095 (0.174) | 1.815 (0.204) | 7.464 | <0.001** | [0.21 0.36] | 1.460 |
| rS_cingul_Marginalis | 2.039 (0.127) | 1.85 (0.132) | 7.316 | <0.001** | [0.14 0.24] | 1.458 |
| rG_temporal_middle | 2.624 (0.14) | 2.403 (0.16) | 7.391 | <0.001** | [0.16 0.28] | 1.452 |
| rS_oc_sup_and_transversal | 1.924 (0.143) | 1.726 (0.138) | 7.034 | <0.001** | [0.14 0.25] | 1.419 |
| rG_occipital_middle | 2.151 (0.12) | 1.949 (0.157) | 7.368 | <0.001** | [0.15 0.26] | 1.418 |
| rS_front_sup | 2.333 (0.135) | 2.072 (0.213) | 7.522 | <0.001** | [0.19 0.33] | 1.410 |
| lG_front_middle | 2.297 (0.145) | 2.053 (0.192) | 7.309 | <0.001** | [0.18 0.31] | 1.404 |
| rG_pariet_inf_Angular | 2.3 (0.151) | 2.066 (0.178) | 7.182 | <0.001** | [0.17 0.3 ] | 1.403 |
| lG_temp_sup_Plan_tempo | 2.286 (0.144) | 2.072 (0.159) | 7.100 | <0.001** | [0.15 0.27] | 1.401 |
| lG_precuneus | 2.34 (0.136) | 2.152 (0.133) | 6.956 | <0.001** | [0.13 0.24] | 1.399 |
| lS_postcentral | 1.89 (0.128) | 1.691 (0.154) | 7.108 | <0.001** | [0.14 0.25] | 1.384 |
| lprecuneus | 2.224 (0.109) | 2.068 (0.117) | 6.918 | <0.001** | [0.11 0.2 ] | 1.372 |
| lG_parietal_sup | 2.06 (0.152) | 1.834 (0.175) | 6.966 | <0.001** | [0.16 0.29] | 1.366 |
| lsuperiorfrontal | 2.322 (0.144) | 2.076 (0.202) | 7.166 | <0.001** | [0.18 0.31] | 1.364 |
| lcaudalmiddlefrontal | 2.255 (0.165) | 1.984 (0.222) | 7.073 | <0.001** | [0.2 0.35] | 1.355 |
| rsuperiorfrontal | 2.415 (0.125) | 2.179 (0.202) | 7.256 | <0.001** | [0.17 0.3 ] | 1.355 |
| lS_temporal_sup | 2.32 (0.119) | 2.164 (0.115) | 6.650 | <0.001** | [0.11 0.2 ] | 1.340 |
| lG_and_S_subcentral | 2.301 (0.146) | 2.082 (0.175) | 6.872 | <0.001** | [0.16 0.28] | 1.339 |
| rfusiform | 2.236 (0.125) | 2.058 (0.14) | 6.761 | <0.001** | [0.13 0.23] | 1.331 |
| lS_precentral_inf_part | 2.152 (0.151) | 1.928 (0.183) | 6.753 | <0.001** | [0.16 0.29] | 1.315 |
| rcaudalmiddlefrontal | 2.251 (0.152) | 2.012 (0.206) | 6.755 | <0.001** | [0.17 0.31] | 1.292 |
| lG_temporal_inf | 2.523 (0.143) | 2.331 (0.153) | 6.476 | <0.001** | [0.13 0.25] | 1.285 |
| linferiortemporal | 2.47 (0.115) | 2.309 (0.135) | 6.501 | <0.001** | [0.11 0.21] | 1.272 |
| rlateraloccipital | 1.814 (0.097) | 1.687 (0.102) | 6.386 | <0.001** | [0.09 0.17] | 1.271 |
| lG_front_sup | 2.238 (0.169) | 1.972 (0.234) | 6.655 | <0.001** | [0.19 0.35] | 1.269 |
| lS_front_middle | 2.205 (0.115) | 2.033 (0.148) | 6.572 | <0.001** | [0.12 0.22] | 1.266 |
| lparsopercularis | 2.42 (0.141) | 2.232 (0.155) | 6.372 | <0.001** | [0.13 0.25] | 1.259 |
| rG_front_sup | 2.292 (0.151) | 2.042 (0.23) | 6.593 | <0.001** | [0.17 0.32] | 1.241 |
| lG_and_S_cingul_Mid_Post | 2.303 (0.119) | 2.141 (0.14) | 6.324 | <0.001** | [0.11 0.21] | 1.235 |
| lS_oc_sup_and_transversal | 1.932 (0.13) | 1.764 (0.142) | 6.199 | <0.001** | [0.11 0.22] | 1.226 |
| lpostcentral | 1.558 (0.102) | 1.423 (0.117) | 6.220 | <0.001** | [0.09 0.18] | 1.221 |
| rpostcentral | 1.569 (0.122) | 1.423 (0.117) | 6.036 | <0.001** | [0.1 0.19] | 1.218 |
| rS_subparietal | 2.157 (0.137) | 1.996 (0.13) | 6.003 | <0.001** | [0.11 0.21] | 1.214 |
| lLat_Fis_post | 2.192 (0.138) | 2.018 (0.147) | 6.086 | <0.001** | [0.12 0.23] | 1.208 |
| lS_front_sup | 2.331 (0.145) | 2.102 (0.217) | 6.376 | <0.001** | [0.16 0.3 ] | 1.203 |
| rG_temp_sup_Plan_tempo | 2.311 (0.184) | 2.085 (0.196) | 5.965 | <0.001** | [0.15 0.3 ] | 1.184 |
| lG_temp_sup_Lateral | 2.58 (0.161) | 2.367 (0.193) | 6.060 | <0.001** | [0.14 0.28] | 1.181 |
| rG_and_S_subcentral | 2.327 (0.171) | 2.111 (0.191) | 5.980 | <0.001** | [0.14 0.29] | 1.178 |
| rG_front_middle | 2.29 (0.133) | 2.1 (0.179) | 6.136 | <0.001** | [0.13 0.25] | 1.176 |
| rS_precentral_sup_part | 2 (0.216) | 1.722 (0.252) | 5.989 | <0.001** | [0.19 0.37] | 1.172 |
| lparstriangularis | 2.315 (0.113) | 2.162 (0.143) | 6.035 | <0.001** | [0.1 0.2] | 1.167 |
| rG_and_S_cingul_Mid_Post | 2.253 (0.15) | 2.08 (0.148) | 5.789 | <0.001** | [0.11 0.23] | 1.163 |
| lS_cingul_Marginalis | 2.14 (0.126) | 1.981 (0.145) | 5.922 | <0.001** | [0.11 0.21] | 1.161 |
| rrostralmiddlefrontal | 2.192 (0.1) | 2.047 (0.14) | 6.087 | <0.001** | [0.1 0.19] | 1.159 |
| rS_interm_prim_Jensen | 2.18 (0.156) | 1.994 (0.166) | 5.762 | <0.001** | [0.12 0.25] | 1.144 |
| llateralorbitofrontal | 2.583 (0.14) | 2.415 (0.152) | 5.766 | <0.001** | [0.11 0.23] | 1.142 |
| lcuneus | 1.596 (0.144) | 1.435 (0.139) | 5.654 | <0.001** | [0.1 0.22] | 1.139 |
| lS_occipital_ant | 2.093 (0.195) | 1.89 (0.167) | 5.491 | <0.001** | [0.13 0.28] | 1.128 |
| rS_circular_insula_sup | 2.727 (0.144) | 2.563 (0.147) | 5.638 | <0.001** | [0.11 0.22] | 1.127 |
| rparsopercularis | 2.39 (0.14) | 2.212 (0.17) | 5.776 | <0.001** | [0.12 0.24] | 1.123 |
| rS_precentral_inf_part | 2.257 (0.131) | 2.077 (0.178) | 5.859 | <0.001** | [0.12 0.24] | 1.120 |
| lS_precentral_sup_part | 1.859 (0.226) | 1.607 (0.224) | 5.581 | <0.001** | [0.16 0.34] | 1.120 |
| rS_temporal_inf | 2.275 (0.131) | 2.125 (0.135) | 5.605 | <0.001** | [0.1 0.2] | 1.119 |
| rposteriorcingulate | 2.139 (0.114) | 2.01 (0.116) | 5.592 | <0.001** | [0.08 0.17] | 1.119 |
| lS_circular_insula_inf | 2.881 (0.172) | 2.68 (0.187) | 5.613 | <0.001** | [0.13 0.27] | 1.111 |
| rsuperiortemporal | 2.556 (0.16) | 2.362 (0.185) | 5.659 | <0.001** | [0.13 0.26] | 1.109 |
| rlateralorbitofrontal | 2.368 (0.139) | 2.219 (0.131) | 5.466 | <0.001** | [0.09 0.2 ] | 1.106 |
| lG_front_inf_Triangul | 2.282 (0.136) | 2.091 (0.196) | 5.827 | <0.001** | [0.13 0.26] | 1.105 |
| rS_front_inf | 2.042 (0.131) | 1.896 (0.135) | 5.501 | <0.001** | [0.09 0.2 ] | 1.098 |
| llateraloccipital | 1.819 (0.106) | 1.694 (0.119) | 5.575 | <0.001** | [0.08 0.17] | 1.098 |
| lS_circular_insula_sup | 2.684 (0.136) | 2.53 (0.143) | 5.511 | <0.001** | [0.1 0.21] | 1.096 |
| lS_parieto_occipital | 1.985 (0.146) | 1.815 (0.162) | 5.552 | <0.001** | [0.11 0.23] | 1.096 |
| lG_front_inf_Opercular | 2.475 (0.158) | 2.275 (0.199) | 5.652 | <0.001** | [0.13 0.27] | 1.093 |
| rG_front_inf_Opercular | 2.544 (0.17) | 2.345 (0.194) | 5.499 | <0.001** | [0.13 0.27] | 1.081 |
| lG_and_S_cingul_Ant | 2.485 (0.105) | 2.338 (0.155) | 5.692 | <0.001** | [0.1 0.2] | 1.077 |
| lfusiform | 2.239 (0.123) | 2.098 (0.137) | 5.464 | <0.001** | [0.09 0.19] | 1.077 |
| lG_cuneus | 1.513 (0.116) | 1.373 (0.139) | 5.521 | <0.001** | [0.09 0.19] | 1.076 |
| lG_occipital_middle | 2.177 (0.134) | 2.02 (0.154) | 5.478 | <0.001** | [0.1 0.21] | 1.075 |
| lLat_Fis_ant_Vertical | 2.407 (0.188) | 2.212 (0.179) | 5.295 | <0.001** | [0.12 0.27] | 1.070 |
| rbankssts | 2.365 (0.149) | 2.198 (0.163) | 5.371 | <0.001** | [0.11 0.23] | 1.062 |
| rG_oc_temp_lat_fusifor | 2.082 (0.148) | 1.914 (0.168) | 5.356 | <0.001** | [0.11 0.23] | 1.052 |
| rLat_Fis_post | 2.422 (0.16) | 2.251 (0.163) | 5.250 | <0.001** | [0.11 0.23] | 1.049 |
| lprecentral | 1.619 (0.148) | 1.456 (0.161) | 5.304 | <0.001** | [0.1 0.22] | 1.049 |
| ltransversetemporal | 1.697 (0.179) | 1.509 (0.179) | 5.223 | <0.001** | [0.12 0.26] | 1.048 |
| lG_temp_sup_G_T_transv | 1.689 (0.177) | 1.483 (0.211) | 5.347 | <0.001** | [0.13 0.28] | 1.044 |
| lmiddletemporal | 2.518 (0.129) | 2.371 (0.149) | 5.309 | <0.001** | [0.09 0.2 ] | 1.042 |
| rG_orbital | 2.32 (0.174) | 2.151 (0.154) | 5.083 | <0.001** | [0.1 0.24] | 1.039 |
| lparacentral | 1.75 (0.154) | 1.569 (0.188) | 5.347 | <0.001** | [0.11 0.25] | 1.039 |
| lG_temp_sup_Plan_polar | 3.258 (0.244) | 2.956 (0.323) | 5.391 | <0.001** | [0.19 0.41] | 1.035 |
| rprecentral | 1.662 (0.123) | 1.518 (0.152) | 5.253 | <0.001** | [0.09 0.2 ] | 1.019 |
| lG_orbital | 2.538 (0.169) | 2.355 (0.187) | 5.141 | <0.001** | [0.11 0.25] | 1.015 |
| rG_temp_sup_Lateral | 2.626 (0.204) | 2.419 (0.206) | 5.054 | <0.001** | [0.13 0.29] | 1.012 |
| lmedialorbitofrontal | 2.253 (0.146) | 2.116 (0.127) | 4.908 | <0.001** | [0.08 0.19] | 1.006 |
| linsula | 3.037 (0.177) | 2.841 (0.208) | 5.112 | <0.001** | [0.12 0.27] | 1.000 |
| rS_oc_middle_and_Lunatus | 1.943 (0.139) | 1.793 (0.159) | 5.069 | <0.001** | [0.09 0.21] | 0.995 |
| rS_collat_transv_ant | 2.533 (0.194) | 2.346 (0.185) | 4.912 | <0.001** | [0.11 0.26] | 0.992 |
| lG_and_S_transv_frontopol | 2.303 (0.179) | 2.123 (0.186) | 4.932 | <0.001** | [0.11 0.25] | 0.983 |
| lG_temporal_middle | 2.562 (0.141) | 2.41 (0.165) | 5.011 | <0.001** | [0.09 0.21] | 0.981 |
| lposteriorcingulate | 2.175 (0.091) | 2.078 (0.105) | 5.000 | <0.001** | [0.06 0.14] | 0.980 |
| rPole_temporal | 2.799 (0.245) | 2.549 (0.269) | 4.884 | <0.001** | [0.15 0.35] | 0.965 |
| lS_orbital_lateral | 2.136 (0.133) | 1.985 (0.171) | 4.999 | <0.001** | [0.09 0.21] | 0.964 |
| rparacentral | 1.577 (0.14) | 1.438 (0.148) | 4.830 | <0.001** | [0.08 0.2 ] | 0.960 |
| rG_temp_sup_G_T_transv | 2.089 (0.281) | 1.811 (0.297) | 4.812 | <0.001** | [0.16 0.39] | 0.956 |
| lG_and_S_cingul_Mid_Ant | 2.472 (0.163) | 2.308 (0.178) | 4.827 | <0.001** | [0.1 0.23] | 0.955 |
| rG_occipital_sup | 1.687 (0.155) | 1.524 (0.181) | 4.875 | <0.001** | [0.1 0.23] | 0.954 |
| rparstriangularis | 2.281 (0.141) | 2.135 (0.161) | 4.840 | <0.001** | [0.09 0.21] | 0.950 |
| rS_parieto_occipital | 1.805 (0.168) | 1.641 (0.179) | 4.755 | <0.001** | [0.1 0.23] | 0.944 |
| rinferiortemporal | 2.414 (0.136) | 2.29 (0.13) | 4.638 | <0.001** | [0.07 0.18] | 0.937 |
| lS_temporal_inf | 2.277 (0.129) | 2.153 (0.134) | 4.693 | <0.001** | [0.07 0.18] | 0.936 |
| rG_oc_temp_med_Lingual | 1.557 (0.104) | 1.451 (0.123) | 4.687 | <0.001** | [0.06 0.15] | 0.916 |
| rS_collat_transv_post | 1.687 (0.161) | 1.535 (0.172) | 4.595 | <0.001** | [0.09 0.22] | 0.912 |
| rS_orbital_lateral | 2.175 (0.159) | 2.03 (0.16) | 4.527 | <0.001** | [0.08 0.21] | 0.907 |
| rinsula | 3.037 (0.194) | 2.863 (0.19) | 4.511 | <0.001** | [0.1 0.25] | 0.907 |
| lG_and_S_frontomargin | 2.139 (0.126) | 2.013 (0.148) | 4.610 | <0.001** | [0.07 0.18] | 0.902 |
| lS_orbital_H_Shaped | 2.486 (0.142) | 2.347 (0.162) | 4.566 | <0.001** | [0.08 0.2 ] | 0.896 |
| rS_orbital_H_Shaped | 2.422 (0.146) | 2.286 (0.156) | 4.504 | <0.001** | [0.08 0.2 ] | 0.894 |
| rmedialorbitofrontal | 2.334 (0.118) | 2.224 (0.125) | 4.503 | <0.001** | [0.06 0.16] | 0.894 |
| lG_postcentral | 1.412 (0.132) | 1.28 (0.158) | 4.565 | <0.001** | [0.07 0.19] | 0.890 |
| rcuneus | 1.39 (0.121) | 1.28 (0.127) | 4.472 | <0.001** | [0.06 0.16] | 0.890 |
| rG_front_inf_Triangul | 2.31 (0.172) | 2.152 (0.182) | 4.461 | <0.001** | [0.09 0.23] | 0.887 |
| rG_and_S_cingul_Ant | 2.546 (0.13) | 2.418 (0.155) | 4.534 | <0.001** | [0.07 0.18] | 0.885 |
| rG_and_S_transv_frontopol | 2.206 (0.131) | 2.072 (0.166) | 4.566 | <0.001** | [0.08 0.19] | 0.882 |
| lS_temporal_transverse | 1.76 (0.223) | 1.583 (0.186) | 4.250 | <0.001** | [0.09 0.26] | 0.877 |
| rS_occipital_ant | 2.105 (0.159) | 1.96 (0.171) | 4.412 | <0.001** | [0.08 0.21] | 0.875 |
| rtransversetemporal | 2.008 (0.232) | 1.793 (0.256) | 4.434 | <0.001** | [0.12 0.31] | 0.875 |
| rS_oc_temp_med_and_Lingual | 2.046 (0.153) | 1.918 (0.142) | 4.300 | <0.001** | [0.07 0.19] | 0.872 |
| rG_postcentral | 1.361 (0.16) | 1.226 (0.156) | 4.277 | <0.001** | [0.07 0.2 ] | 0.861 |
| lG_occipital_sup | 1.662 (0.169) | 1.515 (0.174) | 4.292 | <0.001** | [0.08 0.22] | 0.857 |
| lS_suborbital | 2.237 (0.211) | 2.07 (0.182) | 4.177 | <0.001** | [0.09 0.25] | 0.857 |
| rlingual | 1.585 (0.093) | 1.501 (0.102) | 4.338 | <0.001** | [0.05 0.12] | 0.857 |
| lS_oc_middle_and_Lunatus | 1.918 (0.123) | 1.801 (0.15) | 4.315 | <0.001** | [0.06 0.17] | 0.839 |
| lS_subparietal | 2.233 (0.114) | 2.137 (0.115) | 4.180 | <0.001** | [0.05 0.14] | 0.837 |
| rparsorbitalis | 2.35 (0.19) | 2.181 (0.211) | 4.231 | <0.001** | [0.09 0.25] | 0.834 |
| rS_front_middle | 2.189 (0.119) | 2.066 (0.168) | 4.350 | <0.001** | [0.07 0.18] | 0.828 |
| rG_and_S_frontomargin | 2.122 (0.108) | 2.005 (0.164) | 4.355 | <0.001** | [0.06 0.17] | 0.821 |
| lG_precentral | 1.519 (0.197) | 1.347 (0.219) | 4.160 | <0.001** | [0.09 0.25] | 0.821 |
| rG_and_S_cingul_Mid_Ant | 2.534 (0.157) | 2.402 (0.163) | 4.115 | <0.001** | [0.07 0.2 ] | 0.820 |
| lPole_temporal | 3.017 (0.266) | 2.79 (0.287) | 4.123 | <0.001** | [0.12 0.34] | 0.817 |
| rG_precentral | 1.599 (0.157) | 1.445 (0.209) | 4.247 | <0.001** | [0.08 0.23] | 0.815 |
| lS_collat_transv_post | 1.745 (0.203) | 1.599 (0.16) | 3.913 | <0.001** | [0.07 0.22] | 0.814 |
| rG_cuneus | 1.314 (0.116) | 1.219 (0.121) | 4.033 | <0.001** | [0.05 0.14] | 0.804 |
| lS_collat_transv_ant | 2.553 (0.195) | 2.405 (0.177) | 3.939 | <0.001** | [0.07 0.22] | 0.802 |
| lfrontalpole | 2.304 (0.224) | 2.108 (0.263) | 4.066 | <0.001** | [0.1 0.29] | 0.795 |
| lS_oc_temp_lat | 2.287 (0.164) | 2.15 (0.184) | 3.944 | <0.001** | [0.07 0.21] | 0.777 |
| rS_oc_temp_lat | 2.303 (0.161) | 2.169 (0.182) | 3.947 | <0.001** | [0.07 0.2 ] | 0.777 |
| rPole_occipital | 1.438 (0.112) | 1.358 (0.098) | 3.789 | <0.001** | [0.04 0.12] | 0.776 |
| lG_rectus | 2.227 (0.157) | 2.108 (0.155) | 3.790 | <0.001** | [0.06 0.18] | 0.761 |
| rG_cingul_Post_dorsal | 2.516 (0.165) | 2.386 (0.175) | 3.818 | <0.001** | [0.06 0.2 ] | 0.759 |
| rG_front_inf_Orbital | 2.262 (0.251) | 2.077 (0.241) | 3.728 | <0.001** | [0.09 0.28] | 0.752 |
| lbankssts | 2.293 (0.158) | 2.174 (0.16) | 3.743 | <0.001** | [0.06 0.18] | 0.749 |
| rG_and_S_occipital_inf | 2.139 (0.16) | 2.023 (0.154) | 3.705 | <0.001** | [0.05 0.18] | 0.747 |
| ltemporalpole | 3.365 (0.331) | 3.076 (0.424) | 3.863 | <0.001** | [0.14 0.44] | 0.745 |
| lparsorbitalis | 2.459 (0.191) | 2.32 (0.191) | 3.644 | <0.001** | [0.06 0.22] | 0.730 |
| lG_oc_temp_med_Lingual | 1.618 (0.115) | 1.527 (0.133) | 3.717 | <0.001** | [0.04 0.14] | 0.728 |
| lS_oc_temp_med_and_Lingual | 2.088 (0.149) | 1.976 (0.16) | 3.655 | <0.001** | [0.05 0.17] | 0.725 |
| lG_oc_temp_lat_fusifor | 2.054 (0.138) | 1.942 (0.165) | 3.710 | <0.001** | [0.05 0.17] | 0.723 |
| rG_insular_short | 3.303 (0.344) | 3.074 (0.298) | 3.506 | <0.001** | [0.1 0.36] | 0.719 |
| lS_central | 1.128 (0.113) | 1.05 (0.105) | 3.539 | <0.001** | [0.03 0.12] | 0.718 |
| lrostralanteriorcingulate | 2.516 (0.122) | 2.407 (0.171) | 3.767 | <0.001** | [0.05 0.17] | 0.717 |
| lS_circular_insula_ant | 3.042 (0.311) | 2.845 (0.247) | 3.439 | 0.002* | [0.08 0.31] | 0.715 |
| rG_temporal_inf | 2.442 (0.165) | 2.329 (0.152) | 3.514 | <0.001** | [0.05 0.18] | 0.714 |
| llingual | 1.7 (0.108) | 1.622 (0.113) | 3.545 | <0.001** | [0.03 0.12] | 0.705 |
| rS_circular_insula_inf | 2.893 (0.197) | 2.733 (0.249) | 3.605 | <0.001** | [0.07 0.25] | 0.697 |
| lS_interm_prim_Jensen | 2.304 (0.182) | 2.161 (0.222) | 3.563 | <0.001** | [0.06 0.22] | 0.693 |
| rpericalcarine | 1.159 (0.1) | 1.092 (0.094) | 3.424 | 0.002* | [0.03 0.11] | 0.693 |
| lG_and_S_paracentral | 1.413 (0.155) | 1.292 (0.193) | 3.513 | <0.001** | [0.05 0.19] | 0.680 |
| rG_Ins_lg_and_S_cent_ins | 3.048 (0.306) | 2.842 (0.302) | 3.378 | 0.002* | [0.08 0.33] | 0.679 |
| rS_temporal_transverse | 1.816 (0.234) | 1.659 (0.231) | 3.364 | 0.002* | [0.06 0.25] | 0.676 |
| lG_Ins_lg_and_S_cent_ins | 2.987 (0.239) | 2.788 (0.33) | 3.531 | <0.001** | [0.09 0.31] | 0.674 |
| rS_orbital_med_olfact | 2.234 (0.161) | 2.132 (0.144) | 3.291 | 0.003* | [0.04 0.16] | 0.672 |
| lpericalcarine | 1.347 (0.116) | 1.264 (0.131) | 3.387 | 0.002* | [0.03 0.13] | 0.666 |
| rG_subcallosal | 2.897 (0.261) | 2.697 (0.327) | 3.429 | 0.002* | [0.08 0.32] | 0.664 |
| listhmuscingulate | 2.131 (0.133) | 2.045 (0.129) | 3.280 | 0.003* | [0.03 0.14] | 0.661 |
| rS_central | 1.119 (0.101) | 1.051 (0.104) | 3.270 | 0.003* | [0.03 0.11] | 0.653 |
| lG_and_S_occipital_inf | 2.026 (0.144) | 1.923 (0.167) | 3.310 | 0.002* | [0.04 0.16] | 0.648 |
| rrostralanteriorcingulate | 2.611 (0.162) | 2.51 (0.154) | 3.188 | 0.003* | [0.04 0.16] | 0.644 |
| rparahippocampal | 2.287 (0.238) | 2.145 (0.21) | 3.126 | 0.004* | [0.05 0.23] | 0.639 |
| rLat_Fis_ant_Vertical | 2.336 (0.187) | 2.203 (0.225) | 3.267 | 0.003* | [0.05 0.21] | 0.636 |
| lG_front_inf_Orbital | 2.501 (0.195) | 2.37 (0.213) | 3.210 | 0.003* | [0.05 0.21] | 0.635 |
| lS_orbital_med_olfact | 2.249 (0.161) | 2.145 (0.166) | 3.168 | 0.003* | [0.04 0.17] | 0.633 |
| rG_rectus | 2.292 (0.155) | 2.189 (0.171) | 3.199 | 0.003* | [0.04 0.17] | 0.631 |
| rG_and_S_paracentral | 1.27 (0.175) | 1.167 (0.164) | 3.032 | 0.005* | [0.04 0.17] | 0.614 |
| rLat_Fis_ant_Horizont | 2.185 (0.204) | 2.061 (0.208) | 2.993 | 0.006* | [0.04 0.21] | 0.598 |
| lPole_occipital | 1.453 (0.135) | 1.364 (0.16) | 3.024 | 0.005* | [0.03 0.15] | 0.590 |
| rS_circular_insula_ant | 2.789 (0.231) | 2.654 (0.229) | 2.914 | 0.007* | [0.04 0.23] | 0.585 |
| rG_oc_temp_med_Parahip | 2.71 (0.186) | 2.589 (0.232) | 2.924 | 0.007* | [0.04 0.2 ] | 0.567 |
| rtemporalpole | 3.116 (0.409) | 2.897 (0.389) | 2.734 | 0.011* | [0.06 0.38] | 0.553 |
| rG_temp_sup_Plan_polar | 3.186 (0.319) | 3.017 (0.303) | 2.706 | 0.012* | [0.05 0.29] | 0.547 |
| rS_suborbital | 2.288 (0.22) | 2.169 (0.223) | 2.685 | 0.012* | [0.03 0.21] | 0.537 |
| rS_calcarine | 1.506 (0.099) | 1.454 (0.11) | 2.526 | 0.018* | [0.01 0.09] | 0.498 |
| lS_calcarine | 1.615 (0.109) | 1.56 (0.12) | 2.438 | 0.023* | [0.01 0.1 ] | 0.482 |
| lG_cingul_Post_dorsal | 2.556 (0.136) | 2.493 (0.157) | 2.167 | 0.043* | [0.01 0.12] | 0.425 |
| lparahippocampal | 2.234 (0.228) | 2.152 (0.186) | 1.951 | 0.070 | [-0. 0.17] | 0.404 |
| lG_insular_short | 3.404 (0.319) | 3.25 (0.435) | 2.062 | 0.054 | [0.01 0.3 ] | 0.394 |
| risthmuscingulate | 2.028 (0.152) | 1.972 (0.141) | 1.894 | 0.078 | [-0. 0.11] | 0.384 |
| lS_pericallosal | 1.803 (0.134) | 1.757 (0.121) | 1.757 | 0.104 | [-0.01 0.1 ] | 0.358 |
| lG_cingul_Post_ventral | 2.108 (0.22) | 2.031 (0.224) | 1.743 | 0.106 | [-0.01 0.17] | 0.349 |
| lcaudalanteriorcingulate | 2.311 (0.178) | 2.25 (0.184) | 1.678 | 0.121 | [-0.01 0.13] | 0.335 |
| rfrontalpole | 2.194 (0.193) | 2.123 (0.231) | 1.672 | 0.121 | [-0.01 0.15] | 0.326 |
| lG_subcallosal | 2.712 (0.345) | 2.612 (0.345) | 1.451 | 0.181 | [-0.04 0.24] | 0.291 |
| rG_cingul_Post_ventral | 2.095 (0.193) | 2.038 (0.238) | 1.336 | 0.218 | [-0.03 0.14] | 0.259 |
| rentorhinal | 3.14 (0.265) | 3.059 (0.351) | 1.330 | 0.220 | [-0.04 0.2 ] | 0.255 |
| lLat_Fis_ant_Horizont | 2.305 (0.23) | 2.249 (0.246) | 1.179 | 0.280 | [-0.04 0.15] | 0.234 |
| lG_oc_temp_med_Parahip | 2.599 (0.248) | 2.548 (0.211) | 1.099 | 0.313 | [-0.04 0.14] | 0.226 |
| rcaudalanteriorcingulate | 2.281 (0.208) | 2.242 (0.161) | 1.009 | 0.356 | [-0.04 0.11] | 0.211 |
| rS_pericallosal | 1.728 (0.145) | 1.713 (0.116) | 0.556 | 0.632 | [-0.04 0.07] | 0.115 |
| lentorhinal | 3.129 (0.351) | 3.121 (0.334) | 0.115 | 0.932 | [-0.13 0.14] | 0.023 |
| **Extracted tract-based fractional anisotropy values** | | | | | | |
| SFO_R | 0.449 (0.065) | 0.423 (0.038) | 2.319 | 0.032* | [0 0.05] | 0.502 |
| BCC | 0.634 (0.041) | 0.614 (0.038) | 2.425 | 0.024* | [0 0.04] | 0.492 |
| FX_ST_R | 0.534 (0.045) | 0.515 (0.038) | 2.237 | 0.037* | [0 0.04] | 0.461 |
| CGH_R | 0.5 (0.055) | 0.479 (0.052) | 1.962 | 0.068 | [0 0.04] | 0.397 |
| CGH | 0.495 (0.057) | 0.474 (0.051) | 1.915 | 0.075 | [0 0.04] | 0.391 |
| SFO | 0.449 (0.062) | 0.431 (0.039) | 1.665 | 0.124 | [0 0.04] | 0.358 |
| IC_L | 0.581 (0.043) | 0.594 (0.032) | -1.664 | 0.124 | [-0.03 0] | 0.349 |
| EC_L | 0.424 (0.043) | 0.437 (0.038) | -1.639 | 0.128 | [-0.03 0] | 0.335 |
| CGH_L | 0.491 (0.065) | 0.469 (0.062) | 1.651 | 0.125 | [-0. 0.05] | 0.334 |
| EC | 0.428 (0.039) | 0.439 (0.031) | -1.501 | 0.166 | [-0.03 0] | 0.312 |
| ALIC_L | 0.527 (0.045) | 0.539 (0.039) | -1.412 | 0.193 | [-0.03 0] | 0.291 |
| CC | 0.639 (0.038) | 0.629 (0.031) | 1.339 | 0.218 | [-0. 0.02] | 0.277 |
| IC | 0.578 (0.04) | 0.587 (0.029) | -1.229 | 0.261 | [-0.02 0.01] | 0.259 |
| PLIC_L | 0.656 (0.054) | 0.668 (0.037) | -1.217 | 0.265 | [-0.03 0.01] | 0.259 |
| ALIC | 0.53 (0.045) | 0.54 (0.036) | -1.244 | 0.255 | [-0.03 0.01] | 0.258 |
| RLIC_L | 0.546 (0.044) | 0.556 (0.037) | -1.258 | 0.249 | [-0.03 0.01] | 0.258 |
| EC_R | 0.432 (0.039) | 0.44 (0.027) | -1.154 | 0.290 | [-0.02 0.01] | 0.244 |
| SS_L | 0.524 (0.038) | 0.534 (0.044) | -1.190 | 0.275 | [-0.03 0.01] | 0.234 |
| CST_R | 0.532 (0.053) | 0.519 (0.059) | 1.165 | 0.285 | [-0.01 0.04] | 0.230 |
| SS | 0.523 (0.035) | 0.531 (0.035) | -1.148 | 0.291 | [-0.02 0.01] | 0.230 |
| FX | 0.445 (0.084) | 0.424 (0.097) | 1.157 | 0.288 | [-0.01 0.06] | 0.227 |
| FXST | 0.526 (0.043) | 0.517 (0.039) | 1.097 | 0.313 | [-0.01 0.03] | 0.223 |
| RLIC | 0.541 (0.037) | 0.548 (0.031) | -1.016 | 0.353 | [-0.02 0.01] | 0.210 |
| ACR_R | 0.396 (0.037) | 0.39 (0.029) | 0.915 | 0.408 | [-0.01 0.02] | 0.191 |
| ALIC_R | 0.533 (0.046) | 0.541 (0.037) | -0.897 | 0.416 | [-0.02 0.01] | 0.186 |
| SS_R | 0.521 (0.034) | 0.527 (0.032) | -0.904 | 0.413 | [-0.02 0.01] | 0.183 |
| PLIC | 0.652 (0.049) | 0.658 (0.034) | -0.770 | 0.495 | [-0.02 0.01] | 0.163 |
| PCR_L | 0.424 (0.032) | 0.429 (0.033) | -0.744 | 0.509 | [-0.02 0.01] | 0.149 |
| IC_R | 0.575 (0.038) | 0.58 (0.028) | -0.682 | 0.551 | [-0.02 0.01] | 0.143 |
| SFO_L | 0.449 (0.067) | 0.441 (0.05) | 0.670 | 0.558 | [-0.02 0.03] | 0.141 |
| CGC_R | 0.548 (0.074) | 0.54 (0.056) | 0.609 | 0.596 | [-0.02 0.04] | 0.128 |
| CST_L | 0.526 (0.065) | 0.534 (0.06) | -0.629 | 0.583 | [-0.03 0.02] | 0.128 |
| CGC_L | 0.577 (0.065) | 0.584 (0.057) | -0.576 | 0.619 | [-0.03 0.02] | 0.118 |
| RLIC_R | 0.536 (0.035) | 0.539 (0.03) | -0.538 | 0.641 | [-0.02 0.01] | 0.111 |
| UNC_L | 0.492 (0.069) | 0.499 (0.051) | -0.520 | 0.652 | [-0.03 0.02] | 0.109 |
| CR_R | 0.422 (0.033) | 0.418 (0.024) | 0.512 | 0.656 | [-0.01 0.01] | 0.107 |
| IFO_L | 0.483 (0.056) | 0.488 (0.047) | -0.519 | 0.652 | [-0.03 0.02] | 0.107 |
| ACR | 0.391 (0.036) | 0.388 (0.03) | 0.500 | 0.663 | [-0.01 0.02] | 0.103 |
| PCR | 0.43 (0.03) | 0.433 (0.029) | -0.456 | 0.695 | [-0.01 0.01] | 0.092 |
| IFO | 0.487 (0.052) | 0.491 (0.041) | -0.426 | 0.716 | [-0.02 0.01] | 0.089 |
| GCC | 0.584 (0.055) | 0.58 (0.054) | 0.408 | 0.728 | [-0.02 0.03] | 0.082 |
| PTR_L | 0.53 (0.036) | 0.533 (0.036) | -0.397 | 0.733 | [-0.02 0.01] | 0.080 |
| SCR_L | 0.449 (0.043) | 0.452 (0.04) | -0.397 | 0.733 | [-0.02 0.01] | 0.080 |
| CR_L | 0.416 (0.033) | 0.419 (0.031) | -0.352 | 0.766 | [-0.02 0.01] | 0.071 |
| SCR_R | 0.442 (0.042) | 0.439 (0.032) | 0.305 | 0.801 | [-0.01 0.02] | 0.064 |
| IFO_R | 0.49 (0.057) | 0.493 (0.048) | -0.274 | 0.824 | [-0.02 0.02] | 0.056 |
| PLIC_R | 0.647 (0.046) | 0.649 (0.036) | -0.265 | 0.829 | [-0.02 0.01] | 0.055 |
| PTR | 0.531 (0.033) | 0.532 (0.033) | -0.200 | 0.880 | [-0.01 0.01] | 0.040 |
| UNC | 0.507 (0.054) | 0.509 (0.041) | -0.192 | 0.882 | [-0.02 0.02] | 0.040 |
| UNC_R | 0.518 (0.053) | 0.516 (0.047) | 0.191 | 0.882 | [-0.02 0.02] | 0.039 |
| CST | 0.529 (0.052) | 0.527 (0.055) | 0.182 | 0.887 | [-0.02 0.02] | 0.036 |
| PCR_R | 0.436 (0.032) | 0.437 (0.031) | -0.156 | 0.906 | [-0.01 0.01] | 0.031 |
| SLF_L | 0.457 (0.036) | 0.457 (0.03) | -0.128 | 0.926 | [-0.01 0.01] | 0.026 |
| SCC | 0.689 (0.042) | 0.69 (0.039) | -0.124 | 0.927 | [-0.02 0.02] | 0.025 |
| SLF_R | 0.448 (0.034) | 0.447 (0.025) | 0.102 | 0.940 | [-0.01 0.01] | 0.022 |
| ACR_L | 0.386 (0.037) | 0.385 (0.035) | 0.092 | 0.946 | [-0.01 0.02] | 0.019 |
| SCR | 0.445 (0.041) | 0.445 (0.035) | -0.076 | 0.956 | [-0.02 0.01] | 0.016 |
| CR | 0.419 (0.032) | 0.418 (0.027) | 0.063 | 0.965 | [-0.01 0.01] | 0.013 |
| FX_ST_L | 0.519 (0.048) | 0.519 (0.048) | 0.059 | 0.965 | [-0.02 0.02] | 0.012 |
| AverageFA | 0.431 (0.028) | 0.431 (0.023) | -0.044 | 0.972 | [-0.01 0.01] | 0.009 |
| PTR_R | 0.532 (0.034) | 0.531 (0.036) | 0.045 | 0.972 | [-0.01 0.01] | 0.009 |
| SLF | 0.452 (0.034) | 0.452 (0.025) | -0.028 | 0.980 | [-0.01 0.01] | 0.006 |
| CGC | 0.563 (0.068) | 0.562 (0.054) | 0.024 | 0.981 | [-0.02 0.03] | 0.005 |

Results are reported as mean (standard deviations). Abbreviations: R, right; L, left; G, gyrus; S, sulcus; sup, superior; inf, inferior; med, medial; lat, lateral; temp, temporal; front, frontal; gm, grey matter; FA, fractional anisotropy; ACR, anterior corona radiata; ALIC, anterior limb of internal capsule; BCC, body of corpus callosum; CC, corpus callosum; CGC, cingulate gyrus; CGH, perihippocampal cingulum tract; CR, corona radiata; CST, cortico-spinal tract; EC, external capsule; FX, fornix; FXST, fornix - stria terminalis; GCC, genu of corpus callosum; IC, internal capsule; IFO, inferior fronto-occipital fasciculus; PCR, posterior corona radiata; PLIC, posterior limb of internal capsule; PTR, posterior thalamic radiation; RLIC, retrolenticular limb of the internal capsule; SCC, splenium of corpus callosum; SCR, superior corona radiata; SFO, superior fronto-occipital fasciculus; SS, sagittal stratum; UNC, uncinate fasciculus.
* p<0.05; **p<0.001

**Table S2. Demographic and clinical characteristics of the identified data-driven clusters derived from UMAP-reduced model.**

|  | **Cluster 1 (N=55)** | **Cluster 2 (N=47)** | **t / χ2** | **p** |
| --- | --- | --- | --- | --- |
| **age** | 50.58 (9.28) | 48.09 (10.73) | 1.26 | 0.211 |
| **sex** | 21 M, 34 F | 12 M, 35 F | 1.85 | 0.173 |
| **scanner** | 28 old, 27 new | 18 old, 29 new | 1.63 | 0.234 |
| **number of episodes** | 5.65 (5.82) | 5.68 (6.69) | -0.21 | 0.983 |
| **age of onset** | 33.98 (10.95) | 30.34 (12.51) | 1.57 | 0.12 |
| **duration of illness** | 16.6 (10.67) | 17.74 (10.39) | -0.55 | 0.586 |
| **education** | 12.07 (3.86) | 13.55 (3.65) | -1.98 | 0.051 |
| **pharmacological load** | 4.25 (2.11) | 4.91 (2.13) | -1.57 | 0.119 |
| **BMI** | 25.29 (4.49) | 24.91 (4.94) | 0.41 | 0.684 |
| **TRD diagnosis** | 43 no-TRD, 12 TRD | 28 no-TRD, 19 TRD | 4.15 | 0.042* |
| **HDRS-21 total score** | 23.33 (6.98) | 21.87 (7.2) | 1.03 | 0.304 |
| **CTQ total score^a^** | 44.03 (13.96) | 41.56 (14.2) | 0.7 | 0.486 |
| **CTQ physical abuse^a^** | 6.03 (2.25) | 6.29 (2.64) | -0.42 | 0.674 |
| **CTQ emotional neglect^a^** | 13.77 (5.18) | 12.79 (5.54) | 0.72 | 0.473 |
| **CTQ emotional abuse^a^** | 9.30 (4.25) | 8.91 (4.52) | 0.35 | 0.725 |
| **CTQ physical neglect^a^** | 8.40 (3.94) | 6.88 (2.93) | 1.76 | 0.083 |
| **CTQ sexual abuse^a^** | 5.87 (3.16) | 6.38 (3.64) | -0.6 | 0.549 |
| **CTQ minimization/denial^a^** | 8.10 (3.10) | 9.50 (3.08) | -1.81 | 0.075 |
| **BDI total score^a^** | 17.50 (8.77) | 15.62 (7.59) | 0.92 | 0.361 |
| **BDI Self-Esteem^a^** | 6.57 (1.54) | 5.21 (2.78) | 1.72 | 0.091 |
| **BDI Anergy^a^** | 5.43 (3.06) | 5.82 (2.54) | -0.56 | 0.579 |
| **BDI Dysphoria^a^** | 5.50 (3.32) | 5.09 (2.83) | 0.54 | 0.594 |

Results are reported as mean (standard deviations) for continuous variables, whereas sample sizes are reported for categorical variables. Abbreviations: BMI, body mass index; TRD, treatment-resistant depression; HDRS-21, 21-Hamilton Depression Rating Scale; CTQ, Childhood Trauma Questionnaire; BDI, Beck Depression Inventory.

^a^ subsample of 64 MDD patients with clinical scales (cluster 1: N=30; cluster 2: N=34). *p<0.05, **p<0.001**

**Table S3. T-test statistics and effect sizes of the neuroimaging features driving the separation between the two clusters derived from UMAP-reduced model. Results are sorted by Cohen’s d**

| **Features** | **Cluster 1** | **Cluster 2** | **T** | **pFDR** | **CI 95%** | **Cohen's d** |
| --- | --- | --- | --- | --- | --- | --- |
| **Grey matter volumes** | | | | | | |
| Right Thalamus Proper | 4.243 (0.643) | 3.59 (0.646) | 5.097 | <0.001** | [0.4 0.91] | 1.013 |
| Left Thalamus Proper | 4.64 (0.616) | 4.011 (0.66) | 4.949 | <0.001** | [0.38 0.88] | 0.988 |
| Right Precentral Gyrus | 7.202 (1.047) | 6.328 (0.928) | 4.468 | <0.001** | [0.49 1.26] | 0.879 |
| Right Supplementary Motor Cortex | 3.739 (0.489) | 3.342 (0.484) | 4.112 | <0.001** | [0.21 0.59] | 0.816 |
| Left Medial Frontal Cerebrum | 1.385 (0.217) | 1.222 (0.183) | 4.109 | <0.001** | [0.08 0.24] | 0.806 |
| Right Ventral Ventricle | 0.977 (0.086) | 0.902 (0.103) | 3.950 | <0.001** | [0.04 0.11] | 0.796 |
| Right Parahippocampus Gyrus | 2.433 (0.267) | 2.225 (0.272) | 3.892 | <0.001** | [0.1 0.31] | 0.774 |
| Left Parahippocampus Gyrus | 2.548 (0.249) | 2.354 (0.273) | 3.728 | <0.001** | [0.09 0.3 ] | 0.746 |
| Right Superior Frontal Gyrus | 10.556 (1.389) | 9.518 (1.409) | 3.733 | <0.001** | [0.49 1.59] | 0.742 |
| Right Postcentral Gyrus | 5.793 (0.988) | 5.145 (0.784) | 3.695 | <0.001** | [0.3 1. ] | 0.721 |
| Left Middle Cingulate Gyrus | 3.397 (0.408) | 3.144 (0.285) | 3.672 | <0.001** | [0.12 0.39] | 0.710 |
| Right Amygdala | 0.786 (0.089) | 0.726 (0.081) | 3.582 | <0.001** | [0.03 0.09] | 0.706 |
| Left Precentral Gyrus | 6.823 (1.142) | 6.048 (1.042) | 3.582 | <0.001** | [0.35 1.2 ] | 0.706 |
| Left Ventral Ventricle | 0.974 (0.099) | 0.903 (0.103) | 3.500 | 0.002* | [0.03 0.11] | 0.697 |
| Left Anterior Orbital Gyrus | 1.3 (0.15) | 1.197 (0.149) | 3.472 | 0.002* | [0.04 0.16] | 0.689 |
| Right Superior Occipital Gyrus | 2.436 (0.422) | 2.185 (0.29) | 3.534 | <0.001** | [0.11 0.39] | 0.682 |
| Right Superior Medial Frontal Gyrus | 6.214 (0.877) | 5.64 (0.841) | 3.367 | 0.002* | [0.24 0.91] | 0.667 |
| Right Posterior Cingulate Gyrus | 2.881 (0.353) | 2.659 (0.307) | 3.390 | 0.002* | [0.09 0.35] | 0.666 |
| Left Amygdala | 0.788 (0.097) | 0.728 (0.082) | 3.391 | 0.002* | [0.02 0.09] | 0.665 |
| Left Gyrus Rectus | 1.569 (0.212) | 1.437 (0.185) | 3.356 | 0.002* | [0.05 0.21] | 0.660 |
| Right Entorhinal Area | 1.714 (0.183) | 1.593 (0.185) | 3.297 | 0.003* | [0.05 0.19] | 0.656 |
| Left Medial Orbital Gyrus | 3.263 (0.341) | 3.054 (0.293) | 3.334 | 0.002* | [0.08 0.33] | 0.654 |
| Right Hippocampus | 3.089 (0.279) | 2.909 (0.285) | 3.218 | 0.003* | [0.07 0.29] | 0.640 |
| Left Medial Precentral Gyrus | 1.628 (0.216) | 1.471 (0.275) | 3.158 | 0.004* | [0.06 0.26] | 0.639 |
| Left Middle Occipital Gyrus | 4.253 (0.501) | 3.915 (0.563) | 3.180 | 0.004* | [0.13 0.55] | 0.637 |
| Right Middle Frontal Gyrus | 13.547 (1.881) | 12.379 (1.799) | 3.201 | 0.004* | [0.44 1.89] | 0.634 |
| Left Entorhinal Area | 1.628 (0.187) | 1.512 (0.182) | 3.188 | 0.004* | [0.04 0.19] | 0.632 |
| Right Precuneus | 7.6 (1.021) | 6.986 (0.938) | 3.163 | 0.004* | [0.23 1. ] | 0.624 |
| Right Planum Polare | 1.362 (0.19) | 1.25 (0.166) | 3.172 | 0.004* | [0.04 0.18] | 0.623 |
| Right Angular Gyrus | 7.321 (0.946) | 6.766 (0.828) | 3.158 | 0.004* | [0.21 0.9 ] | 0.621 |
| Right Superior Temporal Gyrus | 5.27 (0.733) | 4.843 (0.633) | 3.159 | 0.004* | [0.16 0.7 ] | 0.620 |
| Right Supramarginal Gyrus | 5.688 (0.825) | 5.175 (0.857) | 3.070 | 0.005* | [0.18 0.85] | 0.612 |
| Right Inferior Frontal Orbital Gyrus | 0.993 (0.191) | 0.892 (0.13) | 3.146 | 0.004* | [0.04 0.16] | 0.607 |
| Left Anterior Cingulate Gyrus | 3.604 (0.485) | 3.313 (0.473) | 3.054 | 0.005* | [0.1 0.48] | 0.605 |
| Left Planum Polare | 1.44 (0.199) | 1.327 (0.17) | 3.078 | 0.005* | [0.04 0.19] | 0.604 |
| Left Hippocampus | 2.918 (0.25) | 2.77 (0.251) | 2.985 | 0.006* | [0.05 0.25] | 0.593 |
| Right Middle Occipital Gyrus | 3.477 (0.57) | 3.165 (0.479) | 3.010 | 0.006* | [0.11 0.52] | 0.590 |
| Left Superior Frontal Gyrus | 10.449 (1.612) | 9.495 (1.637) | 2.957 | 0.007* | [0.31 1.6 ] | 0.588 |
| Left Frontal Pole | 2.584 (0.366) | 2.363 (0.388) | 2.943 | 0.007* | [0.07 0.37] | 0.587 |
| Right Anterior Orbital Gyrus | 1.414 (0.173) | 1.313 (0.173) | 2.950 | 0.007* | [0.03 0.17] | 0.586 |
| Left Middle Temporal Gyrus | 10.6 (1.201) | 9.948 (1.001) | 2.990 | 0.006* | [0.22 1.08] | 0.585 |
| Cerebellar Lobule Cerebellar Vermal Lobules I-V | 3.081 (0.411) | 2.855 (0.361) | 2.958 | 0.007* | [0.07 0.38] | 0.581 |
| Right Medial Frontal Cerebrum | 1.421 (0.234) | 1.297 (0.185) | 2.978 | 0.006* | [0.04 0.21] | 0.581 |
| Left Inferior Occipital Gyrus | 4.275 (0.672) | 3.93 (0.499) | 2.971 | 0.007* | [0.11 0.58] | 0.577 |
| Right Middle Temporal Gyrus | 10.938 (1.245) | 10.252 (1.151) | 2.886 | 0.008* | [0.21 1.16] | 0.570 |
| Left Superior Temporal Gyrus | 5.106 (0.617) | 4.779 (0.527) | 2.886 | 0.008* | [0.1 0.55] | 0.566 |
| Right Frontal Pole | 2.751 (0.38) | 2.541 (0.362) | 2.849 | 0.009* | [0.06 0.36] | 0.564 |
| Left Calcarine and Cerebrum | 1.955 (0.356) | 1.776 (0.27) | 2.890 | 0.008* | [0.06 0.3 ] | 0.562 |
| Left Middle Frontal Gyrus | 13.386 (1.858) | 12.393 (1.656) | 2.853 | 0.009* | [0.3 1.68] | 0.562 |
| Left Supplementary Motor Cortex | 3.796 (0.538) | 3.492 (0.545) | 2.826 | 0.009* | [0.09 0.52] | 0.562 |
| Left Angular Gyrus | 6.628 (0.927) | 6.15 (0.757) | 2.864 | 0.009* | [0.15 0.81] | 0.560 |
| Left Temporal Pole | 7.194 (0.869) | 6.741 (0.752) | 2.822 | 0.009* | [0.13 0.77] | 0.554 |
| Right Lateral Orbital Gyrus | 1.57 (0.231) | 1.449 (0.206) | 2.809 | 0.01* | [0.04 0.21] | 0.553 |
| Left Supramarginal Gyrus | 6.33 (0.899) | 5.862 (0.795) | 2.790 | 0.01* | [0.14 0.8 ] | 0.549 |
| Right Temporal | 1.451 (0.213) | 1.337 (0.199) | 2.769 | 0.011* | [0.03 0.19] | 0.547 |
| Left Superior Medial Frontal Gyrus | 4.805 (0.599) | 4.462 (0.666) | 2.711 | 0.013* | [0.09 0.59] | 0.543 |
| Left Superior Parietal Lobule | 7.228 (1.164) | 6.643 (0.975) | 2.762 | 0.011* | [0.16 1.01] | 0.541 |
| Right Fusiform Gyrus | 5.656 (0.746) | 5.289 (0.609) | 2.735 | 0.012* | [0.1 0.63] | 0.535 |
| Right Inferior Occipital Gyrus | 4.298 (0.607) | 4.007 (0.456) | 2.755 | 0.011* | [0.08 0.5 ] | 0.535 |
| Left Posterior Cingulate Gyrus | 3.267 (0.437) | 3.058 (0.348) | 2.698 | 0.013* | [0.06 0.36] | 0.526 |
| Right Posterior Orbital Gyrus | 2.014 (0.338) | 1.851 (0.273) | 2.691 | 0.013* | [0.04 0.28] | 0.526 |
| Left Inferior Temporal Gyrus | 8.92 (1.148) | 8.348 (1.017) | 2.664 | 0.014* | [0.15 1] | 0.524 |
| Left Posterior Insula | 1.781 (0.205) | 1.676 (0.197) | 2.637 | 0.015* | [0.03 0.18] | 0.522 |
| Right Middle Cingulate Gyrus | 3.363 (0.411) | 3.168 (0.341) | 2.623 | 0.016* | [0.05 0.34] | 0.513 |
| Left Fusiform Gyrus | 5.365 (0.624) | 5.054 (0.593) | 2.581 | 0.017* | [0.07 0.55] | 0.511 |
| Right Medial Postcentral Gyrus | 0.578 (0.135) | 0.513 (0.121) | 2.586 | 0.017* | [0.02 0.12] | 0.509 |
| Right Subcallosal Area | 0.979 (0.142) | 0.915 (0.112) | 2.569 | 0.018* | [0.01 0.11] | 0.501 |
| Right Temporal Transverse Gyrus | 0.935 (0.158) | 0.86 (0.146) | 2.469 | 0.023* | [0.01 0.13] | 0.488 |
| Left Parietal Operculum | 1.788 (0.298) | 1.65 (0.269) | 2.469 | 0.023* | [0.03 0.25] | 0.486 |
| Cerebellar Lobule Cerebellar Vermal Lobules VI-VII | 1.591 (0.284) | 1.459 (0.254) | 2.466 | 0.023* | [0.03 0.24] | 0.485 |
| Right Medial Orbital Gyrus | 3.185 (0.386) | 3.008 (0.338) | 2.465 | 0.023* | [0.03 0.32] | 0.484 |
| Left Central Operculum | 3.033 (0.39) | 2.845 (0.392) | 2.425 | 0.025* | [0.03 0.34] | 0.482 |
| Right Inferior Temporal Gyrus | 9.062 (1.127) | 8.542 (1.045) | 2.417 | 0.025* | [0.09 0.95] | 0.477 |
| Left Putamen | 2.68 (0.477) | 2.472 (0.382) | 2.438 | 0.024* | [0.04 0.38] | 0.476 |
| Right Central Operculum | 3.084 (0.415) | 2.891 (0.399) | 2.393 | 0.027* | [0.03 0.35] | 0.474 |
| Left Frontal Operculum | 1.411 (0.172) | 1.329 (0.176) | 2.375 | 0.028* | [0.01 0.15] | 0.473 |
| Left Medial Postcentral Gyrus | 0.631 (0.128) | 0.571 (0.132) | 2.333 | 0.031* | [0.01 0.11] | 0.465 |
| Left Lateral Orbital Gyrus | 1.694 (0.244) | 1.592 (0.19) | 2.368 | 0.029* | [0.02 0.19] | 0.461 |
| Right Lingual Gyrus | 5.218 (0.806) | 4.859 (0.746) | 2.331 | 0.031* | [0.05 0.66] | 0.460 |
| Left Basal Cerebrum and Forebrain Brain | 0.316 (0.05) | 0.296 (0.038) | 2.334 | 0.031* | [0 0.04] | 0.453 |
| Left Postcentral Gyrus | 6.3 (0.999) | 5.877 (0.856) | 2.300 | 0.033* | [0.06 0.79] | 0.451 |
| Left Temporal | 1.502 (0.244) | 1.395 (0.238) | 2.228 | 0.039* | [0.01 0.2 ] | 0.442 |
| Left Posterior Orbital Gyrus | 2.32 (0.356) | 2.174 (0.309) | 2.222 | 0.04* | [0.02 0.28] | 0.437 |
| Right Putamen | 2.575 (0.477) | 2.388 (0.395) | 2.164 | 0.046* | [0.02 0.36] | 0.424 |
| Right Medial Precentral Gyrus | 1.458 (0.286) | 1.342 (0.258) | 2.152 | 0.047* | [0.01 0.22] | 0.424 |
| Left Inferior Frontal Gyrus | 2.23 (0.319) | 2.101 (0.285) | 2.152 | 0.047* | [0.01 0.25] | 0.424 |
| Left Lingual Gyrus | 5.256 (0.779) | 4.939 (0.718) | 2.136 | 0.048* | [0.02 0.61] | 0.422 |
| Left Precuneus | 8.01 (1.026) | 7.61 (0.876) | 2.128 | 0.049* | [0.03 0.77] | 0.417 |
| Right Cuneus | 2.517 (0.473) | 2.338 (0.384) | 2.109 | 0.051 | [0.01 0.35] | 0.412 |
| Left Exterior Cerebellum | 32.583 (4.286) | 30.914 (3.927) | 2.051 | 0.057 | [0.05 3.28] | 0.405 |
| Right Superior Parietal Lobule | 6.986 (0.974) | 6.596 (0.965) | 2.028 | 0.060 | [0.01 0.77] | 0.403 |
| Right Temporal Pole | 7.186 (0.942) | 6.814 (0.908) | 2.025 | 0.061 | [0.01 0.74] | 0.401 |
| Left Triangular Inferior Frontal Gyrus | 2.513 (0.4) | 2.375 (0.269) | 2.074 | 0.055 | [0.01 0.27] | 0.400 |
| Right Calcarine and Cerebrum | 1.615 (0.321) | 1.503 (0.225) | 2.058 | 0.057 | [0 0.22] | 0.398 |
| Cerebellar Lobule Cerebellar Vermal Lobules VIII-X | 1.38 (0.194) | 1.305 (0.183) | 2.011 | 0.062 | [0 0.15] | 0.397 |
| Right Occipital Fusiform Gyrus | 2.446 (0.391) | 2.302 (0.344) | 1.984 | 0.066 | [0 0.29] | 0.390 |
| Left Subcallosal Area | 1.026 (0.155) | 0.972 (0.118) | 2.007 | 0.063 | [0 0.11] | 0.390 |
| Right Gyrus Rectus | 1.682 (0.257) | 1.588 (0.231) | 1.960 | 0.069 | [0 0.19] | 0.386 |
| Left Occipital Fusiform Gyrus | 2.584 (0.425) | 2.437 (0.358) | 1.892 | 0.079 | [-0.01 0.3 ] | 0.371 |
| Left Cuneus | 2.738 (0.519) | 2.563 (0.412) | 1.893 | 0.079 | [-0.01 0.36] | 0.369 |
| Right Posterior Insula | 1.922 (0.232) | 1.841 (0.225) | 1.777 | 0.101 | [-0.01 0.17] | 0.352 |
| Right Exterior Cerebellum | 32.346 (4.309) | 30.944 (3.711) | 1.765 | 0.103 | [-0.17 2.98] | 0.347 |
| Left Occipital Pole | 1.784 (0.445) | 1.649 (0.314) | 1.781 | 0.100 | [-0.02 0.28] | 0.344 |
| Right Anterior Cingulate Gyrus | 2.96 (0.446) | 2.812 (0.434) | 1.696 | 0.118 | [-0.03 0.32] | 0.336 |
| Left Temporal Transverse Gyrus | 0.92 (0.165) | 0.865 (0.169) | 1.654 | 0.128 | [-0.01 0.12] | 0.329 |
| Right Basal Cerebrum and Forebrain Brain | 0.314 (0.047) | 0.301 (0.035) | 1.635 | 0.132 | [0 0.03] | 0.317 |
| Left Caudate | 2.296 (0.293) | 2.19 (0.377) | 1.553 | 0.155 | [-0.03 0.24] | 0.315 |
| Left Inferior Frontal Orbital Gyrus | 1.202 (0.204) | 1.142 (0.174) | 1.582 | 0.146 | [-0.02 0.13] | 0.310 |
| Right Caudate | 2.634 (0.337) | 2.527 (0.367) | 1.525 | 0.162 | [-0.03 0.25] | 0.305 |
| Left Anterior Insula | 3.495 (0.362) | 3.377 (0.433) | 1.474 | 0.177 | [-0.04 0.28] | 0.297 |
| Right Anterior Insula | 3.411 (0.386) | 3.297 (0.388) | 1.491 | 0.172 | [-0.04 0.27] | 0.296 |
| Left Superior Occipital Gyrus | 1.996 (0.372) | 1.899 (0.296) | 1.463 | 0.179 | [-0.03 0.23] | 0.286 |
| Left Pallidum | 0.251 (0.101) | 0.227 (0.077) | 1.393 | 0.201 | [-0.01 0.06] | 0.271 |
| Right Parietal Operculum | 1.476 (0.259) | 1.417 (0.259) | 1.150 | 0.298 | [-0.04 0.16] | 0.229 |
| Right Frontal Operculum | 1.444 (0.202) | 1.406 (0.183) | 0.999 | 0.363 | [-0.04 0.11] | 0.197 |
| Right Inferior Frontal Gyrus | 2.342 (0.33) | 2.278 (0.333) | 0.962 | 0.381 | [-0.07 0.19] | 0.191 |
| Right Occipital Pole | 1.646 (0.374) | 1.583 (0.313) | 0.935 | 0.392 | [-0.07 0.2 ] | 0.183 |
| Right Triangular Inferior Frontal Gyrus | 2.449 (0.39) | 2.391 (0.325) | 0.815 | 0.458 | [-0.08 0.2 ] | 0.160 |
| Left Accumbens | 0.341 (0.049) | 0.335 (0.044) | 0.691 | 0.533 | [-0.01 0.02] | 0.136 |
| Right Accumbens | 0.323 (0.044) | 0.317 (0.043) | 0.604 | 0.587 | [-0.01 0.02] | 0.120 |
| Right Pallidum | 0.233 (0.096) | 0.225 (0.101) | 0.376 | 0.739 | [-0.03 0.05] | 0.075 |
| Optic Chiasm | 0.117 (0.023) | 0.117 (0.019) | -0.025 | 0.980 | [-0.01 0.01] | 0.005 |
| **Cortical thickness** | | | | | | |
| rinferiorparietal | 2.231 (0.11) | 2.019 (0.124) | 9.080 | <0.001** | [0.17 0.26] | 1.821 |
| rsuperiorparietal | 1.943 (0.132) | 1.704 (0.144) | 8.674 | <0.001** | [0.18 0.29] | 1.735 |
| rS_intrapariet_and_P_trans | 2.018 (0.137) | 1.767 (0.155) | 8.594 | <0.001** | [0.19 0.31] | 1.724 |
| rS_front_sup | 2.317 (0.145) | 2.024 (0.197) | 8.407 | <0.001** | [0.22 0.36] | 1.710 |
| rsupramarginal | 2.27 (0.117) | 2.055 (0.138) | 8.438 | <0.001** | [0.16 0.27] | 1.698 |
| lsupramarginal | 2.274 (0.108) | 2.079 (0.127) | 8.304 | <0.001** | [0.15 0.24] | 1.671 |
| rS_postcentral | 1.885 (0.14) | 1.638 (0.157) | 8.328 | <0.001** | [0.19 0.31] | 1.669 |
| linferiorparietal | 2.267 (0.121) | 2.07 (0.117) | 8.323 | <0.001** | [0.15 0.24] | 1.649 |
| rsuperiorfrontal | 2.401 (0.142) | 2.135 (0.183) | 8.117 | <0.001** | [0.2 0.33] | 1.645 |
| rS_temporal_sup | 2.359 (0.101) | 2.182 (0.122) | 7.887 | <0.001** | [0.13 0.22] | 1.589 |
| lrostralmiddlefrontal | 2.2 (0.114) | 2.009 (0.13) | 7.824 | <0.001** | [0.14 0.24] | 1.570 |
| rmiddletemporal | 2.551 (0.13) | 2.345 (0.133) | 7.841 | <0.001** | [0.15 0.26] | 1.560 |
| lsuperiorparietal | 1.919 (0.127) | 1.712 (0.139) | 7.793 | <0.001** | [0.15 0.26] | 1.559 |
| rG_front_sup | 2.281 (0.167) | 1.991 (0.206) | 7.719 | <0.001** | [0.22 0.36] | 1.558 |
| lG_pariet_inf_Angular | 2.358 (0.145) | 2.13 (0.152) | 7.697 | <0.001** | [0.17 0.29] | 1.535 |
| rG_pariet_inf_Supramar | 2.331 (0.136) | 2.109 (0.154) | 7.642 | <0.001** | [0.16 0.28] | 1.533 |
| rcaudalmiddlefrontal | 2.235 (0.161) | 1.97 (0.191) | 7.518 | <0.001** | [0.2 0.34] | 1.513 |
| lS_intrapariet_and_P_trans | 1.992 (0.137) | 1.78 (0.145) | 7.544 | <0.001** | [0.16 0.27] | 1.505 |
| rG_temporal_middle | 2.599 (0.154) | 2.376 (0.146) | 7.505 | <0.001** | [0.16 0.28] | 1.484 |
| lG_pariet_inf_Supramar | 2.358 (0.133) | 2.158 (0.138) | 7.413 | <0.001** | [0.15 0.25] | 1.477 |
| lG_front_middle | 2.271 (0.162) | 2.021 (0.18) | 7.328 | <0.001** | [0.18 0.32] | 1.468 |
| lcaudalmiddlefrontal | 2.23 (0.186) | 1.944 (0.203) | 7.338 | <0.001** | [0.21 0.36] | 1.468 |
| lS_front_inf | 2.102 (0.142) | 1.9 (0.133) | 7.378 | <0.001** | [0.15 0.26] | 1.458 |
| rG_occipital_middle | 2.128 (0.129) | 1.924 (0.154) | 7.192 | <0.001** | [0.15 0.26] | 1.448 |
| lS_front_sup | 2.317 (0.16) | 2.059 (0.2) | 7.109 | <0.001** | [0.19 0.33] | 1.437 |
| rG_pariet_inf_Angular | 2.274 (0.151) | 2.037 (0.181) | 7.113 | <0.001** | [0.17 0.3 ] | 1.433 |
| lsuperiorfrontal | 2.295 (0.163) | 2.044 (0.193) | 7.014 | <0.001** | [0.18 0.32] | 1.412 |
| lG_parietal_sup | 2.035 (0.156) | 1.805 (0.172) | 7.045 | <0.001** | [0.17 0.3 ] | 1.410 |
| lS_precentral_inf_part | 2.13 (0.156) | 1.897 (0.178) | 6.978 | <0.001** | [0.17 0.3 ] | 1.400 |
| rS_subparietal | 2.145 (0.13) | 1.968 (0.124) | 7.048 | <0.001** | [0.13 0.23] | 1.395 |
| rS_cingul_Marginalis | 2.013 (0.133) | 1.831 (0.13) | 6.973 | <0.001** | [0.13 0.23] | 1.383 |
| rfusiform | 2.217 (0.149) | 2.035 (0.11) | 7.105 | <0.001** | [0.13 0.23] | 1.379 |
| lS_postcentral | 1.865 (0.138) | 1.668 (0.15) | 6.877 | <0.001** | [0.14 0.25] | 1.376 |
| lS_front_middle | 2.189 (0.117) | 2.008 (0.147) | 6.774 | <0.001** | [0.13 0.23] | 1.370 |
| rG_parietal_sup | 2.056 (0.186) | 1.789 (0.208) | 6.787 | <0.001** | [0.19 0.34] | 1.360 |
| lprecuneus | 2.205 (0.112) | 2.05 (0.116) | 6.824 | <0.001** | [0.11 0.2 ] | 1.359 |
| rG_precuneus | 2.098 (0.151) | 1.87 (0.186) | 6.723 | <0.001** | [0.16 0.3 ] | 1.357 |
| rprecuneus | 2.044 (0.131) | 1.86 (0.144) | 6.736 | <0.001** | [0.13 0.24] | 1.349 |
| lG_precuneus | 2.315 (0.138) | 2.133 (0.133) | 6.750 | <0.001** | [0.13 0.23] | 1.337 |
| llateralorbitofrontal | 2.571 (0.146) | 2.385 (0.135) | 6.708 | <0.001** | [0.13 0.24] | 1.324 |
| rS_precentral_inf_part | 2.246 (0.139) | 2.044 (0.167) | 6.557 | <0.001** | [0.14 0.26] | 1.322 |
| lG_and_S_subcentral | 2.274 (0.152) | 2.058 (0.177) | 6.575 | <0.001** | [0.15 0.28] | 1.321 |
| rlateraloccipital | 1.8 (0.097) | 1.671 (0.101) | 6.555 | <0.001** | [0.09 0.17] | 1.306 |
| lG_temp_sup_Plan_tempo | 2.256 (0.151) | 2.052 (0.162) | 6.527 | <0.001** | [0.14 0.27] | 1.303 |
| lS_temporal_sup | 2.3 (0.115) | 2.148 (0.119) | 6.545 | <0.001** | [0.11 0.2 ] | 1.303 |
| lG_front_sup | 2.209 (0.194) | 1.939 (0.222) | 6.481 | <0.001** | [0.19 0.35] | 1.301 |
| rlateralorbitofrontal | 2.358 (0.135) | 2.192 (0.121) | 6.552 | <0.001** | [0.12 0.22] | 1.290 |
| rG_front_middle | 2.273 (0.144) | 2.072 (0.171) | 6.373 | <0.001** | [0.14 0.26] | 1.283 |
| rS_oc_sup_and_transversal | 1.894 (0.157) | 1.71 (0.128) | 6.514 | <0.001** | [0.13 0.24] | 1.274 |
| lfusiform | 2.23 (0.134) | 2.072 (0.117) | 6.373 | <0.001** | [0.11 0.21] | 1.252 |
| rG_orbital | 2.311 (0.168) | 2.118 (0.14) | 6.339 | <0.001** | [0.13 0.25] | 1.241 |
| rS_precentral_sup_part | 1.973 (0.237) | 1.684 (0.228) | 6.265 | <0.001** | [0.2 0.38] | 1.241 |
| lsuperiortemporal | 2.451 (0.149) | 2.262 (0.156) | 6.212 | <0.001** | [0.13 0.25] | 1.239 |
| rrostralmiddlefrontal | 2.178 (0.107) | 2.027 (0.138) | 6.089 | <0.001** | [0.1 0.2] | 1.234 |
| linferiortemporal | 2.45 (0.129) | 2.292 (0.127) | 6.216 | <0.001** | [0.11 0.21] | 1.233 |
| lG_temporal_inf | 2.497 (0.155) | 2.313 (0.147) | 6.138 | <0.001** | [0.12 0.24] | 1.214 |
| lparsopercularis | 2.395 (0.145) | 2.214 (0.157) | 6.025 | <0.001** | [0.12 0.24] | 1.204 |
| lG_and_S_cingul_Mid_Post | 2.282 (0.12) | 2.124 (0.146) | 5.948 | <0.001** | [0.11 0.21] | 1.200 |
| rposteriorcingulate | 2.127 (0.11) | 1.991 (0.117) | 6.010 | <0.001** | [0.09 0.18] | 1.200 |
| rG_front_inf_Opercular | 2.527 (0.176) | 2.314 (0.184) | 5.941 | <0.001** | [0.14 0.28] | 1.185 |
| rsuperiortemporal | 2.537 (0.166) | 2.335 (0.179) | 5.866 | <0.001** | [0.13 0.27] | 1.172 |
| rprecentral | 1.652 (0.125) | 1.493 (0.148) | 5.816 | <0.001** | [0.1 0.21] | 1.171 |
| rG_temp_sup_Plan_tempo | 2.283 (0.186) | 2.06 (0.197) | 5.860 | <0.001** | [0.15 0.3 ] | 1.169 |
| rS_interm_prim_Jensen | 2.159 (0.157) | 1.972 (0.166) | 5.803 | <0.001** | [0.12 0.25] | 1.158 |
| rparsopercularis | 2.37 (0.137) | 2.189 (0.177) | 5.712 | <0.001** | [0.12 0.24] | 1.157 |
| rG_and_S_cingul_Mid_Post | 2.232 (0.15) | 2.061 (0.148) | 5.764 | <0.001** | [0.11 0.23] | 1.144 |
| lS_circular_insula_sup | 2.668 (0.14) | 2.51 (0.137) | 5.742 | <0.001** | [0.1 0.21] | 1.138 |
| rS_temporal_inf | 2.258 (0.132) | 2.106 (0.134) | 5.718 | <0.001** | [0.1 0.2] | 1.138 |
| lG_orbital | 2.524 (0.181) | 2.325 (0.167) | 5.753 | <0.001** | [0.13 0.27] | 1.135 |
| lG_and_S_frontomargin | 2.135 (0.124) | 1.986 (0.143) | 5.582 | <0.001** | [0.1 0.2] | 1.122 |
| lG_and_S_transv_frontopol | 2.29 (0.176) | 2.092 (0.18) | 5.598 | <0.001** | [0.13 0.27] | 1.114 |
| lS_cingul_Marginalis | 2.119 (0.125) | 1.965 (0.153) | 5.510 | <0.001** | [0.1 0.21] | 1.112 |
| lG_front_inf_Opercular | 2.452 (0.16) | 2.25 (0.206) | 5.464 | <0.001** | [0.13 0.28] | 1.107 |
| rG_and_S_subcentral | 2.296 (0.187) | 2.092 (0.184) | 5.578 | <0.001** | [0.13 0.28] | 1.106 |
| rpostcentral | 1.547 (0.13) | 1.412 (0.113) | 5.620 | <0.001** | [0.09 0.18] | 1.104 |
| rparsorbitalis | 2.349 (0.193) | 2.138 (0.19) | 5.549 | <0.001** | [0.14 0.29] | 1.101 |
| lG_and_S_cingul_Ant | 2.469 (0.122) | 2.32 (0.15) | 5.440 | <0.001** | [0.09 0.2 ] | 1.098 |
| rbankssts | 2.347 (0.145) | 2.177 (0.167) | 5.464 | <0.001** | [0.11 0.23] | 1.098 |
| lparstriangularis | 2.293 (0.13) | 2.148 (0.136) | 5.509 | <0.001** | [0.09 0.2 ] | 1.098 |
| rparstriangularis | 2.272 (0.146) | 2.109 (0.151) | 5.509 | <0.001** | [0.1 0.22] | 1.098 |
| rS_orbital_lateral | 2.169 (0.149) | 2.001 (0.158) | 5.496 | <0.001** | [0.11 0.23] | 1.097 |
| linsula | 3.019 (0.188) | 2.812 (0.196) | 5.433 | <0.001** | [0.13 0.28] | 1.083 |
| rG_temp_sup_Lateral | 2.606 (0.2) | 2.389 (0.204) | 5.418 | <0.001** | [0.14 0.3 ] | 1.078 |
| lS_orbital_H_Shaped | 2.479 (0.135) | 2.32 (0.163) | 5.342 | <0.001** | [0.1 0.22] | 1.077 |
| lS_parieto_occipital | 1.964 (0.156) | 1.796 (0.155) | 5.422 | <0.001** | [0.11 0.23] | 1.076 |
| lLat_Fis_post | 2.164 (0.15) | 2.006 (0.145) | 5.403 | <0.001** | [0.1 0.22] | 1.071 |
| rS_oc_temp_med_and_Lingual | 2.041 (0.149) | 1.891 (0.132) | 5.379 | <0.001** | [0.09 0.2 ] | 1.058 |
| lG_temp_sup_Lateral | 2.547 (0.172) | 2.352 (0.199) | 5.253 | <0.001** | [0.12 0.27] | 1.055 |
| rPole_temporal | 2.777 (0.263) | 2.51 (0.243) | 5.322 | <0.001** | [0.17 0.37] | 1.051 |
| lprecentral | 1.6 (0.15) | 1.437 (0.161) | 5.239 | <0.001** | [0.1 0.22] | 1.047 |
| rS_occipital_ant | 2.098 (0.156) | 1.93 (0.165) | 5.245 | <0.001** | [0.1 0.23] | 1.046 |
| lG_front_inf_Triangul | 2.256 (0.158) | 2.073 (0.193) | 5.172 | <0.001** | [0.11 0.25] | 1.044 |
| rG_and_S_transv_frontopol | 2.199 (0.134) | 2.046 (0.161) | 5.177 | <0.001** | [0.09 0.21] | 1.043 |
| lparacentral | 1.729 (0.17) | 1.548 (0.178) | 5.216 | <0.001** | [0.11 0.25] | 1.040 |
| lpostcentral | 1.535 (0.12) | 1.416 (0.11) | 5.268 | <0.001** | [0.07 0.16] | 1.039 |
| lG_and_S_cingul_Mid_Ant | 2.457 (0.162) | 2.284 (0.177) | 5.149 | <0.001** | [0.11 0.24] | 1.030 |
| rG_oc_temp_lat_fusifor | 2.061 (0.173) | 1.896 (0.145) | 5.254 | <0.001** | [0.1 0.23] | 1.030 |
| rG_and_S_frontomargin | 2.119 (0.109) | 1.978 (0.164) | 4.981 | <0.001** | [0.08 0.2 ] | 1.020 |
| lG_temp_sup_Plan_polar | 3.22 (0.265) | 2.923 (0.323) | 5.041 | <0.001** | [0.18 0.42] | 1.017 |
| rG_front_inf_Triangul | 2.299 (0.178) | 2.124 (0.167) | 5.108 | <0.001** | [0.11 0.24] | 1.010 |
| lS_precentral_sup_part | 1.82 (0.243) | 1.588 (0.213) | 5.137 | <0.001** | [0.14 0.32] | 1.010 |
| lfrontalpole | 2.301 (0.228) | 2.062 (0.248) | 5.030 | <0.001** | [0.14 0.33] | 1.006 |
| rS_circular_insula_sup | 2.701 (0.154) | 2.551 (0.144) | 5.088 | <0.001** | [0.09 0.21] | 1.005 |
| rS_collat_transv_ant | 2.512 (0.216) | 2.323 (0.149) | 5.203 | <0.001** | [0.12 0.26] | 1.005 |
| rG_precentral | 1.594 (0.159) | 1.412 (0.205) | 4.941 | <0.001** | [0.11 0.26] | 1.001 |
| llateraloccipital | 1.8 (0.119) | 1.684 (0.111) | 5.061 | <0.001** | [0.07 0.16] | 1.000 |
| lS_occipital_ant | 2.06 (0.194) | 1.877 (0.171) | 5.045 | <0.001** | [0.11 0.25] | 0.992 |
| rG_occipital_sup | 1.67 (0.171) | 1.503 (0.166) | 4.985 | <0.001** | [0.1 0.23] | 0.988 |
| lG_temporal_middle | 2.544 (0.156) | 2.392 (0.153) | 4.969 | <0.001** | [0.09 0.21] | 0.986 |
| lmiddletemporal | 2.498 (0.14) | 2.358 (0.145) | 4.939 | <0.001** | [0.08 0.2 ] | 0.984 |
| rS_oc_temp_lat | 2.301 (0.164) | 2.137 (0.169) | 4.928 | <0.001** | [0.1 0.23] | 0.981 |
| rS_front_inf | 2.019 (0.133) | 1.885 (0.141) | 4.891 | <0.001** | [0.08 0.19] | 0.976 |
| rS_oc_middle_and_Lunatus | 1.925 (0.144) | 1.777 (0.16) | 4.865 | <0.001** | [0.09 0.21] | 0.975 |
| lmedialorbitofrontal | 2.235 (0.141) | 2.102 (0.131) | 4.931 | <0.001** | [0.08 0.19] | 0.974 |
| rG_temp_sup_G_T_transv | 2.057 (0.292) | 1.776 (0.286) | 4.902 | <0.001** | [0.17 0.39] | 0.972 |
| lposteriorcingulate | 2.163 (0.088) | 2.067 (0.111) | 4.795 | <0.001** | [0.06 0.14] | 0.970 |
| lS_oc_temp_med_and_Lingual | 2.089 (0.146) | 1.946 (0.152) | 4.851 | <0.001** | [0.08 0.2 ] | 0.967 |
| lS_temporal_inf | 2.264 (0.13) | 2.137 (0.132) | 4.862 | <0.001** | [0.07 0.18] | 0.967 |
| rG_and_S_cingul_Mid_Ant | 2.526 (0.157) | 2.377 (0.156) | 4.804 | <0.001** | [0.09 0.21] | 0.954 |
| rS_orbital_H_Shaped | 2.409 (0.15) | 2.268 (0.151) | 4.734 | <0.001** | [0.08 0.2 ] | 0.941 |
| lS_interm_prim_Jensen | 2.306 (0.193) | 2.121 (0.202) | 4.716 | <0.001** | [0.11 0.26] | 0.940 |
| rS_front_middle | 2.181 (0.125) | 2.044 (0.168) | 4.618 | <0.001** | [0.08 0.2 ] | 0.938 |
| rG_front_inf_Orbital | 2.257 (0.254) | 2.035 (0.215) | 4.781 | <0.001** | [0.13 0.31] | 0.937 |
| rinferiortemporal | 2.399 (0.135) | 2.276 (0.13) | 4.669 | <0.001** | [0.07 0.17] | 0.925 |
| rmedialorbitofrontal | 2.322 (0.116) | 2.21 (0.128) | 4.609 | <0.001** | [0.06 0.16] | 0.923 |
| rparacentral | 1.558 (0.152) | 1.424 (0.137) | 4.687 | <0.001** | [0.08 0.19] | 0.923 |
| lPole_temporal | 3.001 (0.28) | 2.751 (0.264) | 4.650 | <0.001** | [0.14 0.36] | 0.919 |
| rG_and_S_cingul_Ant | 2.533 (0.138) | 2.401 (0.151) | 4.559 | <0.001** | [0.07 0.19] | 0.912 |
| lG_oc_temp_lat_fusifor | 2.052 (0.152) | 1.916 (0.145) | 4.597 | <0.001** | [0.08 0.19] | 0.910 |
| rtransversetemporal | 1.986 (0.241) | 1.764 (0.25) | 4.549 | <0.001** | [0.13 0.32] | 0.906 |
| lS_subparietal | 2.225 (0.111) | 2.123 (0.116) | 4.527 | <0.001** | [0.06 0.15] | 0.902 |
| lS_collat_transv_post | 1.734 (0.202) | 1.575 (0.141) | 4.650 | <0.001** | [0.09 0.23] | 0.899 |
| rLat_Fis_post | 2.392 (0.165) | 2.242 (0.169) | 4.512 | <0.001** | [0.08 0.22] | 0.898 |
| lS_oc_sup_and_transversal | 1.895 (0.147) | 1.764 (0.146) | 4.500 | <0.001** | [0.07 0.19] | 0.893 |
| lG_and_S_occipital_inf | 2.029 (0.155) | 1.894 (0.147) | 4.506 | <0.001** | [0.08 0.19] | 0.892 |
| rG_cingul_Post_dorsal | 2.509 (0.165) | 2.361 (0.17) | 4.465 | <0.001** | [0.08 0.21] | 0.889 |
| lcuneus | 1.564 (0.156) | 1.432 (0.139) | 4.515 | <0.001** | [0.07 0.19] | 0.889 |
| lG_rectus | 2.22 (0.154) | 2.085 (0.151) | 4.463 | <0.001** | [0.08 0.2 ] | 0.885 |
| lG_cuneus | 1.487 (0.134) | 1.368 (0.137) | 4.418 | <0.001** | [0.07 0.17] | 0.879 |
| rG_and_S_occipital_inf | 2.133 (0.156) | 2.001 (0.15) | 4.365 | <0.001** | [0.07 0.19] | 0.864 |
| rinsula | 3.013 (0.196) | 2.846 (0.191) | 4.347 | <0.001** | [0.09 0.24] | 0.862 |
| lbankssts | 2.286 (0.147) | 2.152 (0.166) | 4.287 | <0.001** | [0.07 0.2 ] | 0.860 |
| ltemporalpole | 3.348 (0.349) | 3.022 (0.412) | 4.273 | <0.001** | [0.17 0.48] | 0.860 |
| lG_occipital_middle | 2.146 (0.148) | 2.016 (0.158) | 4.274 | <0.001** | [0.07 0.19] | 0.853 |
| lS_orbital_lateral | 2.111 (0.14) | 1.976 (0.179) | 4.210 | <0.001** | [0.07 0.2 ] | 0.852 |
| lS_collat_transv_ant | 2.539 (0.185) | 2.384 (0.18) | 4.293 | <0.001** | [0.08 0.23] | 0.851 |
| lS_circular_insula_inf | 2.838 (0.188) | 2.679 (0.193) | 4.182 | <0.001** | [0.08 0.23] | 0.832 |
| lS_oc_middle_and_Lunatus | 1.903 (0.135) | 1.789 (0.145) | 4.083 | <0.001** | [0.06 0.17] | 0.815 |
| rG_oc_temp_med_Parahip | 2.717 (0.206) | 2.551 (0.206) | 4.056 | <0.001** | [0.08 0.25] | 0.806 |
| rG_insular_short | 3.287 (0.326) | 3.035 (0.298) | 4.076 | <0.001** | [0.13 0.37] | 0.804 |
| lG_occipital_sup | 1.641 (0.176) | 1.502 (0.169) | 4.038 | <0.001** | [0.07 0.21] | 0.799 |
| rparahippocampal | 2.285 (0.234) | 2.112 (0.193) | 4.084 | <0.001** | [0.09 0.26] | 0.799 |
| rS_collat_transv_post | 1.662 (0.173) | 1.527 (0.167) | 4.000 | <0.001** | [0.07 0.2 ] | 0.792 |
| rG_postcentral | 1.341 (0.169) | 1.215 (0.147) | 4.020 | <0.001** | [0.06 0.19] | 0.790 |
| lS_oc_temp_lat | 2.271 (0.17) | 2.133 (0.181) | 3.928 | <0.001** | [0.07 0.21] | 0.784 |
| rrostralanteriorcingulate | 2.608 (0.163) | 2.487 (0.141) | 3.985 | <0.001** | [0.06 0.18] | 0.783 |
| rLat_Fis_ant_Horizont | 2.185 (0.201) | 2.029 (0.2) | 3.906 | <0.001** | [0.08 0.23] | 0.776 |
| lG_precentral | 1.495 (0.198) | 1.331 (0.226) | 3.863 | <0.001** | [0.08 0.25] | 0.775 |
| lG_Ins_lg_and_S_cent_ins | 2.975 (0.264) | 2.751 (0.318) | 3.837 | <0.001** | [0.11 0.34] | 0.773 |
| rLat_Fis_ant_Vertical | 2.331 (0.195) | 2.174 (0.218) | 3.803 | <0.001** | [0.07 0.24] | 0.762 |
| lG_oc_temp_med_Lingual | 1.609 (0.126) | 1.514 (0.123) | 3.829 | <0.001** | [0.05 0.14] | 0.759 |
| llingual | 1.693 (0.113) | 1.61 (0.106) | 3.833 | <0.001** | [0.04 0.13] | 0.758 |
| lpericalcarine | 1.341 (0.119) | 1.25 (0.127) | 3.722 | <0.001** | [0.04 0.14] | 0.743 |
| rS_parieto_occipital | 1.772 (0.179) | 1.638 (0.182) | 3.731 | <0.001** | [0.06 0.21] | 0.742 |
| lS_circular_insula_ant | 3.022 (0.307) | 2.819 (0.23) | 3.791 | <0.001** | [0.1 0.31] | 0.736 |
| rS_orbital_med_olfact | 2.225 (0.161) | 2.117 (0.136) | 3.682 | <0.001** | [0.05 0.17] | 0.722 |
| lLat_Fis_ant_Vertical | 2.359 (0.201) | 2.219 (0.187) | 3.638 | <0.001** | [0.06 0.22] | 0.719 |
| ltransversetemporal | 1.65 (0.2) | 1.515 (0.178) | 3.606 | <0.001** | [0.06 0.21] | 0.710 |
| lS_suborbital | 2.206 (0.212) | 2.064 (0.183) | 3.609 | <0.001** | [0.06 0.22] | 0.709 |
| lG_postcentral | 1.385 (0.159) | 1.278 (0.144) | 3.572 | <0.001** | [0.05 0.17] | 0.704 |
| rG_oc_temp_med_Lingual | 1.534 (0.108) | 1.45 (0.133) | 3.456 | 0.002* | [0.04 0.13] | 0.698 |
| rS_suborbital | 2.288 (0.216) | 2.138 (0.218) | 3.460 | 0.002* | [0.06 0.24] | 0.688 |
| rS_circular_insula_ant | 2.783 (0.228) | 2.627 (0.223) | 3.462 | 0.002* | [0.07 0.24] | 0.687 |
| lG_and_S_paracentral | 1.399 (0.167) | 1.277 (0.19) | 3.415 | 0.002* | [0.05 0.19] | 0.685 |
| lG_temp_sup_G_T_transv | 1.637 (0.212) | 1.493 (0.208) | 3.451 | 0.002* | [0.06 0.23] | 0.684 |
| lG_insular_short | 3.433 (0.321) | 3.177 (0.433) | 3.342 | 0.002* | [0.1 0.41] | 0.679 |
| lS_central | 1.117 (0.111) | 1.043 (0.107) | 3.400 | 0.002* | [0.03 0.12] | 0.673 |
| rPole_occipital | 1.424 (0.111) | 1.353 (0.099) | 3.409 | 0.002* | [0.03 0.11] | 0.671 |
| lrostralanteriorcingulate | 2.5 (0.137) | 2.398 (0.169) | 3.325 | 0.003* | [0.04 0.16] | 0.671 |
| rG_subcallosal | 2.874 (0.264) | 2.673 (0.338) | 3.308 | 0.003* | [0.08 0.32] | 0.670 |
| rtemporalpole | 3.109 (0.411) | 2.849 (0.367) | 3.375 | 0.002* | [0.11 0.41] | 0.664 |
| rS_circular_insula_inf | 2.87 (0.204) | 2.719 (0.257) | 3.242 | 0.003* | [0.06 0.24] | 0.655 |
| lparsorbitalis | 2.436 (0.19) | 2.311 (0.196) | 3.259 | 0.003* | [0.05 0.2 ] | 0.649 |
| rfrontalpole | 2.215 (0.196) | 2.081 (0.222) | 3.221 | 0.003* | [0.05 0.22] | 0.646 |
| rS_temporal_transverse | 1.794 (0.242) | 1.644 (0.222) | 3.269 | 0.003* | [0.06 0.24] | 0.645 |
| rG_Ins_lg_and_S_cent_ins | 3.019 (0.298) | 2.823 (0.313) | 3.222 | 0.003* | [0.08 0.32] | 0.643 |
| rG_temporal_inf | 2.422 (0.166) | 2.323 (0.151) | 3.172 | 0.004* | [0.04 0.16] | 0.625 |
| lS_temporal_transverse | 1.718 (0.224) | 1.587 (0.194) | 3.165 | 0.004* | [0.05 0.21] | 0.622 |
| rS_central | 1.109 (0.101) | 1.045 (0.105) | 3.093 | 0.005* | [0.02 0.1 ] | 0.616 |
| rlingual | 1.565 (0.095) | 1.502 (0.109) | 3.067 | 0.005* | [0.02 0.1 ] | 0.616 |
| rG_rectus | 2.277 (0.157) | 2.179 (0.173) | 2.973 | 0.007* | [0.03 0.16] | 0.595 |
| rG_temp_sup_Plan_polar | 3.17 (0.328) | 2.993 (0.284) | 2.912 | 0.008* | [0.06 0.3 ] | 0.572 |
| lS_orbital_med_olfact | 2.231 (0.16) | 2.139 (0.172) | 2.796 | 0.01* | [0.03 0.16] | 0.559 |
| lG_front_inf_Orbital | 2.477 (0.196) | 2.364 (0.222) | 2.679 | 0.014* | [0.03 0.2 ] | 0.537 |
| lcaudalanteriorcingulate | 2.32 (0.178) | 2.225 (0.178) | 2.685 | 0.013* | [0.02 0.17] | 0.533 |
| rG_and_S_paracentral | 1.252 (0.181) | 1.162 (0.159) | 2.673 | 0.014* | [0.02 0.16] | 0.526 |
| rentorhinal | 3.169 (0.278) | 3.006 (0.343) | 2.605 | 0.017* | [0.04 0.29] | 0.526 |
| lLat_Fis_ant_Horizont | 2.325 (0.243) | 2.21 (0.223) | 2.486 | 0.022* | [0.02 0.21] | 0.490 |
| rcuneus | 1.356 (0.133) | 1.292 (0.131) | 2.452 | 0.024* | [0.01 0.12] | 0.487 |
| lPole_occipital | 1.435 (0.165) | 1.362 (0.136) | 2.454 | 0.023* | [0.01 0.13] | 0.480 |
| rpericalcarine | 1.141 (0.099) | 1.095 (0.1) | 2.312 | 0.032* | [0.01 0.08] | 0.460 |
| lparahippocampal | 2.228 (0.221) | 2.137 (0.181) | 2.291 | 0.034* | [0.01 0.17] | 0.448 |
| lS_calcarine | 1.606 (0.113) | 1.557 (0.119) | 2.133 | 0.049* | [0 0.1] | 0.426 |
| lG_cingul_Post_dorsal | 2.548 (0.143) | 2.485 (0.155) | 2.112 | 0.051 | [0 0.12] | 0.422 |
| rG_cuneus | 1.283 (0.125) | 1.231 (0.126) | 2.118 | 0.050 | [0 0.1] | 0.421 |
| listhmuscingulate | 2.106 (0.148) | 2.053 (0.118) | 2.002 | 0.063 | [0 0.1] | 0.391 |
| rcaudalanteriorcingulate | 2.29 (0.196) | 2.222 (0.159) | 1.943 | 0.071 | [0 0.14] | 0.380 |
| lS_pericallosal | 1.793 (0.13) | 1.756 (0.125) | 1.471 | 0.177 | [-0.01 0.09] | 0.291 |
| risthmuscingulate | 2.014 (0.149) | 1.973 (0.144) | 1.404 | 0.198 | [-0.02 0.1 ] | 0.278 |
| rS_pericallosal | 1.735 (0.14) | 1.701 (0.111) | 1.351 | 0.215 | [-0.02 0.08] | 0.264 |
| lG_oc_temp_med_Parahip | 2.596 (0.251) | 2.538 (0.194) | 1.334 | 0.220 | [-0.03 0.15] | 0.260 |
| lG_subcallosal | 2.689 (0.339) | 2.613 (0.355) | 1.095 | 0.322 | [-0.06 0.21] | 0.218 |
| rS_calcarine | 1.486 (0.105) | 1.464 (0.111) | 1.002 | 0.362 | [-0.02 0.06] | 0.200 |
| lG_cingul_Post_ventral | 2.083 (0.229) | 2.04 (0.218) | 0.951 | 0.385 | [-0.05 0.13] | 0.188 |
| lentorhinal | 3.146 (0.359) | 3.099 (0.317) | 0.701 | 0.528 | [-0.09 0.18] | 0.138 |
| rG_cingul_Post_ventral | 2.06 (0.207) | 2.064 (0.238) | -0.077 | 0.946 | [-0.09 0.09] | 0.015 |
| **Extracted tract-based fractional anisotropy values** | | | | | | |
| CGH_R | 0.5 (0.053) | 0.473 (0.052) | 2.593 | 0.017* | [0.01 0.05] | 0.515 |
| CGH | 0.495 (0.055) | 0.469 (0.051) | 2.433 | 0.025* | [0 0.05] | 0.480 |
| FX_ST_R | 0.531 (0.044) | 0.514 (0.038) | 2.162 | 0.046* | [0 0.03] | 0.425 |
| CGH_L | 0.49 (0.064) | 0.465 (0.062) | 1.942 | 0.071 | [0 0.05] | 0.385 |
| FXST | 0.527 (0.043) | 0.513 (0.039) | 1.680 | 0.122 | [0 0.03] | 0.331 |
| SFO_R | 0.442 (0.062) | 0.425 (0.036) | 1.721 | 0.113 | [0 0.04] | 0.329 |
| BCC | 0.628 (0.044) | 0.616 (0.034) | 1.535 | 0.159 | [0 0.03] | 0.299 |
| SLF | 0.456 (0.033) | 0.447 (0.022) | 1.506 | 0.167 | [0 0.02] | 0.290 |
| CR_R | 0.423 (0.031) | 0.416 (0.024) | 1.444 | 0.184 | [0 0.02] | 0.281 |
| SLF_L | 0.461 (0.037) | 0.452 (0.027) | 1.448 | 0.184 | [0 0.02] | 0.280 |
| ACR_R | 0.397 (0.035) | 0.388 (0.029) | 1.401 | 0.199 | [0 0.02] | 0.275 |
| CST_R | 0.532 (0.053) | 0.517 (0.061) | 1.346 | 0.216 | [-0.01 0.04] | 0.270 |
| SLF_R | 0.451 (0.033) | 0.443 (0.023) | 1.358 | 0.213 | [0 0.02] | 0.262 |
| SFO | 0.445 (0.058) | 0.432 (0.039) | 1.348 | 0.216 | [-0.01 0.03] | 0.260 |
| CR | 0.422 (0.031) | 0.415 (0.026) | 1.276 | 0.242 | [0 0.02] | 0.250 |
| FX | 0.442 (0.085) | 0.421 (0.099) | 1.128 | 0.307 | [-0.02 0.06] | 0.227 |
| CGC_R | 0.55 (0.068) | 0.535 (0.058) | 1.138 | 0.303 | [-0.01 0.04] | 0.223 |
| ACR | 0.392 (0.034) | 0.385 (0.031) | 1.093 | 0.322 | [-0.01 0.02] | 0.216 |
| SCR | 0.449 (0.041) | 0.441 (0.032) | 1.100 | 0.321 | [-0.01 0.02] | 0.214 |
| PCR | 0.435 (0.031) | 0.429 (0.027) | 1.077 | 0.329 | [-0.01 0.02] | 0.211 |
| IFO_R | 0.486 (0.054) | 0.497 (0.048) | -1.055 | 0.340 | [-0.03 0.01] | 0.208 |
| SCR_R | 0.444 (0.039) | 0.436 (0.032) | 1.051 | 0.341 | [-0.01 0.02] | 0.206 |
| CR_L | 0.421 (0.033) | 0.414 (0.03) | 1.037 | 0.347 | [-0.01 0.02] | 0.205 |
| CC | 0.637 (0.037) | 0.63 (0.031) | 1.026 | 0.352 | [-0.01 0.02] | 0.201 |
| FX_ST_L | 0.523 (0.048) | 0.514 (0.047) | 1.015 | 0.357 | [-0.01 0.03] | 0.201 |
| PCR_R | 0.44 (0.033) | 0.433 (0.029) | 1.010 | 0.359 | [-0.01 0.02] | 0.199 |
| PCR_L | 0.429 (0.034) | 0.423 (0.029) | 0.984 | 0.370 | [-0.01 0.02] | 0.193 |
| SCR_L | 0.454 (0.045) | 0.446 (0.036) | 0.958 | 0.382 | [-0.01 0.02] | 0.187 |
| PTR_L | 0.535 (0.038) | 0.528 (0.033) | 0.935 | 0.392 | [-0.01 0.02] | 0.184 |
| PTR | 0.534 (0.034) | 0.529 (0.032) | 0.890 | 0.417 | [-0.01 0.02] | 0.176 |
| AverageFA | 0.433 (0.026) | 0.429 (0.024) | 0.888 | 0.417 | [-0.01 0.01] | 0.175 |
| IFO | 0.486 (0.049) | 0.493 (0.042) | -0.818 | 0.458 | [-0.03 0.01] | 0.161 |
| EC_R | 0.434 (0.037) | 0.439 (0.027) | -0.820 | 0.458 | [-0.02 0.01] | 0.159 |
| SFO_L | 0.448 (0.063) | 0.44 (0.051) | 0.754 | 0.496 | [-0.01 0.03] | 0.147 |
| ACR_L | 0.388 (0.035) | 0.383 (0.036) | 0.726 | 0.513 | [-0.01 0.02] | 0.144 |
| PTR_R | 0.534 (0.034) | 0.529 (0.036) | 0.693 | 0.533 | [-0.01 0.02] | 0.138 |
| EC | 0.432 (0.038) | 0.437 (0.031) | -0.681 | 0.538 | [-0.02 0.01] | 0.133 |
| ALIC_R | 0.54 (0.045) | 0.535 (0.037) | 0.670 | 0.543 | [-0.01 0.02] | 0.131 |
| CST | 0.531 (0.053) | 0.524 (0.055) | 0.604 | 0.587 | [-0.01 0.03] | 0.120 |
| PLIC_L | 0.661 (0.051) | 0.666 (0.038) | -0.586 | 0.598 | [-0.02 0.01] | 0.114 |
| CGC | 0.565 (0.064) | 0.559 (0.056) | 0.533 | 0.635 | [-0.02 0.03] | 0.105 |
| SS_L | 0.528 (0.04) | 0.532 (0.043) | -0.485 | 0.669 | [-0.02 0.01] | 0.097 |
| SS | 0.526 (0.034) | 0.529 (0.035) | -0.475 | 0.674 | [-0.02 0.01] | 0.095 |
| GCC | 0.584 (0.057) | 0.579 (0.051) | 0.465 | 0.680 | [-0.02 0.03] | 0.091 |
| EC_L | 0.43 (0.042) | 0.434 (0.039) | -0.435 | 0.701 | [-0.02 0.01] | 0.086 |
| RLIC_R | 0.536 (0.032) | 0.539 (0.032) | -0.431 | 0.702 | [-0.02 0.01] | 0.086 |
| UNC_R | 0.519 (0.05) | 0.515 (0.049) | 0.418 | 0.710 | [-0.02 0.02] | 0.083 |
| SS_R | 0.523 (0.032) | 0.526 (0.034) | -0.392 | 0.728 | [-0.02 0.01] | 0.078 |
| PLIC | 0.654 (0.046) | 0.657 (0.035) | -0.367 | 0.744 | [-0.02 0.01] | 0.071 |
| IC_L | 0.587 (0.041) | 0.59 (0.033) | -0.361 | 0.747 | [-0.02 0.01] | 0.070 |
| IFO_L | 0.485 (0.054) | 0.488 (0.048) | -0.326 | 0.772 | [-0.02 0.02] | 0.064 |
| ALIC_L | 0.533 (0.043) | 0.535 (0.041) | -0.251 | 0.830 | [-0.02 0.01] | 0.050 |
| RLIC | 0.544 (0.034) | 0.545 (0.033) | -0.244 | 0.832 | [-0.01 0.01] | 0.048 |
| ALIC | 0.537 (0.043) | 0.535 (0.037) | 0.230 | 0.841 | [-0.01 0.02] | 0.045 |
| UNC_L | 0.495 (0.065) | 0.497 (0.052) | -0.197 | 0.866 | [-0.03 0.02] | 0.038 |
| IC | 0.583 (0.037) | 0.584 (0.03) | -0.163 | 0.891 | [-0.01 0.01] | 0.032 |
| SCC | 0.69 (0.042) | 0.689 (0.038) | 0.152 | 0.896 | [-0.01 0.02] | 0.030 |
| CGC_L | 0.58 (0.061) | 0.582 (0.059) | -0.148 | 0.896 | [-0.03 0.02] | 0.029 |
| UNC | 0.509 (0.05) | 0.508 (0.044) | 0.150 | 0.896 | [-0.02 0.02] | 0.029 |
| PLIC_R | 0.648 (0.044) | 0.649 (0.035) | -0.131 | 0.908 | [-0.02 0.01] | 0.026 |
| CST_L | 0.53 (0.065) | 0.531 (0.06) | -0.103 | 0.928 | [-0.03 0.02] | 0.020 |
| IC_R | 0.578 (0.035) | 0.578 (0.03) | 0.057 | 0.959 | [-0.01 0.01] | 0.011 |
| RLIC_L | 0.552 (0.042) | 0.552 (0.038) | -0.039 | 0.972 | [-0.02 0.02] | 0.008 |

Results are reported as mean (standard deviations). Abbreviations: R, right; L, left; G, gyrus; S, sulcus; sup, superior; inf, inferior; med, medial; lat, lateral; temp, temporal; front, frontal; gm, grey matter; FA, fractional anisotropy; ACR, anterior corona radiata; ALIC, anterior limb of internal capsule; BCC, body of corpus callosum; CC, corpus callosum; CGC, cingulate gyrus; CGH, perihippocampal cingulum tract; CR, corona radiata; CST, cortico-spinal tract; EC, external capsule; FX, fornix; FXST, fornix - stria terminalis; GCC, genu of corpus callosum; IC, internal capsule; IFO, inferior fronto-occipital fasciculus; PCR, posterior corona radiata; PLIC, posterior limb of internal capsule; PTR, posterior thalamic radiation; RLIC, retrolenticular limb of the internal capsule; SCC, splenium of corpus callosum; SCR, superior corona radiata; SFO, superior fronto-occipital fasciculus; SS, sagittal stratum; UNC, uncinate fasciculus.
* p<0.05; **p<0.001

**Table S4. Linear discriminant analysis standardized coefficients for clusters’ labels derived from the UMAP-reduced model.**

| **Clinical scales** | **Coefficients** |
| --- | --- |
| CTQ physical abuse | 0.150 |
| CTQ emotional neglect | 1.002 |
| CTQ emotional abuse | 0.288 |
| CTQ physical neglect | -0.520 |
| CTQ sexual abuse | 0.246 |
| CTQ minimisation/denial | 1.099 |
| BDI Negative Self-Esteem | -1.034 |
| BDI Anergy | 1.080 |
| BDI Dysphoria | -0.121 |

Abbreviations: CTQ, Childhood Trauma Questionnaire; BDI, Beck Depression Inventory

**Figure S1. Results from the stability-based relative clustering approach with UMAP dimensionality reduction** A) Probability density distributions identified by Gaussian Mixture Model and Support Vector Machine. B) Normalized stability plot. Error bars represent the 95% confidence interval for normalized stability from the 10x2 repeated cross-validation. The optimal number of clusters that minimizes the normalized stability was 2, achieving an accuracy of 67%. C) Standardized coefficients of the linear discriminant analysis considering clusters’ labels as fixed factors and BDI and CTQ domains as dependent variables. Positive weights (orange) reflect a higher probability to be assigned to the high-TRD cluster, whereas negative weights (green) indicate a higher probability to belong to the low-TRD cluster. D) Graphical representation of the proportion of TRD patients between the identified clusters.

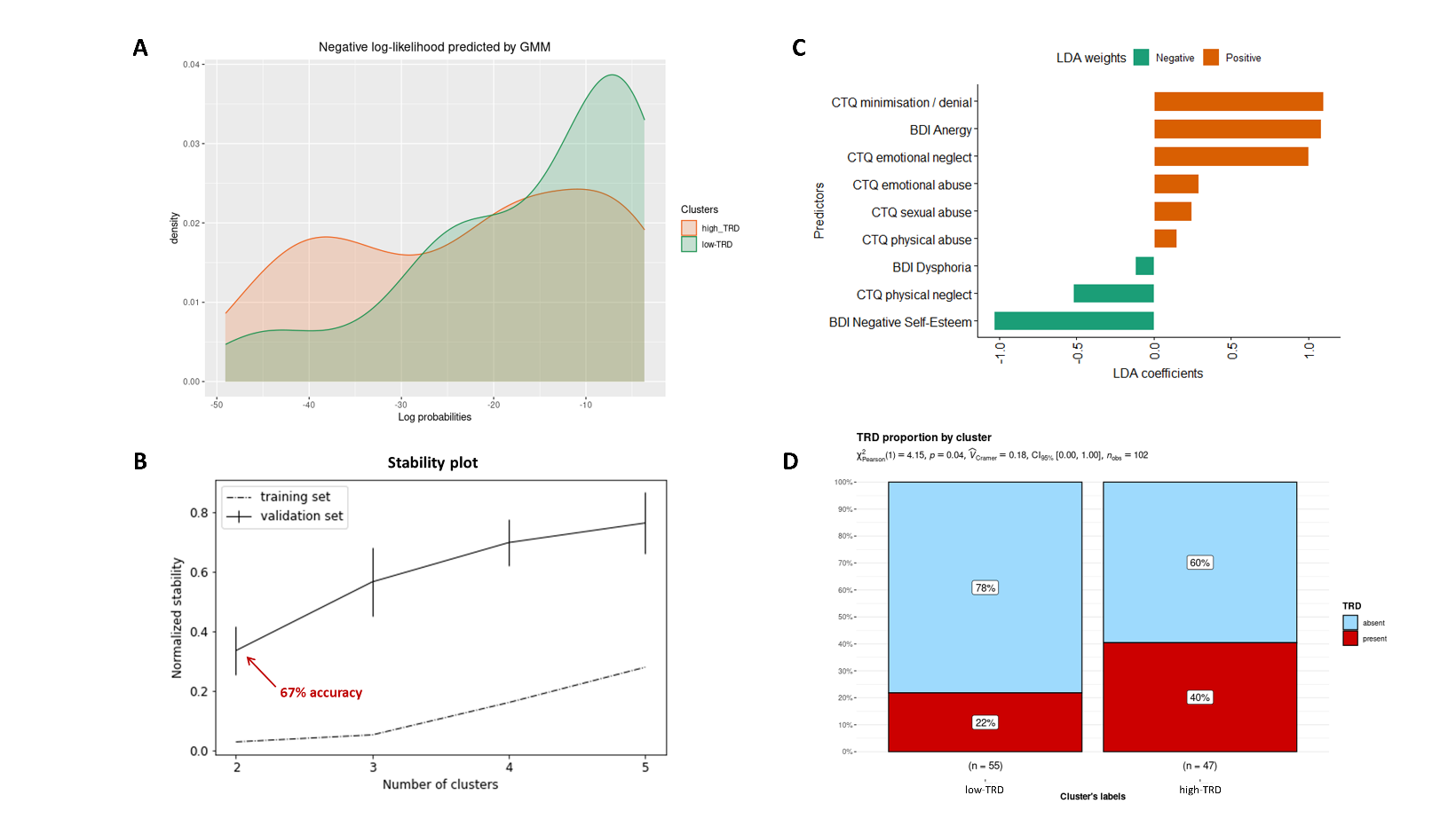

**References**

1.	Hamilton M (1960): A rating scale for depression. *Journal of neurology, neurosurgery, and psychiatry*. 23:56-62.

2.	Sackeim HA (2001): The definition and meaning of treatment-resistant depression. *Journal of Clinical Psychiatry*. 62:10-17.

3.	Landi I, Mandelli V, Lombardo MV (2021): reval: A Python package to determine best clustering solutions with stability-based relative clustering validation. *Patterns*. 2:100228.
